# Clinical and Cost-Effectiveness of an Online Parent-Directed Behavioural Sleep Intervention in addition to Standard Care for Sleep Problems in Children with Epilepsy in the UK: the CASTLE Sleep-E Multicentre, Randomised Controlled Trial

**DOI:** 10.1101/2025.05.14.25326746

**Authors:** P Gringras, A Anilkumar, L Bray, B Carter, T Coffey, G Cook, WAS Hardy, DA Hughes, S Lalnunhlimi, C Morris, H Saron, L Stibbs-Eaton, C. Tudur Smith, L Wiggs, DK Pal

**Affiliations:** Department of Children’s Sleep Medicine, Women’s and Children’s Institute, King’s College London, London, UK; Liverpool Clinical Trials Centre, Institute of Population Health, Faculty of Health and Life Sciences, University of Liverpool, Liverpool, UK; Faculty of Health, Social Care and Medicine, Edge Hill University, Ormskirk, UK; Faculty of Health, Social Care and Medicine, Edge Hill University, Ormskirk, UK.; Liverpool Clinical Trials Centre, Institute of Population Health, Faculty of Health and Life Sciences, University of Liverpool, Liverpool, UK.; Oxford Institute of Applied Health Research, Faculty of Health, Science and Technology, Oxford Brookes University, Oxford, UK; Centre for Health Economics and Medicines Evaluation, North Wales Medical School, Ardudwy, Bangor University, Bangor, Wales; Centre for Health Economics and Medicines Evaluation, North Wales Medical School, Ardudwy, Bangor University, Bangor, Wales.; MRC Centre for Neurodevelopmental Disorders & Basic and Clinical Neurosciences Department, Institute of Psychiatry, Psychology & Neuroscience, King’s College London, London, United Kingdom; Centre for Psychological Research, Faculty of Health, Science and Technology, Oxford Brookes University, Gipsy Lane, Oxford, UK; PenCRU (Peninsula Childhood Disability Research Unit), University of Exeter Medical School, University of Exeter, Exeter UK

## Abstract

**Objectives:** Sleep problems are common in children with epilepsy. We aimed to evaluate the clinical effectiveness and incremental cost-effectiveness of an online resource for parents of children with epilepsy (CASTLE Online Sleep Intervention, or COSI) combined with standard care (SC) versus SC alone over six months.

**Design:** We conducted a multicentre, parallel-group, unblinded, randomised controlled trial with an internal pilot.

**Setting:** 26 UK NHS paediatric epilepsy outpatient clinics.

**Participants:** Children aged 4–12 years with clinician-confirmed epilepsy and caregiver-reported sleep problems.

**Interventions:** Participants were randomly assigned (1:1) via a computer-generated minimisation algorithm to SC or SC plus COSI.

**Main outcome measures:** The primary outcome, assessed at three months, was the Children’s Sleep Habits Questionnaire (CSHQ). Cost-effectiveness was estimated at six months, with utilities from the Child Health Utility 9-Dimensions Index and costs from an NHS and Personal Social Services perspective. Safety analyses included all randomised participants. Analyses were performed on an intention to treat basis.

**Results:** 85 children were enrolled between 30.08.2022 and 18.10. 2023, (42 SC; 43 SC+COSI). At three months, the adjusted mean CSHQ difference between SC+COSI and SC was 3.00 (95% CI 0.06–5.93; p=0.05), indicating no significant superiority. No adverse events were reported. Children in the SC+COSI group showed a mean 16.5-minute reduction in sleep onset latency and parents increased their knowledge. Only 23 (53%) families accessed the core online materials. Incremental mean cost of SC+COSI was £1,232 (95% credibility interval £535–£3,455) with a mean incremental QALY of 0.00 (95% CI -0.03 to 0.04), yielding an incremental cost-effectiveness ratio of £433,167 per QALY gained a (0.04 probability of being cost-effective at the £30,000/QALY threshold).

**Conclusions:** An online Parent-Directed Behavioural Sleep Intervention did not reduce parent-rated sleep problems in children with epilepsy in the NHS context. However, improved objective sleep onset latency and enhanced parental knowledge suggest that the underlying behaviour change techniques have merit. Blended delivery through specialist nurses or similar support may improve engagement and outcomes.

**Trial registration:** ISRCTN13202325.

**Funding:** NIHR Programme Grant for Applied Research RP-PG-0615-20007.

## Introduction

### Background and Rationale

There is a movement towards a more holistic clinical approach to the management of childhood epilepsy, incorporating aspects that are important to children and their parents such as mental health and sleep.^1^ This gradual broadening of emphasis presents a practical challenge because the NHS workforce is not equipped to meet the demands of managing these issues within current resources.^2^ Sleep problems, in particular, represent a substantial and rising public health issue,^3^ and are common in children. Children’s sleep problems are associated with impairments in learning, behaviour, daytime functioning, and overall quality of life, as well as disruptions to their family’s sleep, functioning and relationships.^4^ Sleep can be a source of considerable anxiety in children with epilepsy (CWE) because of the fear of seizures.^5^ Unsurprisingly, parents report that sleep problems are 12 times more common in CWE, even in the absence of nocturnal seizures.^6^ Furthermore, sleep disruption may also exacerbate seizure activity by affecting sleep architecture and quality, creating a vicious cycle sometimes leading to pseudo drug-resistance.^7^ Sleep has been identified as a vital concern for young people with epilepsy^1^ and is recognised by people with lived experience as a research priority.^8^ However, there exists an enormous imbalance between patient needs and children’s sleep-related healthcare provision in the UK.^2^ Accessible and scalable approaches to address sleep problems in this population are therefore urgently needed, with the potential for substantial health and economic benefits.

Tailored sleep behaviour intervention methods are necessary for CWE because of specific clinical considerations.^5^ In CWE, insomnia-related sleep problems typically include difficulty with sleep initiation (settling and falling asleep), maintenance (night or early morning wakings), short sleep duration, daytime sleepiness, and sleep-related anxieties. Several parent-implemented, face-to-face behavioural management techniques exist for treating behavioural insomnia in typically developing children.^9^ These techniques have also been used successfully with children who have a range of neurodevelopmental disorders, including ADHD and autism but their use in CWE remains under-explored. Interventions delivered online are now a well-proven strategy to improve access to services, particularly in mental health care. Behavioural sleep management is especially suited to eHealth adaptation because techniques can be standardised and delivered in modular form. Evidence supports the effectiveness of online sleep management programmes for typically developing children and some clinical populations.^10^ With this rationale, we developed CASTLE Online Sleep Intervention (COSI) - an online, parent-directed, modular behavioural sleep intervention specifically for children with epilepsy.^5^ This study aimed to evaluate the clinical and incremental cost-effectiveness of standard care (SC) plus COSI compared with SC alone for children with epilepsy in NHS context.

### Hypothesis

SC+COSI will improve the mean total parent-reported child sleep problem [Children’s Sleep Habits Questionnaire (CSHQ)] score by 6.25 points from baseline to 3-months compared with SC alone in children with epilepsy aged 4-12 years.

## Methods

### Study Design

This was a multicentre, parallel-group, unblinded randomised controlled trial with internal pilot conducted across 26 hospital paediatric outpatient clinics in the UK. The study was prospectively registered on the International Standard Randomised Controlled Trial Number Registry (ISRCTN13202325). The study protocol has been published in the NIHR Library. Reporting follows guidance from the Consolidated Standards of Reporting Trials (CONSORT) 2010^11^, CONSORT extension

for non-pharmacological trials 2017^12^ and Consolidated Health Economic Evaluation Reporting Standards 2022 (CHEERS 2022).^13^

### Patient and Public Involvement

The study was designed and conducted in collaboration with the research programme Advisory Panel (AP). The advisory panel (AP) consisted of three children with epilepsy, ten parents of children with epilepsy and one adult with epilepsy who has lived with epilepsy since childhood. Two of the AP were consulted at the grant application stage and were co-applicants on the programme grant. The AP advised on all aspects of the study design, including the outcome measures. Patient-facing documents were co-designed with the AP to ensure that the language and terminology used were suitable, consistent, and understandable. Strategies and materials for public dissemination were developed in collaboration with the AP.

### Sample size

The trial aimed to recruit 110 participants, sufficient to detect a clinically meaningful difference of 6.25 with a standard deviation of 8.9 in 3-month CSHQ scores with 90% power at a 5% significance level, accounting for potential attrition of 10%. However, owing to diminished recruitment following the Covid-19 pandemic, 85 participants were randomised, which provided 80% power to detect change in the primary endpoint.

### Participants

Children aged 4 to 12 years were recruited from 26 NHS outpatient paediatric epilepsy clinics across the United Kingdom. Eligibility criteria included clinician-confirmed epilepsy and parent-reported sleep problems.^14^ Exclusion criteria included children with moderate/severe learning disabilities or families without sufficient English to understand the intervention materials.

### Randomisation

Participants were randomised with a 1:1 allocation ratio using a computer-generated minimisation algorithm with a random element of 0.8 balancing for site and sleep medication.

### Blinding

Due to the nature of the intervention, blinding was not feasible for participants, caregivers, or clinical outcome assessors. Statisticians were blinded until the point of analysis.

### Interventions

Participants randomised to the control Standard Care (SC) group received usual care provided by their paediatric epilepsy services. Participants randomised to the experimental SC+COSI group received SC plus access to an online parent-directed behavioural sleep intervention. The COSI intervention is a self-paced, tailored, modular e-learning package incorporating sleep information (modules A-B) and 20 evidence-based behavioural sleep strategies (modules D-J).^5^ Through co-production with parents of children with epilepsy, adaptations were made to the content and presentation of standard intervention material to acknowledge and emphasise key seizure-specific issues to best meet parents of CWE’s needs.^5^ Adaptations included embedding parent and child experiences in the intervention in short animations, adding information requested by parents, such as the links between sleep and seizures and managing child and parental anxieties around sleep, and sudden unexpected death in epilepsy, as well as developing functionality to personalise content delivery.

### Assessment

Parent-reported and child-reported outcomes were assessed at baseline, three months and six months. Seizure outcomes were collected by clinicians at follow-up appointments and entered into a central, secure REDCAP database. A revised study design was implemented to extend the recruitment period,

which ensured that every patient had primary outcome data collected at the three-month follow-up visit, but a subset of patients did not receive a six-month follow-up visit.

### Outcomes

#### Primary Outcome

The primary outcome was the total parent-reported child sleep problem score on the (CSHQ) at 3 months post-randomisation^15^.

#### Secondary Outcomes

**Children:** Time to 6-month seizure remission; time to first seizure; total score on Strengths and Difficulties Questionnaire (SDQ); SleepSuite overnight memory consolidation assessment; actigraphy variables (total sleep time, sleep latency, sleep efficiency), Children with Epilepsy Quality of Life (CHEQOL): Parent and Child Questionnaire, total sickness-related school absence days.

**Caregivers:** Knowledge about Sleep in Childhood (KASC) scale score, Insomnia Severity Index (ISI); Hospital Anxiety and Depression Scale (HADS), WHO-5 Well-Being Index, Parenting Self-Agency Measure (PSAM).

#### Health Economic Outcome

Incremental cost-effectiveness ratio (ICER), calculated using costs from the NHS and Personal Social Services perspective and QALYs derived from the Child Health Utility 9 Dimension Index (CHU9D).

#### COSI Engagement Assessments

Descriptive, e-learning data about parents’ use of the intervention website, including how many modules parents looked at and how many times they viewed each individual module. Parents were defined as COSI engagers if they accessed one or more of the modules that included advice about child sleep-behaviour change strategies (D-J), on one or more occasions.

#### COSI Evaluation Outcome

COSI was evaluated via an embedded online questionnaire, which became available 3 months after randomisation to the COSI arm of the trial. Families were prompted to complete this via a text message and, if required, by a phone call reminder after a week of non-completion. The evaluation questionnaire assessed the following aspects of COSI:

i. Functionality of website (e.g. the frequency with which families could access the different features of COSI such as viewing videos). Responses were on a 5-point scale of ‘never’ (0) to ‘always’ (4).
ii. Parental approval of COSI via the question “Would you recommend this sleep programme to other families who have children with epilepsy?”. Responses were on a 5-point scale of ‘never’ (0) to ‘always’ (4).
iii. Use of COSI content - for each of the 20 individual sleep management strategies included in COSI, parents reported if they had used the technique (yes/no response) and, if ‘yes’, how helpful they found this on a 5-point scale (0-4) from ‘very unhelpful’ to ‘very helpful’ or, if ‘no’, they indicated whether this was because they were already using this method, because they had unsuccessfully tried it in the past or because they chose not to.

#### Qualitative Research Assessments

The aim was to critically explore the experiences of children and parents/caregivers in relation to living with epilepsy, sleep (importance, challenges, management strategies), use of COSI and factors related to participation in the trial (e.g. why participate, challenges to sustaining participation, and use of actigraphs). A subset of families (maximum 60, split between the two groups) were invited to participate in remote audio or audio-video interviews within 3 weeks of completion of other data collection at 3 and 6 months after randomisation.

#### Internal Pilot

Recruitment rate, consent rate, follow-up rate, intervention failure rate, and completeness of data for the primary outcome were assessed during a 6-month internal pilot. Pre-specified progression criteria for continuation to the full trial comprised: Green - Continue if at least 22 participants had been randomised; Amber - Implement additional recruitment strategies if number of participants randomised between 15 and 21; Red - If recruitment <15 discuss ending the trial with the oversight committees.

### Statistical Methods

Baseline categorical data are presented using counts and percentages and continuous data presented as median with range. Primary outcome data were analysed using intention-to-treat (ITT) principles. Total CSHQ score was calculated using the 5-point scoring approach with higher score reflecting greater sleep problems. As the 22-item CSHQ tool had not been validated, the overall Cronbach alpha was calculated as measure of internal consistency and individual items that showed increase in Cronbach’s alpha on deletion and very low “Corrected Item-Total Correlation” (< .20) were omitted from the total score value (items 16,19,20) for the primary analysis. Sensitivity analyses were conducted to examine the effect of alternative scoring approaches and to make comparisons with the more widely used 33-item CSHQ questionnaire^16^ (see Appendix 1).

Continuous outcomes were compared at relevant time points (3 months or 6 months) using linear mixed effects regression models with fixed effects included for intervention, baseline measure, baseline sleep medication status (Y/N) and random effects for centre. Time to event outcomes were analysed using a Cox proportional hazards model including intervention, baseline sleep medication status (Y/N) and centre effects. Analyses were based on complete cases, which are unbiased when data is missing completely at random^17^. The adjusted treatment effect estimates (mean difference for continuous outcomes and hazard ratio for time to event outcomes) comparing SC+COSI against SC are presented with a 95% confidence interval with statistical tests conducted at the 5% two-sided significance level.

Unadjusted analyses were examined for completeness. Analyses included patients with follow-up data captured within ±1 month of the timepoint of interest. Owing to the revised study design, a subset of participants did not achieve a full 6 months of follow-up and sensitivity analyses were conducted that included participants with data collected at > 1 month after the timepoint of interest.

### Health Economic Evaluation

The cost-effectiveness analysis (Appendix 2) used patient-level data to compare SC+COSI with SC alone on an ITT basis, over the 6±1 month trial period. All resource use was measured, irrespective of whether it was related to epilepsy. Within-trial resource use was obtained from routine Patient Level Information and Costing System (PLICS) data, trial case report forms, and resource use questionnaires that were administered at or shortly after baseline, randomisation, follow-up at 3 month (± 1-month), and follow-up at 6 month (± 1-month) post-randomisation visits. Unit costs were obtained from standard sources, valued in pound sterling (£) based on 2021-22 prices, and with inflation indices applied as necessary (Appendix 2). The cost of COSI was estimated as the cost of developing the intervention divided by the number of users. Utilities were estimated from responses to the CHU9D and applying UK tariffs.

Missing data were treated as missing at random and imputed using multiple imputation with chained equations with two-level predictive mean matching to create 70 complete data sets. The complete datasets were analysed using a multivariate Bayesian regression model, with a log-link function and hurdle Gamma distribution for costs and an identity link with Gaussian distribution for QALYs. The model included covariates for treatment allocation with random effects based on treatment site, use of sleep medication at baseline (binary), baseline costs, and baseline utility. Posterior draws from the joint distribution of incremental costs and QALYs were used to analyse uncertainties in the economic evaluation, presenting the probability that the SC+COSI was cost-effective across a range of willingness-to-pay thresholds. Sensitivity analyses were used to explore key cost drivers, the impact of different cost perspectives, varying the cost of COSI, and the inclusion of spillover utility for primary carers. The robustness of modelling assumptions and methods were assessed with respect to handling missing data, and the inclusion of participants who were randomised but too late to be followed up for the 6±1 month trial period. The analysis was conducted in R (Version 4.4.1).

### Protocol Conduct

Three substantial amendments were made and 14 deviations from the protocol were recorded. The trial was conducted and reported according to the published protocol, which was approved by an independent programme steering committee and a data monitoring and ethics committee. East Midlands – Nottingham 1 ethics committee of the Health Research Authority, UK gave ethical approval for this work. The data monitoring and ethics committee reviewed trial data and conduct at regular intervals throughout the trial. The trial was prospectively registered with the ISRCTN (ISRCTN13202325).

## Results

85 children were enrolled between 30.08.2022 and 18.10. 2023, (42 SC; 43 SC+COSI). Eighty-five children were enrolled and randomised (42 to SC, 43 to SC+COSI). Participant flow is detailed in Figure 1.

Baseline characteristics of child participants are provided in Table 1. Groups were comparable at baseline regarding age, sex, ethnicity, diagnosis of epilepsy syndrome, and baseline sleep problem scores.

**Table 1.** Baseline characteristics of trial participants.

| Characteristic | Category/Measure | SC+COSI<br>(N=43) | SC<br>(N=42) | Overall<br>(N=85) |
| --- | --- | --- | --- | --- |
| Gender | Female<br>Male | 13 (30.23%)<br>30 (69.77%) | 16 (38.10%)<br>26 (61.90%) | 29 (34.12%)<br>56 (65.88%) |
| Age (years) | Minimum<br>Median<br>Maximum | 4<br>8<br>12 | 4<br>9.5<br>12 | 4<br>9<br>12 |
| Age (years): n (%) | 0-<6<br>6-<12<br>>=12 | 11 (25.58%)<br>32 (74.42%)<br>0 | 7 (16.67%)<br>35 (83.33%)<br>0 | 18 (21.18%)<br>67 (78.82%)<br>0 |
| Ethnicity | White<br>Indian subcontinent<br>Chinese or other Asian<br>Black/Afro-Caribbean<br>Arabic<br>Any other ethnic group | 39 (90.70%)<br>0<br>1 (2.32%)<br>0<br>0<br>3 (6.98%) | 37 (88.10%)<br>0<br>0<br>0<br>0<br>5 (11.90%) | 76 (89.41%)<br>0<br>1 (1.18%)<br>0<br>0<br>8(9.41%) |
| Child diagnosed with a particular epilepsy syndrome? | Yes<br>No<br>Missing | 31 (72.09%)<br>8 (18.60%)<br>4 (9.30%) | 31 (73.81%)<br>4 (9.52%)<br>7 (16.67%) | 62 (72.94%)<br>12 (14.12%)<br>11 (12.94%) |
| Child taking sleep medication? | Yes<br>No | 10 (23.26%)<br>33 (76.74%) | 8 (19.05%)<br>34 (80.95%) | 18 (21.18%)<br>67 (78.82%) |

### Outcomes and Estimation

#### Primary Outcome

All 85 randomised participants completed the baseline CSHQ questionnaire but only 72 (85%) also completed the 3-month CSHQ form within the required ±1 month follow-up window and were included in the primary analysis. Among the 34 participants who completed the baseline and 3-month CSHQ scores within the SC+COSI arm, 19 (56%) were COSI engagers. The mean CSHQ score at 3 months did not differ significantly between SC and SC+COSI groups (mean difference = 3.00(1.46); 95% CI: 0.06-5.93, p= (0.05)), Table 2.

**Table 2.** Primary endpoint analysis: Total CSHQ score at 3 months (±1 month) from a linear mixed effects regression model based on 19 CSHQ items (5-point scoring).

| **Treatment group** | **N** | **Sleep medication at baseline: Yes (%)** | **Mean (SD) CSHQ score at baseline** | **Mean (SD) CSHQ score at 3 months** | **Point Estimate (SE)** | **95% CI** | **p-value** |
| --- | --- | --- | --- | --- | --- | --- | --- |
| SC+COSI | 34 | 8 (23.5%) | 48.00 (11.15) | 48.18 (12.64) | 3.00 (1.46) | 0.06, 5.93 | 0.05 |
| SC | 38 | 8 (22.1%) | 47.45 (12.52) | 44.61 (11.83) |  |  |  |
| Total | 72 | 16 (22.2%) | 47.71 (11.81) | 46.29 (12.26) |  |  |  |

#### Secondary Outcomes

29 families provided complete actigraphy data at baseline and 3-month follow-up. Children in the SC+COSI group fell asleep on average -16.35 minutes faster (95% CI: -30.63 to -2.07; p=0.03) and 75% of these families were COSI engagers. Parental sleep knowledge improved significantly in the SC+COSI group, 2.03 (0.48) (95% CI: 1.01 to 3.05; p= <0.001); no significant differences were observed in seizure control or other secondary outcomes (Tables 3-5).

**Table 3.** Time to event seizure outcomes.

|  | N (%)<br>COSI | N (%)<br>SC | N (%)<br>Total | Number of<br>Remissions<br>SC+COSI | Number of<br>Remissions<br>SC | Number of<br>Remissions<br>Total | Median Time<br>to event, days | Hazard Ratio<br>(HR) | 95% CI | p-value |
| --- | --- | --- | --- | --- | --- | --- | --- | --- | --- | --- |
| **Time to 6-month**<br>**seizure remission** | 30 | 31 | 61 | 12 (38.70%) | 17 (56.67%) | 29 (47.54) | NA | 1.80 | 0.50,6.39 | 0.37 |
|  | N (%)<br>COSI | N (%)<br>SC | N (%)<br>Total | Number of<br>Seizures<br>SC+COSI | Number of<br>Seizures<br>SC | Number of<br>Seizures<br>Total | Median Time<br>to event, days | Hazard Ratio<br>(HR) | 95% CI | p-value |
| **Time to first**<br>**seizure** | 40 | 41 | 81 | 22 (55%) | 18 (43.9%) | 40 (49.38%) | 130 | 1.18 | 0.56. 2.50 | 0.66 |

**Table 4.** Secondary non-seizure outcomes.

| Secondary Outcome | Timepoint (months) | N COSI | N SC | N Total | Mean (SD) at baseline COSI | Mean (SD) at baseline SC | Mean (SD) Baseline Total | Mean (SD) at timepoint COSI | Mean (SD) at timepoint SC | Mean (SD) timepoint Total | Point Estimate (SE) | 95% CI | p-value |
| --- | --- | --- | --- | --- | --- | --- | --- | --- | --- | --- | --- | --- | --- |
| KASC (parent) | 3 | 16 | 24 | 40 | 11.50 (2.34) | 10.54 (2.19) | 10.93 (2.27) | 13.50 (1.15) | 11.08 (1.95) | 12.05 (2.05) | 2.03 (0.48) | 1.01, 3.05 | <0.001 |
| ISI (parent) | 3 | 16 | 24 | 40 | 9.38 (5.38) | 12.88 (4.48) | 11.48 (5.10) | 8.56 (6.23) | 11.79 (6.32) | 10.50 (6.41) | 0.15 (1.45) | -2.92, 3.23 | 0.92 |
|  | 6 | 9 | 11 | 20 | 9.44 (3.28) | 13.73 (5.14) | 11.80 (4.82) | 11.00 (5.59) | 14.09 (7.30) | 12.70 (6.61) | 1.97 (1.84) | -3.14, 7.08 | 0.34 |
| Actigraphy (total sleep time minutes): Parents | 3 | 13 | 18 | 31 | 380.41 (82.77) | 405.83 (56.85) | 395.17 (68.81) | 377.01 (80.53) | 409.14 (48.10) | 395.67 (64.53) | -12.48 (15.63) | -46.25, 21.30 | 0.44 |
| Actigraphy (sleep latency minutes): Parents | 3 | 13 | 17 | 30 | 24.32 (22.73) | 20.34 (16.16) | 22.07 (19.02) | 26.58 (24.80) | 18.34 (15.93) | 21.91 (20.29) | 4.94 (5.32) | -6.66, 16.53 | 0.37 |
| Actigraphy (sleep efficiency minutes): Parents | 3 | 13 | 17 | 30 | 78.21 (13.33) | 79.85 (12.84) | 79.14 (12.85) | 77.66 (13.89) | 82.46 (10.07) | 80.38 (11.90) | -3.15 (2.44) | -8.47, 2.18 | 0.22 |
| Actigraphy (total sleep time minutes): Child | 3 | 14 | 19 | 33 | 475.46 (64.37) | 471.49 (46.51) | 473.17 (53.89) | 468.61 (44.18) | 469.1 (40.88) | 468.89 (41.63) | -0.05 (9.58) | -20.74, 20.64 | 1.00 |
| Actigraphy (sleep latency minutes): Child | 3 | 12 | 17 | 29 | 39.71 (38.07) | 35.08 (16.51) | 37 (27.03) | 34.18 (27.84) | 40.79 (28.51) | 38.06 (27.93) | -16.35 (6.41) | -30.63, -2.07 | 0.03 |
| Actigraphy (sleep efficiency minutes): Child | 3 | 12 | 17 | 29 | 74.67 (10.28) | 78.48 (7.73) | 76.90 (8.91) | 76.38 (6.83) | 77.77 (7.06) | 77.20 (6.88) | 1.10 (1.52) | -2.30, 4.50 | 0.49 |
| CHEQOL | 6 | 5 | 10 | 15 | 77.4 (7.30) | 64.5 (17.44) | 68.8 (15.83) | 82.8 (17.54) | 69.3 (18.60) | 73.8 (18.81) | 2.36 (6.74) | -26.65, 31.36 | 0.76 |
| **Total sickness related school absence days:** | 3 | 36 | 38 | 74 | 5.28 (6.9) | 6.39 (16.01) | 5.85 (12.37) | 4.14 (5.77) | 4.89 (11.65) | 4.53 (9.21) | -0.31 (1.04) | -2.40, 1.78 | 0.76 |
|  | 6 | 22 | 26 | 48 | 5.18 (6.4) | 7 (17.78) | 6.17 (13.69) | 5.05 (6.74) | 5.38 (11.02) | 5.23 (9.21) | 0.50 (2.20) | -4.03, 5.03 | 0.82 |
| **Total SDQ score** | 3 | 18 | 25 | 43 | 14.39 (7.48) | 18.00 (8.57) | 16.49 (8.24) | 13.78 (7.38) | 16.04 (9.19) | 15.09 (8.46) | 1.25 (0.23) | -0.89, 3.39 | 0.23 |
|  | 6 | 9 | 11 | 20 | 13.44 (5.94) | 21.91 (7.38) | 18.10 (7.89) | 13.33 (5.98) | 20.91 (6.66) | 17.50 (7.30) | -0.78 (0.95) | -3.41, 1.86 | 0.46 |
| **HADS (parent)** | 3 | 16 | 24 | 40 | 14.94 (8.14) | 15.75 (6.42) | 15.43 (7.07) | 14.19 (7.82) | 14.25 (8.89) | 14.23 (8.37) | 0.96 (1.26) | -1.71, 3.63 | 0.46 |
|  | 6 | 9 | 11 | 20 | 14.56 (7.63) | 17.45 (5.22) | 16.15 (6.41) | 16.67 (8.26) | 20.09 (6.93) | 18.55 (7.56) | -1.49 (2.47) | -8.36, 5.37 | 0.58 |
| **WHO-5 (parent)** | 6 | 9 | 11 | 20 | 72.67 (12.11) | 67.00 (18.51) | 69.55 (15.83) | 78.22 (20.47) | 71.73 (19.40) | 74.65 (19.63) | 0.87 (5.09) | -11.58, 13.32 | 0.87 |
| **PSAM (parent)** | 3 | 16 | 24 | 40 | 5.64 (0.83) | 5.33 (0.82) | 5.46 (0.83) | 5.41 (0.81) | 5.41 (0.91) | 5.41 (0.86) | -0.31 (0.15) | -0.63, 0.02 | 0.06 |
|  | 6 | 9 | 11 | 20 | 5.64 (0.71) | 4.98 (0.84) | 5.28 (0.83) | 5.93 (0.82) | 4.98 (0.78) | 5.41 (0.92) | 0.41 (0.24) | -0.29, 1.12 | 0.18 |

**Table 5.** Sleep related memory consolidation outcomes.

| Secondary Outcome | Timepoint (months) | N COSI | N SC | N Total | Mean (SD) at baseline COSI | Mean (SD) at baseline SC | Mean (SD) Baseline Total | Mean (SD) at timepoint COSI | Mean (SD) at timepoint SC | Mean (SD) timepoint Total | Point Estimate (SE) | 95% CI | p-value |
| --- | --- | --- | --- | --- | --- | --- | --- | --- | --- | --- | --- | --- | --- |
| Sleep Suite: Total number of correctly identified animals in Animal task | 3 | 11 | 10 | 21 | -0.09 (1.92) | 1.20 (1.03) | 0.52 (1.66) | -0.36 (1.29) | 0.10 (1.29) | -0.14 (1.28) | 0.06 (0.60) | -1.74, 1.61 | 0.92 |
| Sleep Suite: Mean reaction time in emotional face task (seconds) | 3 | 14 | 11 | 25 | -0.08 (0.17) | -0.04 (0.18) | -0.06 (0.17) | -0.02 (0.22) | -0.07 (0.28) | -0.04 (0.24) | 0.04 (0.10) | 0.21, 0.30 | 0.70 |
| Sleep Suite: Mean intraindividual coefficient in emotional face task | 3 | 14 | 11 | 25 | 0.02 (0.11) | 0.04 (0.14) | 0.03 (0.12) | 0.00 (0.08) | -0.01 (0.11) | -0.01 (0.09) | 0.03 (0.04) | -0.06, 0.12 | 0.49 |
| Sleep Suite: Number of omissions in the bubble task for the emotion section | 3 | 14 | 11 | 25 | -3.36 (7.93) | -5.82 (6.26) | -4.44 (7.21) | -1.43 (7.48) | -2.64 (8.30) | -1.96 (7.71) | 1.46 (3.0) | -5.87, 8.79 | 0.64 |
| Sleep Suite: Number of commission errors in the bubble task for the emotion section | 3 | 14 | 11 | 25 | -3.29 (27.43) | -1.36 (24.62) | -2.44 (25.71) | -5.57 (20.40) | -0.91 (25.83) | -3.52 (22.56) | -5.19 (8.66) | -26.37, 15.99 | 0.57 |
| Sleep Suite: Maze Task | 3 | 14 | 11 | 25 | -3.30 (31.60) | -24.93 (63.61) | -12.82 (48.45) | -17.81 (31.66) | -20.65 (34.59) | -19.06 (32.30) | 3.12 (14.28) | -31.81, 38.06 | 0.83 |

#### COSI Engagement Outcome

Of the 43 families assigned to the COSI arm, 33 logged-on. Ten of these only viewed the introductory modules (A & B). Twenty-three were COSI engagers (i.e. accessed behaviour-change content in modules D-J on one or more occasions), whilst 20 families did not engage with the core behaviour change material.

#### COSI Evaluation

Twelve parents completed the evaluation questionnaire. However, one had not accessed any content and so their responses were excluded. The remaining 11 parents were COSI engagers.

i. Functionality: all elements of COSI functioned well (mean scores all ≥ 3.6, maximum 4.0).
ii. Parental Approval: The mean parental approval score was 3.6 (maximum 4.0), indicating families would strongly recommend COSI to other parents of children with epilepsy.
iii. Use of COSI: All 20 sleep behaviour change strategies were reportedly used (19/20 strategies were used by multiple families). Of the 94 reports of a strategy being used as a result of COSI, 81 (86.2%) of these were reported to be ‘helpful or ‘very helpful’. The other 13 (13.8%) uses were reported to have had no impact and none were reported to make sleep worse. Where strategies were not used and an explanation was given (n=122) this was because families were already using this (n=76), chose not to use it (n=26) or had tried the strategy unsuccessfully in the past (n=18).

### Qualitative Interview Outcomes

Twenty-two families participated in interviews: 13 in the SC arm (7 boys, 6 girls; 4-13yrs, mean 9.3yrs) and nine in the SC+COSI arm, of which six families were categorized as COSI engagers (4 boys, 2 girls; 8-12yrs, mean 9.8yrs) and three as COSI non-engagers (1 boy, 2 girls; 10-13yrs, mean 11.3yrs). Two of the three COSI non-engagers cited family crises as reasons for non-engagement. Five COSIengager parents talked of being familiar with some aspects of COSI content, noting that suggested routines and practices were already “well ingrained” or had been “advised before”. Despite familiarity, benefits did accrue such as success using a “star chart to try to encourage him to sleep in his own bed” and introducing “wind down” time or the potential for introducing “more relaxation” practices. One boy thought COSI practices were “helping me a bit” and believed they would help his sleep “improve over the long-term”. Perceived gaps in COSI information included advice about managing “restless … tossing and turning” sleep.

Although COSI content was reported as “high quality”, one father reported feeling “overloaded with information”, some of which he felt conflicted with other sources. Time pressures and distractions affected engagement. Parental sleep disruption and tiredness was problematic for four COSI engager families - problems included falling asleep, staying asleep, waking up or restless sleep.

### Health Economic Outcome

Fifty-eight participants (29 in each group) had 6±1-month follow-up data prior to the trial closing and therefore included in the base case analysis; all 85 randomised participants were included in sensitivity analyses.(Appendix 2) There were missing cost and utility data at each timepoint, the proportions of missing data in the SC+COSI group were generally higher than in the SC group.^18^ Most of the costs were associated with secondary care and COSI^18^ (Table 6). Unadjusted baseline costs were higher in the SC group and baseline utilities were similar in both groups.^18^ In the adjusted, base case analysis, mean total within-trial costs were £2,021 [95% credibility interval (CrI): 1,352; 3,455] for the SC+COSI group and £1,232 [95% CrI: 535; 2,183] for the SC group. SC+COSI was associated with 0.42 [95% CrI: 0.39; 0.45] QALYs compared with 0.41 [95% CrI: 0.38; 0.45] for SC alone. SC+COSI was more costly than SC alone, with incremental costs of £1,232 [95% CrI: 535; 2,183] and there was a difference of 0.0028 [95% CrI: - 0.0326; 0.0368] QALYs compared to SC alone. The resulting ICER is £433,167 per QALY gained (Table 6). At willingness to pay thresholds of £20,000 and £30,000 per QALY, SC+COSI had negative net monetary benefits (NMBs), and probabilities of 0.01 and 0.04, respectively, of being cost-effective (Figure 2). The sensitivity analyses all supported the base case analysis, with the lowest ICER being £70,246 per QALY gained. SC+COSI was associated with negative NMB even when the cost of COSI was zero and was dominated by SC in two sensitivity analyses (Appendix 2).

**Table 6.**
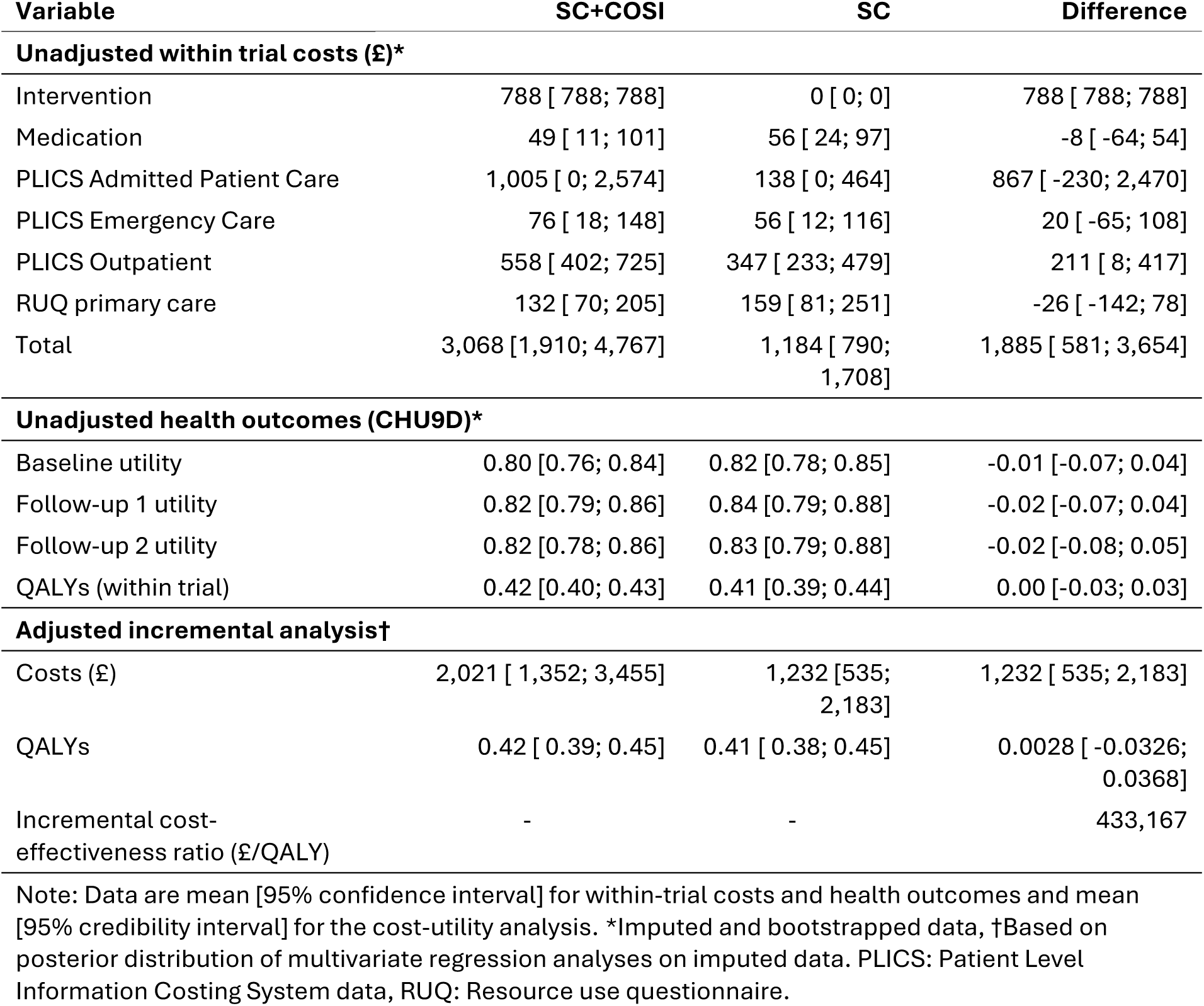
Costs, health outcomes, and results of the cost-effectiveness analysis.

### Adverse Events

No serious adverse events were reported in the trial.

## Discussion

This is the first clinical trial of an online parent-directed behavioural sleep intervention for children with epilepsy and tested the real-life feasibility of transferring the tools for management of sleep problems directly to parents/caregivers without any professional support. The intervention did not reduce parent-rated sleep problems in children with epilepsy in the NHS context. However, improved objective sleep onset latency and enhanced parental knowledge suggest that the underlying behaviour change techniques may have merit.

We were addressing the question of whether sleep problems can be tackled in this population across the NHS without input from either specialist or non-specialist staff. We found that adding COSI to standard care did not improve subjective parent-reported child sleep problems compared to standard care alone and was not cost-effective. Uptake was low (53%) in line with other digital health interventions in mental health and might partially explain the lack of effectiveness.^19^ Delivering the intervention to a child proxy (parent/caregiver) represents another potential source of effect dilution. Objectively though, the intervention brought forward child sleep onset by 16.35 minutes, a clinically significant improvement.^20^ Caregivers who engaged with COSI would strongly recommend the intervention to other parents, and they found 86% of the techniques helpful or very helpful. COSI also improved parental knowledge. We conclude that the underlying behavioural change techniques are efficacious but at the population level an online parent-based intervention does not result in parent-reported improvements in child sleep problems. The trial informs public health strategy for managing sleep problems in children with complex needs and suggests alternative future implementation approaches such as sleep training of the clinical care team.

When behaviour-change techniques are translated to digital format, there is often a trade-off between access and scalability against low user engagement and inconsistent adherence - which are common issues in remote self-guided programmes^19^. With COSI, engagement (logging on) and attrition (progressing to and revisiting core modules) were equally problematic. Sometimes known as Eysenbach’s Law^21^, this feature of eHealth trials is a distinct characteristic. Additionally, web metrics alone do not reliably indicate how participants use behaviour change content, making it difficult to assess actual adherence – unfortunately evaluation questionnaires were only completed by 11 families. The subpopulation for whom the intervention brought forward child sleep onset were largely (75%) COSI engagers – a high level of dropout underestimates the effect of the intervention among users who continue to use it.^21^ Digital interventions can improve specific objective measures and increase knowledge, yet they frequently fail to yield meaningful, sustained changes in subjective outcomes without direct human interaction.^19 22^ Digital health researchers emphasise that blended approaches - combining digital tools with professional support - may enhance effectiveness, balancing scalability with the relational support that underpins more robust behaviour change.^19 22^ Successful delivery of mental health assessment and intervention through paediatric epilepsy specialist nurses and graduate psychologists supports the feasibility and scalability of such a blended approach.^23^

We may ask why the evidence-based behavioural sleep interventions at the core of COSI were not effective for sleep problems in childhood epilepsy yet have proven effective in other paediatric populations, including those with neurodevelopmental disorders, and using varied delivery methods. There may be unique factors in the childhood epilepsy population - such as the bidirectional link between sleep and seizures, the confounding role of antiseizure medications, heightened parental anxiety about sudden unexpected death in epilepsy (SUDEP)^24^ - that might have impacted the intervention effect.

Parents whose children experienced either longstanding or mild sleep issues might have engaged less or exhibited ceiling or floor effects, highlighting the importance of patient selection. However, the severity of sleep problems in the trial was similar to that in other neurodevelopmental disorder research samples, suggesting low symptom severity is an unlikely explanation.^25^

Despite a lack of clinically significant change in the primary endpoint of subjective sleep problems, there were important findings in secondary outcomes. First, actigraphy-measured sleep-onset latency decreased by 16.35 minutes in the intervention group - a notable improvement given that a 10-minute difference is often considered clinically significant.^20^ Sleep-onset latency is frequently cited as a problem in CWE and reported as a major concern for parents.^24^ Interestingly, there seems to be little or no correlation between polysomnography or actigraphy measures with any subscale of the CSHQ.^26^ Second, parents’ knowledge of child sleep improved significantly in the SC + COSI group. Previous studies show that increased parental sleep knowledge correlates with healthier child sleep habits, and such knowledge can be enhanced by both educational and behavioural interventions.^27^ However, knowledge alone may not suffice for behaviour change.

**Limitations**. There were several limitations of the trial. First, the originally intended sample size was not attained owing to clinical disruption during and after the COVID-19 pandemic. Loss of statistical power may theoretically have contributed to inability to demonstrate an effect. Second, while the trial correctly employed an intention-to-treat analysis, (non)adherence measures such as the relative risk of dropping out or of stopping the use of an application, as well as a “usage half-life”, and models reporting demographic and other factors predicting usage discontinuation might have been helpful to modify further iterations of this digital intervention.^19^ Third, there were incomplete data which increased the uncertainty of health economic outcomes and decreased the statistical power of secondary outcomes because the burden of online trial participation may have been excessive for some participants. Fourth, although we used a shorter unvalidated version of a common sleep problems questionnaire, calculations of Cronbach’s alpha and sub-analyses of items common to both short and long instruments (Appendix 1) revealed no significant differences between treatment groups. Last, the lack of blinding could have introduced bias in reporting outcomes although this might be expected to be in favour of the intervention, which was not apparent.

In summary, this web-based intervention for parents of children with epilepsy did not lead to improvements in parent-reported child sleep problems and was not cost-effective. Nevertheless, significant benefits were observed in objective sleep onset latency and in parents’ knowledge of child sleep – benefits seen mostly in a sub-population of COSI engagers. However, the trial faced real-life challenges common in eHealth including poor uptake, high attrition, and uncertainty regarding how fully participants engaged with digital content. The trial findings suggest that sleep problems in childhood epilepsy cannot be tackled across the NHS using online parent-directed resources and standard care alone. Since specialist sleep psychologists will remain scarce in the foreseeable future, training of other non-sleep staff already working with these families, such as epilepsy specialist nurses, in combination with digital tools, may provide an approach to enhance effectiveness - blending technical guidance with motivational support. The evaluation of such a model could be a focus for future research.

## Data Availability

All data produced in the present work are contained in the manuscript and its included appendices

## Acknowledgements

The authors would like to thank all the families who participated in the trial, site principal investigators and research staff, previous trial and programme managers, graduate researchers, families who were members of our Patient and Public Involvement Research Advisory Panel, the independent data monitoring and safety monitoring committee, and the programme steering committee.

## Contributions

DKP, PG conceived the idea for the study and acted as joint Chief Investigators. Together with LB, BC, DAH, CM, CTS, LW they submitted the successful grant application. TC and LSE led on trial management. LW was the workpackage lead for development of the COSI intervention assisted by GC. CM led the workpackage on development of core outcome set. BC led the qualitative workpackage assisted by HS. PG and DKP are co-inventors of the SleepSuite memory consolidation iPad game. PG produced the first draft of the manuscript. AA did the statistical analyses for the manuscript supervised by CTS. LB was the patient and public involvement lead. SL was the programme manager. DAH led the economic analysis and WH performed the economic analyses. All authors had full access to all the data in the study (with the exception of the patient-level PLICS data, which DAH and WASH alone had access to, and the qualitative data which BC and HS alone had access to). All authors had final responsibility for the decision to submit for publication, approved the final version of the manuscript and agree to be accountable for all aspects of the work in ensuring that questions related to the accuracy or integrity of any part of the work are appropriately investigated and resolved.

## Data sharing

Data are not publicly available. In line with the ethical approval of the study and to protect the identity of research participants, qualitative data will not be available for sharing.

All requests for data will be reviewed by the CASTLE data access committee, to verify whether the request is subject to any intellectual property or confidentiality obligations. Requests for access to the participant-level data from this study can be submitted via email to the corresponding author with detailed proposals for approval. A signed data access agreement with the CASTLE team is required before accessing shared data.

King’s College London will consider all data sharing requests and review the requirements both from a practical logistical perspective and in consideration and compliance with GDPR and current Data Protection legislation. All data sharing arrangements will be discussed with our collaborators and with our Information Governance Data Controller utilising a Data Protection Impact Assessment and Data Sharing Agreement(s) to be put in place with the Requester. We will also request acknowledgement of the Terms of the original funding from NIHR.

## Appendices

## Appendix 1. Sensitivity analysis of CSHQ-22

### Baseline CSHQ-22 individual item score (N=85)

The baseline individual CSHQ-22 items summary measures for all the 85 participants randomised are given below. This table includes both the 5-point scoring and the 3-point scoring for the CSHQ items at baseline split by treatment arm. Items marked in blue are the 17 items that are common between the 22-item and 33-item questionnaire.

**Table 1.**
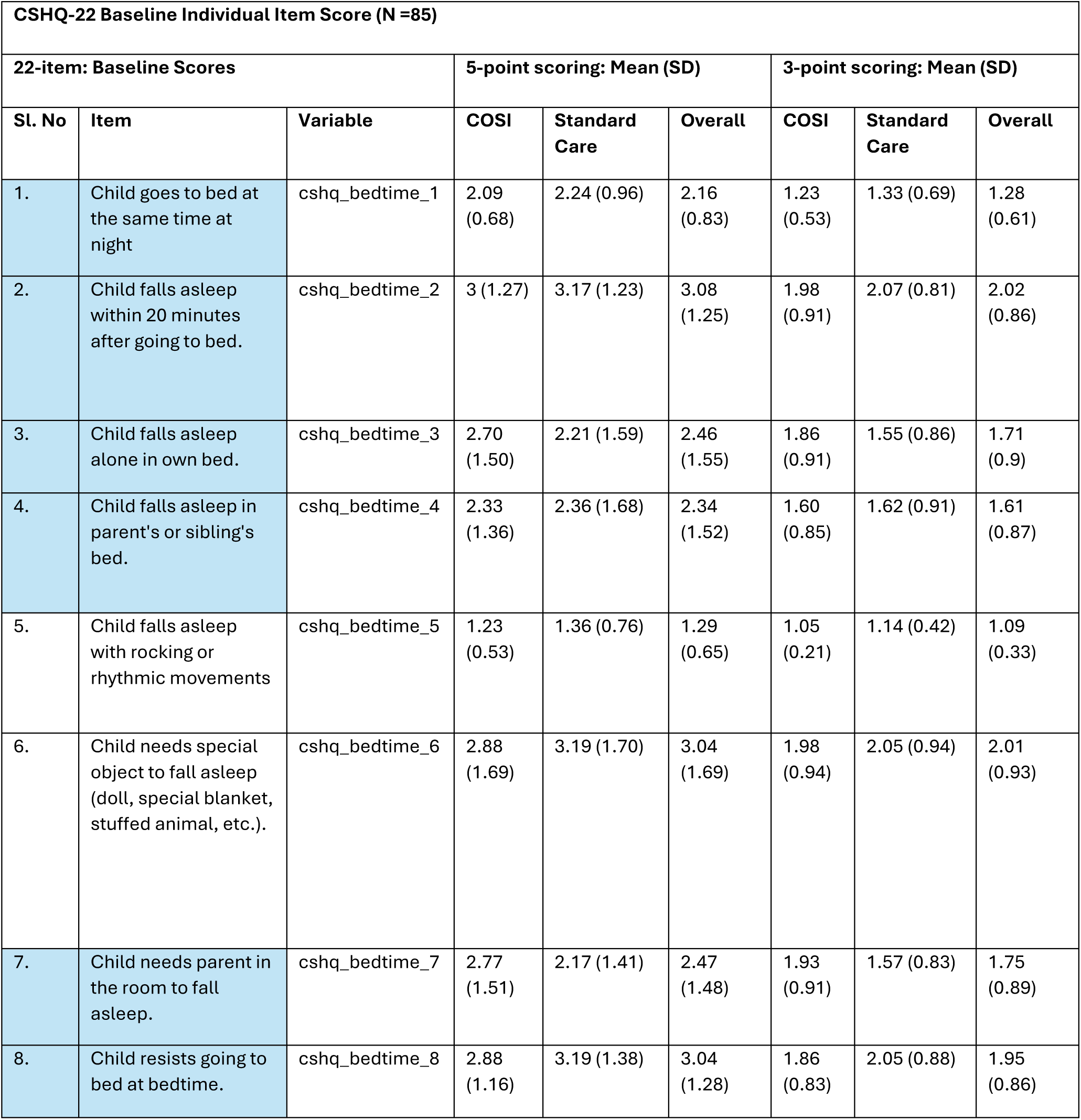
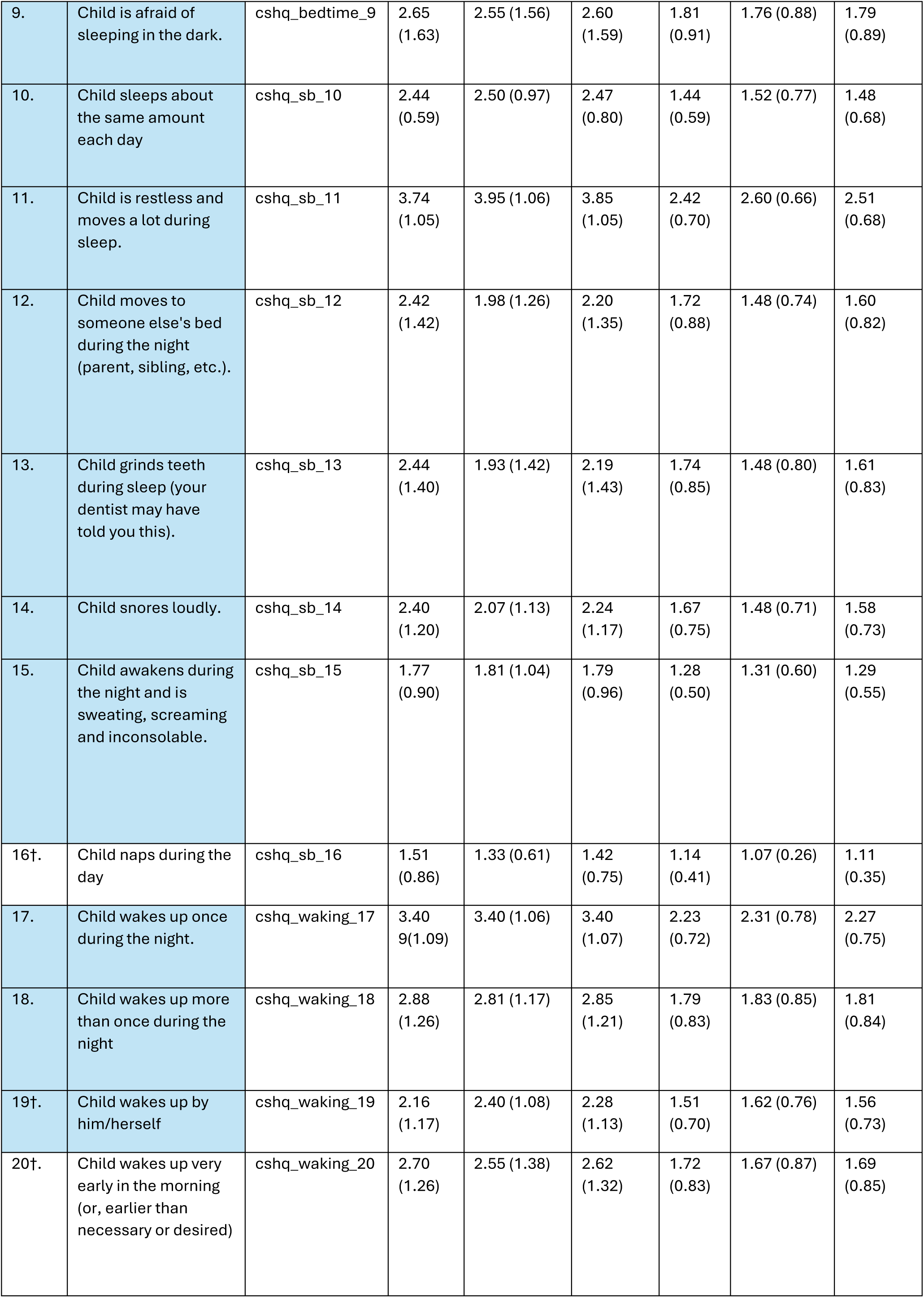
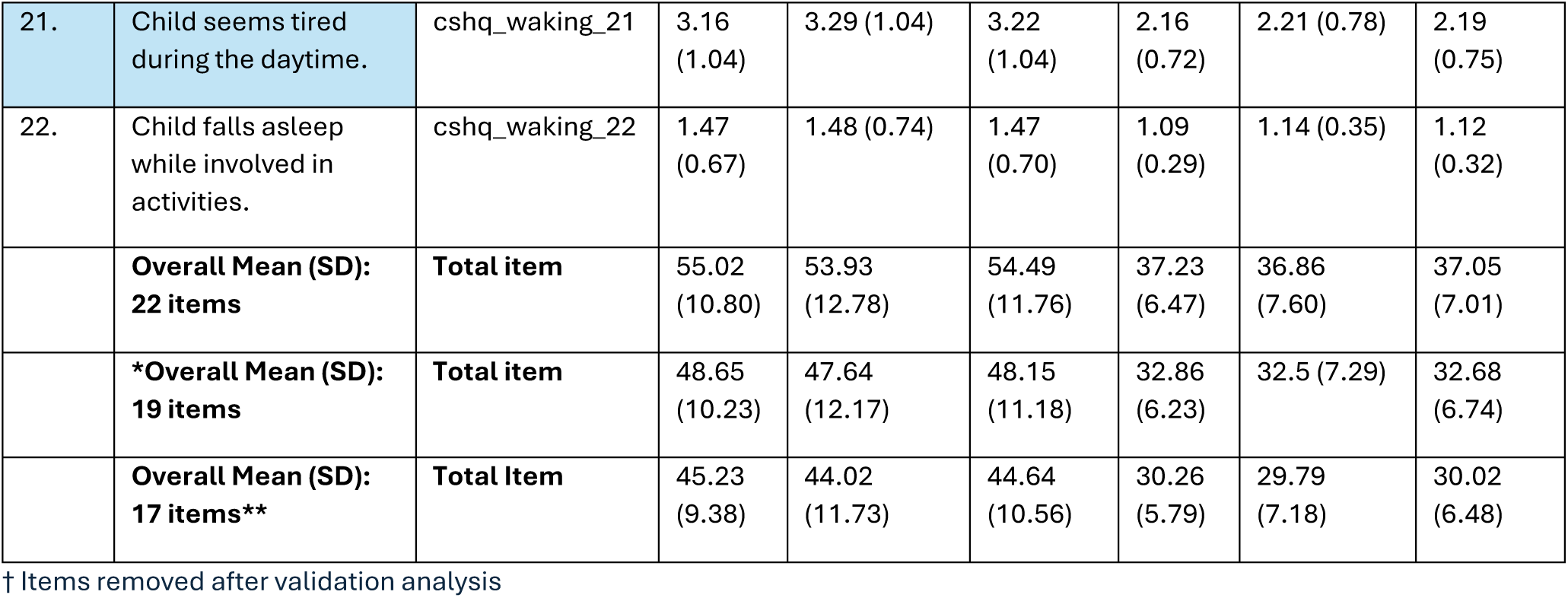
CSHQ-22 baseline Individual item score (N=85). 22 items of the CSHQ-22. *Three items are excluded after internal consistency analysis (16,19,20) **17 items overlap with the CSHQ-33 version (highlighted in blue).

### Baseline CSHQ-22 individual item score (N=72)

The baseline summary measures for each individual item on the CSHQ-22, based on the data from all 72 participants who completed the 3-month CSHQ-22 questionnaire and are within the 3-month follow-up window (included in primary analysis set), are given below. This table includes both the 5-point scoring and the 3-point scoring for the CSHQ-22 items at baseline split by treatment arm. Items marked in blue are the 17 items that are common between the 22-item and 33-item questionnaire.

**Table 2.**
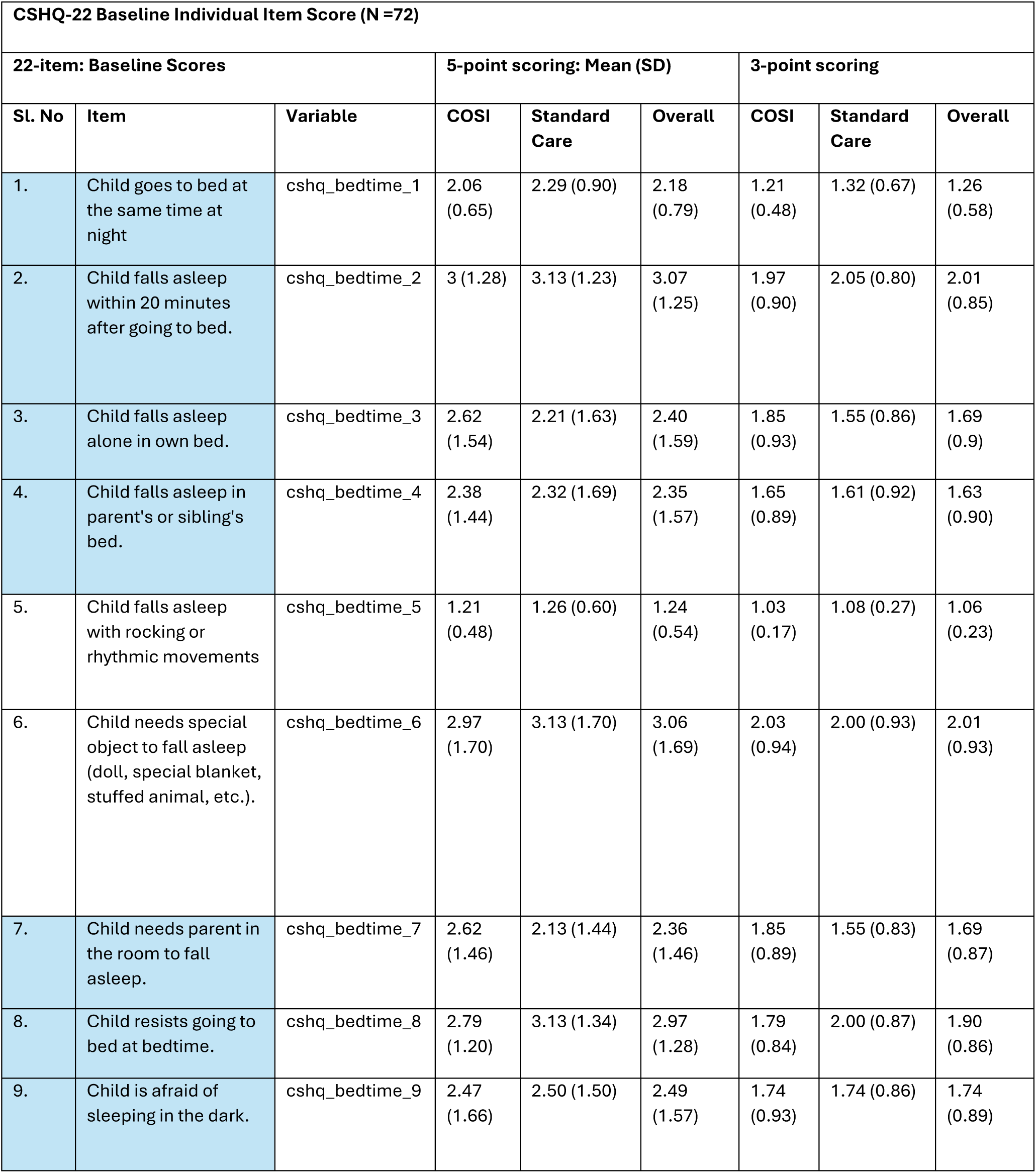
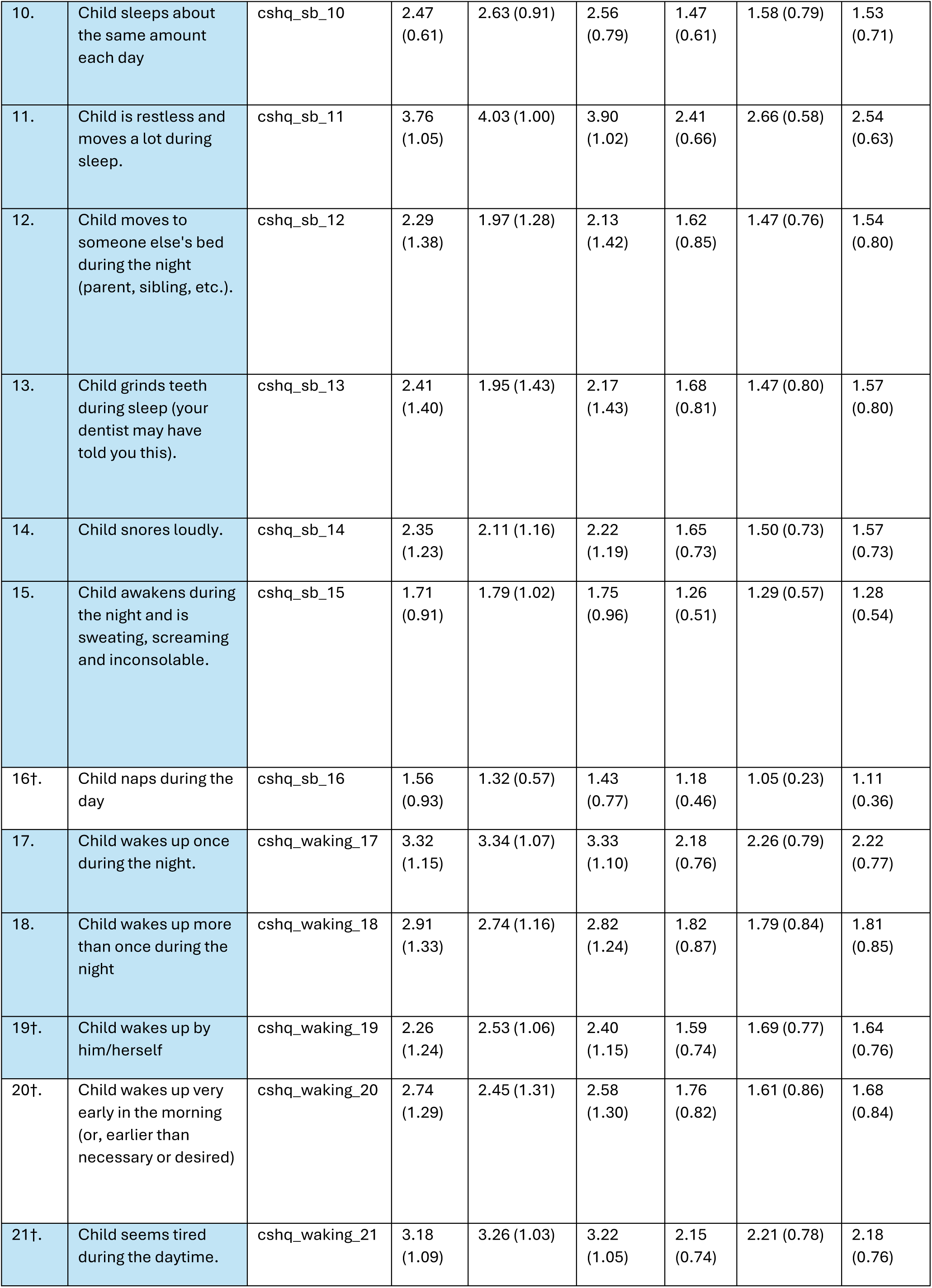
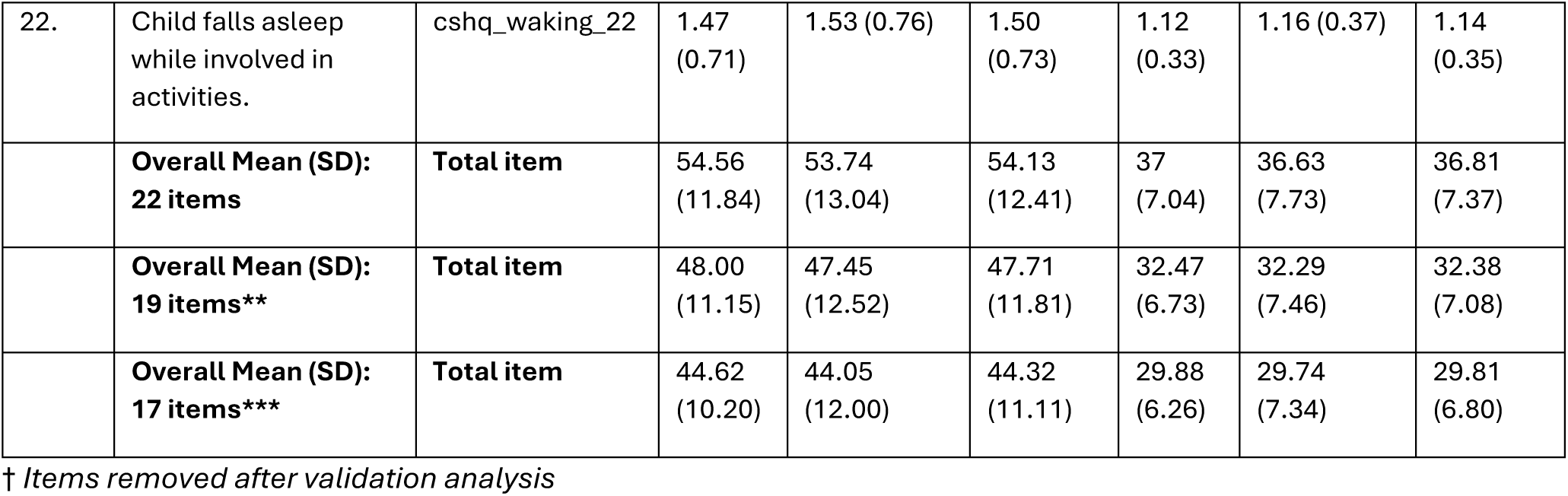
CSHQ-22 Baseline individual score.

### Cronbach Alpha Coefficient: 22-item scale

The overall standardised Cronbach coefficient alpha is 0.82 and the overall raw Cronbach coefficient alpha is 0.82 for the full 22-item questionnaire. For the 22-item questionnaire the overall Cronbach’s alpha was examined as a measure of internal consistency. The item-total corrected correlations and Cronbach’s alpha excluding each item for all the 22-items were calculated. Items that showed an increase in Cronbach’s alpha (raw variable) on deletion and very low Corrected Item-Total Correlation (<.20) was omitted from the total score.

**Table 3.** Cronbach’s coefficient alpha with deleted variables.

| Cronbach Coefficient Alpha with deleted variables |  |  |  |  |
| --- | --- | --- | --- | --- |
|  | Raw variable |  | Standardised variable |  |
| Deleted variable | Correlation with total | Alpha | Correlation with total | Alpha |
| csHQ_bedtime_1 | 0.203925 | 0.824169 | 0.200189 | 0.825431 |
| csHQ_bedtime_2 | 0.480833 | 0.812793 | 0.471867 | 0.813036 |
| csHQ_bedtime_3 | 0.452172 | 0.81425 | 0.447818 | 0.81416 |
| csHQ_bedtime_4 | 0.474947 | 0.813569 | 0.499755 | 0.811726 |
| csHQ_bedtime_5 | 0.225305 | 0.82365 | 0.237776 | 0.823755 |
| csHQ_bedtime_6 | 0.375394 | 0.820102 | 0.370797 | 0.817724 |
| csHQ_bedtime_7 | 0.492023 | 0.812034 | 0.480946 | 0.81261 |
| csHQ_bedtime_8 | 0.660293 | 0.803881 | 0.65918 | 0.804102 |
| csHQ_bedtime_9 | 0.398256 | 0.817669 | 0.393655 | 0.816672 |
| csHQ_sb_10 | 0.298688 | 0.821377 | 0.292986 | 0.821271 |
| csHQ_sb_11 | 0.619424 | 0.806709 | 0.612374 | 0.806365 |
| csHQ_sb_12 | 0.466044 | 0.813508 | 0.463013 | 0.81345 |
| csHQ_sb_13 | 0.419964 | 0.815959 | 0.387304 | 0.816965 |
| csHQ_sb_14 | 0.38906 | 0.81747 | 0.392279 | 0.816735 |
| csHQ_sb_15 | 0.557153 | 0.812445 | 0.554199 | 0.809149 |
| csHQ_sb_16 | 0.185684 | 0.824592 | 0.20472 | 0.82523 |
| csHQ_waking_17 | 0.30128 | 0.821376 | 0.288878 | 0.821457 |
| csHQ_waking_18 | 0.626273 | 0.805822 | 0.621565 | 0.805922 |
| csHQ_morning_19 | -0.107881 | 0.838568 | -0.088668 | 0.837907 |
| csHQ_morning_20 | 0.181425 | 0.827927 | 0.160202 | 0.827201 |
| csHQ_morning_21 | 0.362498 | 0.818654 | 0.385051 | 0.817068 |
| csHQ_morning_22 | 0.345504 | 0.820317 | 0.364388 | 0.818018 |

**Sensitivity Analysis: Total CSHQ score at 3 months (±1 month) from a linear mixed effects regression model calculated after removing item 19 for the 3-point scored 22-item questionnaire**

The correlation of Item 19 with the total score was -0.11 for the raw variable and -0.09 for the standardised variable.

**Table 4.** Total CSHQ score at 3 months (±1 month) from a linear mixed effects regression model calculated after removing item 19.

| Treatment group | N | Mean (SD) CSHQ score at baseline | Mean (SD) CSHQ score at 3 months | Point Estimate (SE) | 95% CI | p-value |
| --- | --- | --- | --- | --- | --- | --- |
| COSI | 34 | 35.41 (7.15) | 35.26 (7.70) | 1.98 (0.96) | 0.05, 3.90 | 0.04 |
| Standard Care | 38 | 34.95 (7.54) | 32.84 (6.96) |  |  |  |
| Total | 72 | 35.17 (7.31) | 33.99 (7.37) |  |  |  |

**Sensitivity Analysis: Total CSHQ score at 3 months (±1 month) from a linear mixed effects regression model calculated after removing item 16, 19 and 20 for the 3-point scored 22-item questionnaire** Item 16 had a correlation of 0.19 with the total score for the raw variable and Item 20 had a correlation of 0.18 with the total score for the raw variable.

**Table 5.** Total CSHQ score at 3 months (±1 month) from a linear mixed effects regression model calculated after removing items 16,19 and 20.

| Treatment group | N | Mean (SD) CSHQ score at baseline | Mean (SD) CSHQ score at 3 months | Point Estimate (SE) | 95% CI | p-value |
| --- | --- | --- | --- | --- | --- | --- |
| COSI | 34 | 32.47 (6.73) | 32.47 (7.30) | 1.81 (0.92) | -0.03, 3.66 | 0.05 |
| Standard Care | 38 | 32.29 (7.46) | 30.21 (6.77) |  |  |  |
| Total | 72 | 32.38 (7.08) | 31.17 (7.05) |  |  |  |

**Sensitivity Analysis: Total CSHQ score at 3 months (±1 month) from a linear mixed effects regression model calculated after removing items 19 and 20 for the 3-point scored 22-item questionnaire**

Item 19 had a correlation of -0.09 with the total score for the standardised variable, while item 20 had a correlation of 0.16 with the total score for the standardised variable.

**Table 6.** Total CSHQ score at 3 months (±1 month) from a linear mixed effects regression model calculated after removing items 19 and 20.

| Treatment group | N | Mean (SD) CSHQ score at baseline | Mean (SD) CSHQ score at 3 months | Point Estimate (SE) | 95% CI | p-value |
| --- | --- | --- | --- | --- | --- | --- |
| COSI | 34 | 33.65 (6.80) | 33.44 (7.37) | 1.84 (0.93) | -0.03, 3.72 | 0.05 |
| Standard Care | 38 | 33.34 (7.52) | 31.29 (6.77) |  |  |  |
| Total | 72 | 33.49 (7.14) | 32.31 (7.09) |  |  |  |

**Sensitivity Analysis: Total CSHQ score at 3 months (±1 month) using a linear mixed effects regression model for the 5-point scale incorporating all the 22-items**

**Table 7:** Total CSHQ score at 3 months (±1 month) from a linear mixed effects regression model calculated for the 5-point scale including all the 22-items.

| Treatment group | N | Sleep medication: Yes (%) | Mean (SD) CSHQ score at baseline | Mean (SD) CSHQ score at 3 months | Point Estimate (SE) | 95% CI | p-value |
| --- | --- | --- | --- | --- | --- | --- | --- |
| COSI | 34 | 8 (23.53%) | 54.56 (11.84) | 54.74 (13.03) | 2.95 (1.53) | -0.13, 6.02 | 0.06 |
| Standard Care | 38 | 8 (22.05%) | 53.74 (13.04) | 50.95 (12.39) |  |  |  |
| Total | 72 | 16 (22.22%) | 54.13 (12.41) | 52.74 (12.75) |  |  |  |

**Sensitivity Analysis: Total CSHQ score at 6 months (±1 month) using a linear mixed effects regression model for the 5-point scale incorporating all the 22-items**

**Table 8.** : Total CSHQ score at 6 months (±1 month) from a linear mixed effects regression model calculated for the 5-point scale including all the 22-items.

| Treatment group | N | Mean (SD) CSHQ score at baseline | Mean (SD) CSHQ score at 6 months | Point Estimate (SE) | 95% CI | p-value |
| --- | --- | --- | --- | --- | --- | --- |
| COSI | 20 | 52.80 (10.09) | 53.45 (9.97) | 3.33 (2.25) | -1.39, 8.04 | 0.16 |
| Standard Care | 20 | 53.55 (14.52) | 50.80 (13.38) |  |  |  |
| Total | 40 | 53.18 (12.35) | 52.13 (11.72) |  |  |  |

**Sensitivity Analysis: Total CSHQ score at >=4 months from a linear mixed effects regression model for the 5-point scale incorporating all the 22-items**

**Table 9:** Total CSHQ score at ≥4 months from a linear mixed effects regression model calculated for the 5-point scale including all the 22-items.

| Treatment group | N | Mean (SD) CSHQ score at baseline | Mean (SD) CSHQ score at $\geq 4$ | Point Estimate (SE) | 95% CI | p-value |
| --- | --- | --- | --- | --- | --- | --- |
| COSI | 26 | 52.19 (10.32) | 51.42 (10.10) | 1.26 (1.89) | -2.60, 5.13 | 0.51 |
| Standard Care | 24 | 53.25 (13.74) | 51.0 (12.54) |  |  |  |
| Total | 50 | 52.70 (11.97) | 51.22 (11.22) |  |  |  |

**Sensitivity Analysis: Total CSHQ score at 3 months (±1 month) from a linear mixed effects regression model calculated after removing item 19 for the 5-point scored 22-item questionnaire**

**Table 10.** : Total CSHQ score at 3 months (±1 month) from a linear mixed effects regression model calculated after removing item 19 for the 5-point scale 22-item questionnaire.

| Treatment group | N | Mean (SD) CSHQ score at baseline | Mean (SD) CSHQ score at 3 months | Point Estimate (SE) | 95% CI | p-value |
| --- | --- | --- | --- | --- | --- | --- |
| COSI | 34 | 52.29 (11.91) | 52.50 (13.30) | **3.06 (1.53)** | **-0.02, 6.14** | **0.05** |
| Standard Care | 38 | 51.21 (12.73) | 48.39 (12.24) |  |  |  |
| Total | 72 | 51.72 (12.28) | 50.33 (12.83) |  |  |  |

**Sensitivity Analysis: Total CSHQ score at 3 months (±1 month) from a linear mixed effects regression model calculated after removing items 19 and 20 for the 5-point scored 22-item questionnaire**

**Table 11:** Total CSHQ score at 3 months (±1 month) from a linear mixed effects regression model calculated after removing item 19, 20 for the 5-point scale 22-item questionnaire.

| Treatment group | N | Mean (SD) CSHQ score at baseline | Mean (SD) CSHQ score at 3 months | Point Estimate (SE) | 95% CI | p-value |
| --- | --- | --- | --- | --- | --- | --- |
| COSI | 34 | 49.56 (11.37) | 49.76 (12.75) | **2.92 (1.50)** | **-0.08, 5.93** | **0.06** |
| Standard Care | 38 | 48.76 (12.68) | 46.05 (11.97) |  |  |  |
| Total | 72 | 49.14 (12.0) | 47.81 (12.40) |  |  |  |

**Sensitivity Analysis: Total CSHQ score at 3 months (±1 month) using an unadjusted linear mixed effects regression model for the 5-point scale after removing items 16, 19 and 20**

**Table 12:**
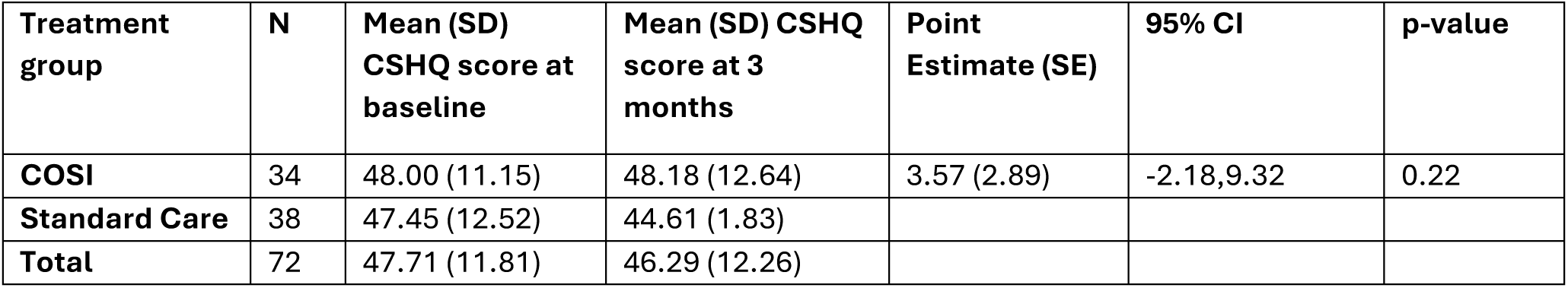
Total CSHQ score at 3 months (±1 month) from an unadjusted linear mixed effects regression model calculated after removing item 16,19, 20 for the 5-point scale 22-item questionnaire.

| Treatment group | N | Mean (SD) CSHQ score at baseline | Mean (SD) CSHQ score at 3 months | Point Estimate (SE) | 95% CI | p-value |
| --- | --- | --- | --- | --- | --- | --- |
| **COSI** | 34 | 48.00 (11.15) | 48.18 (12.64) | 3.57 (2.89) | -2.18,9.32 | 0.22 |
| **Standard Care** | 38 | 47.45 (12.52) | 44.61 (1.83) |  |  |  |
| **Total** | 72 | 47.71 (11.81) | 46.29 (12.26) |  |  |  |

**Baseline CSHQ 33 item scale with imputed individual item scores (N=85)**

This table includes the mean (sd) score for each item on the CSHQ-33 item scale using the 3-point scoring at baseline. Missing items (data not collected) have been imputed based on the mean score for that domain.

**Table 13:** Baseline CSHQ Individual Item score (N=85).

| 33-item: Baseline Scores |  |  | 3-point scoring |  |  |
| --- | --- | --- | --- | --- | --- |
| Sl. No | Item | Variable | COSI | Standard Care | Overall |
| 1. | Goes to bed at same time | csdq_resistance_1 | 1.23 (0.53) | 1.33 (0.69) | 1.28 (0.61) |
| 2. | Falls asleep in 20 minutes | csdq_sleeponset_2 | 1.98 (0.91) | 2.07 (0.81) | 2.02 (0.86) |
| 3. | Falls asleep in own bed | csdq_resistance_3 | 1.86 (0.91) | 1.55 (0.86) | 1.71 (0.9) |
| 4. | Falls asleep in other's bed | csdq_resistance_4 | 1.60 (0.85) | 1.62 (0.91) | 1.61 (0.87) |
| 5. | Needs parent in room to sleep | csdq_resistance_5 | 1.93 (0.91) | 1.57 (0.83) | 1.75 (0.23) |
| 6. | Struggles at bedtime | csdq_resistance_6 | 1.86 (0.83) | 2.05 (0.88) | 1.95 (0.86) |
| 7. | Afraid of sleeping in the dark | csdq_anxiety_7 | 1.81 (0.91) | 1.76 (0.88) | 1.79 (0.89) |
| 8. | Afraid of sleeping alone | csdq_resistance_8 | 1.70 (0.57) | 1.62 (0.64) | 1.66 (0.60) |
| 9. | Sleeps too little | csdq_sleepduration_9 | 1.44 (0.59) | 1.52 (0.77) | 1.48 (0.68) |
| 10. | Sleeps the right amount | csdq_sleepduration_10 | 1.44 (0.59) | 1.52 (0.77) | 1.48 (0.68) |
| 11. | Sleeps same amount each day | csdq_sleepduration_11 | 1.44 (0.59) | 1.52 (0.77) | 1.48 (0.68) |
| 12. | Wets the bed at night | csHQ_parasomnia_12 | 1.81 (0.46) | 1.79 (0.47) | 1.80 (0.46) |
| 13. | Talks during sleep | csHQ_parasomnia_13 | 1.81 (0.46) | 1.79 (0.47) | 1.80 (0.46) |
| 14. | Restless and moves a lot | csHQ_parasomnia_14 | 2.42 (0.70) | 2.60 (0.66) | 2.51 (0.68) |
| 15. | Sleepwalks | csHQ_parasomnia_15 | 1.81 (0.46) | 1.79 (0.47) | 1.80 (0.46) |
| 16. | Moves to other's bed in night | csHQ_nightwalking_16 | 1.72 (0.88) | 1.48 (0.74) | 1.60 (0.82) |
| 17. | Grinds teeth during sleep | csHQ_parasomnia_17 | 1.74 (0.85) | 1.48 (0.80) | 1.61 (0.83) |
| 18. | Snores loudly | csHQ_sleepdisorder_18 | 1.67 (0.75) | 1.48 (0.71) | 1.58 (0.73) |
| 19. | Stops breathing | csHQ_sleepdisorder_19 | 1.67 (0.75) | 1.48 (0.71) | 1.58 (0.73) |
| 20. | Snorts and gasps | csHQ_sleepdisorder_20 | 1.67 (0.75) | 1.48 (0.71) | 1.58 (0.73) |
| 21. | Trouble sleeping away | csHQ_anxiety_21 | 1.81 (0.61) | 1.65 (0.63) | 1.73 (0.62) |
| 22. | Awakens screaming, sweating | csHQ_parasomnia_22 | 1.28 (0.50) | 1.31 (0.60) | 1.29 (0.55) |
| 23. | Alarmed by scary dream | csHQ_parasomnia_23 | 1.81 (0.46) | 1.79 (0.46) | 1.80 (0.46) |
| 24. | Awakens once during night | csHQ_nightwalking_24 | 2.23 (0.72) | 2.31 (0.78) | 2.27 (0.75) |
| 25. | Awakens more than once | csHQ_nightwalking_25 | 1.79 (0.83) | 1.83 (0.85) | 1.81 (0.84) |
| 26. | Wakes by himself | csHQ_daytime_26 | 1.51 (0.70) | 1.62 (0.76) | 1.56 (0.73) |
| 27 | Wakes up in a negative mood | csHQ_daytime_27 | 1.84 (0.48) | 1.92 (0.55) | 1.88 (0.52) |
| 28 | Others wake child | csHQ_daytime_28 | 1.84 (0.48) | 1.92 (0.55) | 1.88 (0.52) |
| 29 | Hard time getting out of bed | csHQ_daytime_29 | 1.84 (0.48) | 1.92 (0.55) | 1.88 (0.52) |
| 30 | Takes long time to be alert | csHQ_daytime_30 | 1.84 (0.48) | 1.92 (0.55) | 1.88 (0.52) |
| 31 | Seems tired | csHQ_daytime_31 | 2.16 (0.72) | 2.21 (0.78) | 2.19 (0.75) |
| 32 | Watching TV | csHQ_daytime_32 | 1.84 (0.48) | 1.92 (0.55) | 1.88 (0.52) |
| 33 | Riding in car | csHQ_daytime_33 | 1.84 (0.48) | 1.92 (0.55) | 1.88 (0.52) |
|  | **Overall Mean (SD): 33 items** | **Total item** | 58.28 (9.78) | 57.73 (13.33) | 58.01 (11.61) |

**Figure 1.**
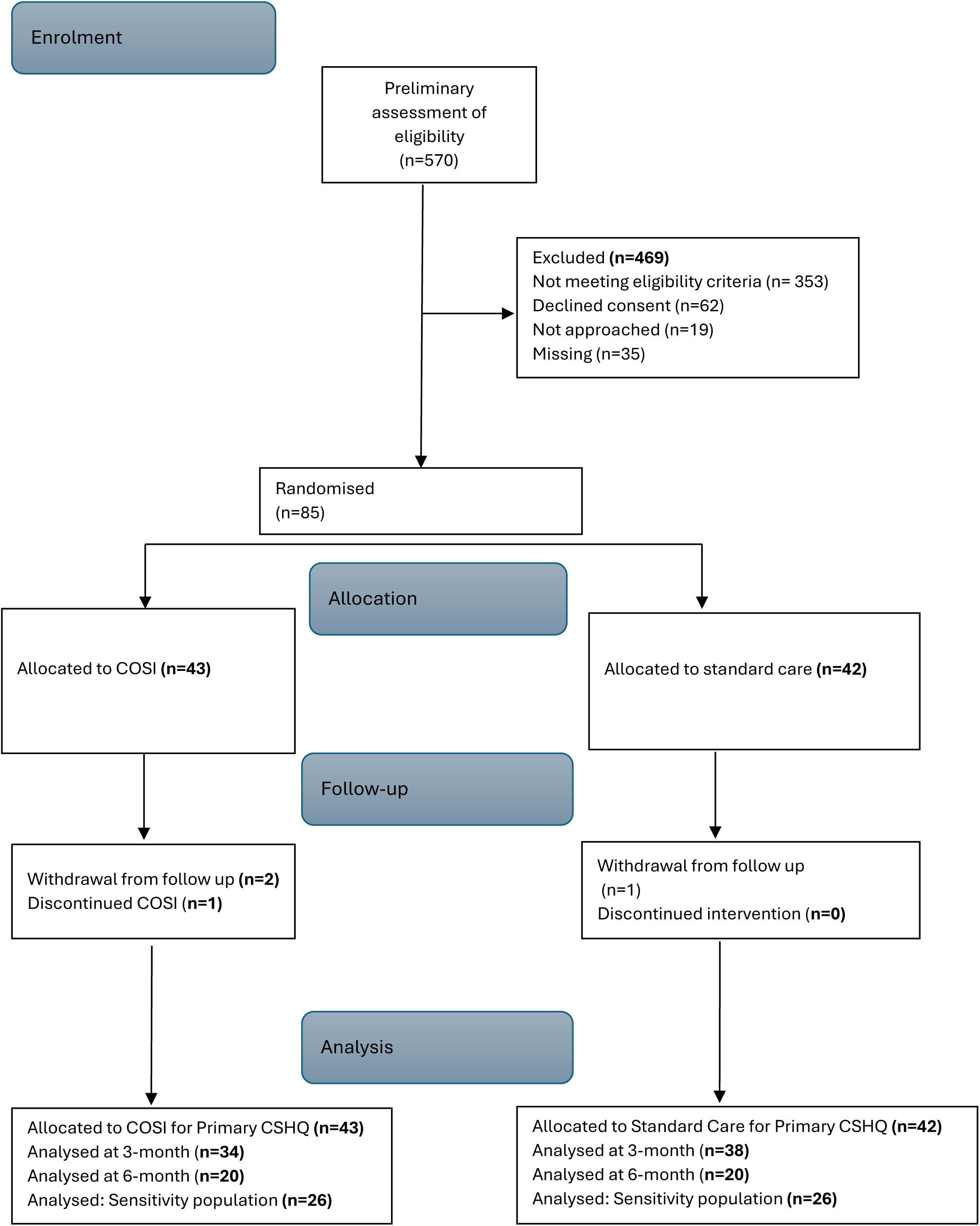
Participant Flow Diagram.

**Figure 2.**
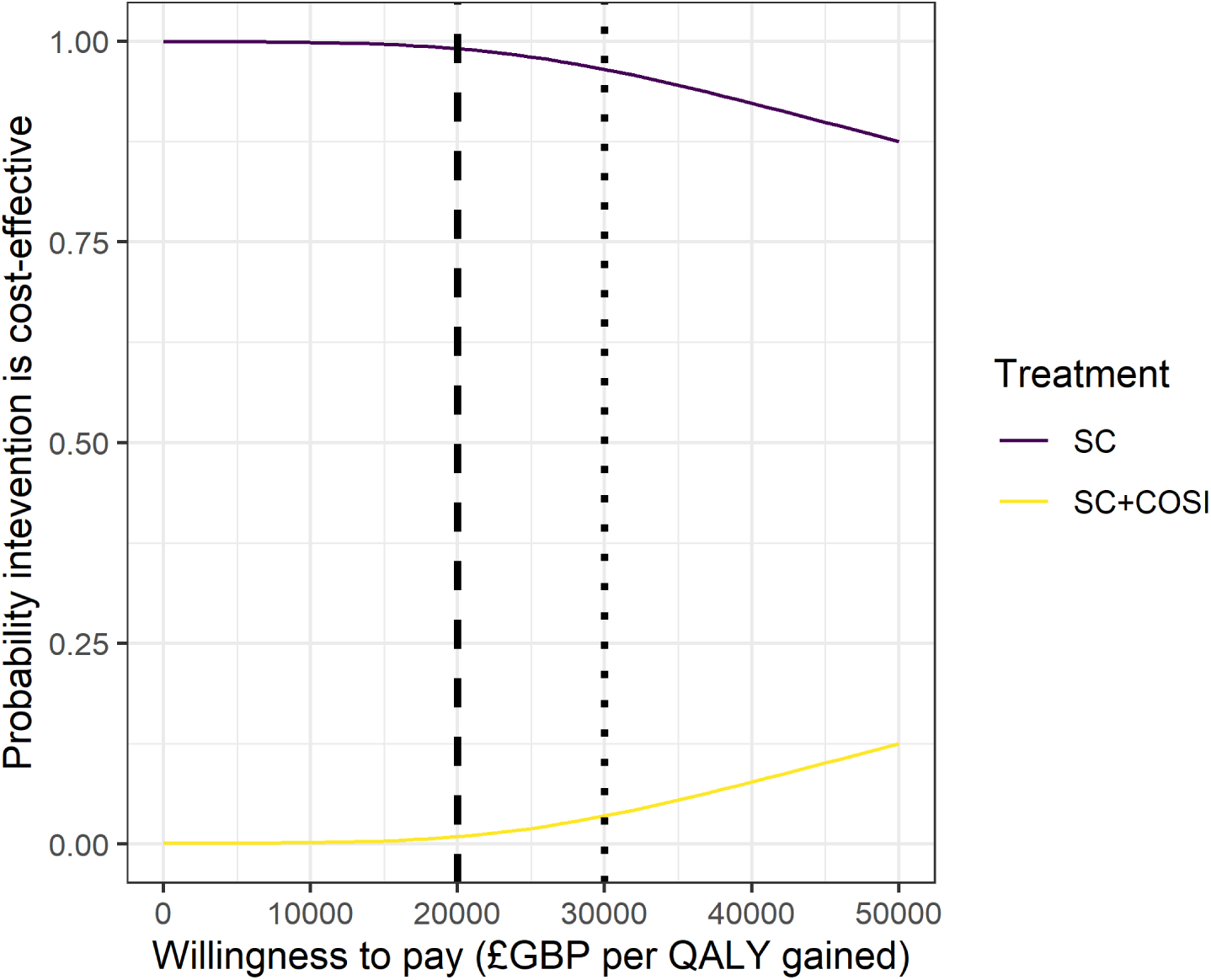
Cost-effectiveness acceptability curve for the base case health economic analysis. Vertical lines represent willingness to pay thresholds of £20,000 per QALY (dashed) and £30,000 per QALY (dotted).

## Appendix 2. Health Economic Analysis

## 1. Abstract

Childhood epilepsy is a common neurological condition that can affect the quality of life of the children with epilepsy and their parents, with sleep problems being one of the difficulties faced. The CASTLE Online Sleep Intervention (COSI) has been designed to improve the sleep of children with epilepsy. **Methods:** To evaluate the cost-effectiveness, expressed as the incremental cost-effectiveness ratio (ICER), of supplementing standard care (SC) with COSI compared to SC alone, we performed a cost-utility analysis from a national health service (NHS) and personal social services (PSS) cost-perspective, with quality adjusted life years (QALYs) generated from utility data collected using the Child Health Utility 9 Dimension Index (CHU9D). Resource use was measured through a complimentary approach using patient level costing information system (PLICS) data for secondary care and resource use questionnaires for primary and community care. Incremental costs and QALYs were estimated using Bayesian multivariate regression models applied to multiply imputed datasets, to account for missing data, this also provided estimates of uncertainty. **Results:** SC + COSI was associated with higher costs (£1,232 [95% CrI £ 535; £2,183]) and a difference in QALYs of 0.00 [ -0.03; 0.04], the resulting ICER was £433,167 per QALY, and probabilities of being cost-effective at the £20,000 and £30,000 per QALY gained thresholds was 0.01 and 0.04 respectively. The results of a range of sensitivity analyses, which challenged key assumptions, all supported the primary analysis, suggesting that SC + COSI would not be a cost-effective use of NHS + PSS resources at willingness to pay thresholds usually considered by NICE.

## 2. Introduction

This is a report of the health economic analysis of the CASTLE Sleep-E trial comparing the cost-effectiveness of standard care (SC) to SC supplemented with the CASTLE Online Sleep Intervention (COSI) for children with epilepsy.

### 2.1 Background and objectives

Epilepsy is one of the most common long-term neurological conditions worldwide, with incidence peaking in childhood (5–9 years) and again in later life (over 80 years).^1^ In children aged 5 to under 13, it accounts for over 2.6 million lost disability-adjusted life years annually—1.8% of the global burden of disease.^2^ Sleep problems are a key concern for children with epilepsy and their parents, as those children often fear death during seizures and parents often co-sleep or constantly monitor them.^3,4^ Despite this concern, sleep difficulties are frequently overlooked.^5,6^

Recent evidence shows parent-based behavioural sleep interventions are effective for both typically-developing children and those with neurological and neuro-developmental disorders.^6^ However, few trials have focused on epilepsy, and potential harms are seldom addressed.^7^ Where harms are reported (in adults), they are usually mild, such as transient fatigue.^8^

Although the benefits of sleep interventions—improving seizure control, learning, and parental well-being—likely outweigh the risks^5,6^ the cost-effectiveness of sleep interventions in public health systems remains untested.

Aims of the economic evaluation:

1. To compare the cost-effectiveness of SC and SC + COSI in children (4-12 years old) with epilepsy from the perspective of a National Health Service (NHS) and Personal Social Services (PSS),
2. To compare health outcomes, measured in quality-adjusted life years (QALYs), and costs between the treatments.

## 3. Methods

### 3.1 Overview

We conducted the primary economic evaluation from an NHS and PSS perspective in England to estimate whether SC + COSI represents a cost-effective addition to current SC alone for children with epilepsy. This evaluation was based on a within-trial analysis, using individual, patient-level data from trial sites and CASTLE Sleep-E trial, comparing the costs incurred and utility experienced within each of the treatment groups over the first six (± one) months post-randomisation.

The primary outcome was the incremental cost-effectiveness ratio (ICER), expressed as costs (£GBP) per quality-adjusted life year (QALY) gained, calculated according to the Health Economics Analysis Plan (HEAP) (*Section 10.2*). The health economic analysis was carried out in RStudio (see *Package citations* for version and package information), and is reported according to the Consolidated Health Economic Evaluation Reporting Standards 2022 (CHEERS 2022) statement^9^ (*Table 16*).

### 3.2 Data sources

Data were collected from routine healthcare records, from participants, and their primary carers as part of the trial during or shortly after site visits.

#### 3.2.1 Visit Timing

Participants and their primary carer were asked to attend four visits:

1. Baseline,
2. Randomisation, to be within 8 weeks of the baseline visit,
3. Follow-up 1, to be 3 ± 1 months post-randomisation,
4. Follow-up 2, to be 6 ± 1 months post-randomisation or by the end of the trial (2024-01-31), whichever was sooner.

Consequently, some participants may only have attended one follow-up visit because there was insufficient time for the second follow-up visit before the trial closed. *Table 1* summarises information about the timing of questionnaire completion and *Figure 1* shows the visit timing for each participant.

### Note

- Baseline and randomisation were asynchronous (mean difference = 33.8 days, SD = 17.75)
- Self-report resource use and utility were measured at the baseline visit and covered the period for the six months prior
- Patient Level Information Costing System (PLICS) data for secondary care resource use covered the period from 180 days before randomisation to their final follow-up visit

**Table 1:** Questionnaire timing.

| Location | Measure |
| --- | --- |
| **Baseline** |  |
| At Site | Demographics |
| At Site | Medical history |
| At Home | ISI (primary carer) |
| At Home | CHU9D (child if $\geq 7$ years) |
| At Home | CHU9D proxy (primary carer if child $< 7$ years) |
| At Home | EQ-5D-Y (child if $\geq 8$ years) |
| At Home | EQ-5D-Y proxy (primary carer if child $< 8$ years) |
| At Home | EQ-5D-5L (primary carer) |
| At Home | Resource use questionnaire |
| **Randomisation** |  |
| At Site | Seizure occurrence review |
| At Site | Hospital admissions |
| At Site | Concomitant medications CRF |
| At Site | School absences |
| **Follow-up 1** |  |
| At Site | Seizure occurrence review |
| At Site | Hospital admissions |
| At Site | Concomitant medications CRF |
| At Site | School absences |
| At Home | ISI (primary carer) |
| At Home | CHU9D (child if $\geq 7$ years) |
| At Home | CHU9D proxy (primary carer if child $< 7$ years) |
| At Home | EQ-5D-Y (child if $\geq 8$ years) |
| At Home | EQ-5D-Y proxy (primary carer if child $< 8$ years) |
| At Home | EQ-5D-5L (primary carer) |
| At Home | Resource use questionnaire |
| **Follow-up 2** |  |
| At Site | Seizure occurrence review |
| At Site | Hospital admissions |
| At Site | Concomitant medications CRF |
| At Site | School absences |
| At Home | ISI (primary carer) |
| At Home | CHU9D (child if $\geq 7$ years) |
| At Home | CHU9D proxy (primary carer if child $< 7$ years) |
| At Home | EQ-5D-Y (child if $\geq 8$ years) |
| At Home | EQ-5D-Y proxy (primary carer if child $< 8$ years) |
| At Home | EQ-5D-5L (primary carer) |
| At Home | Resource use questionnaire |

**Figure 1:**
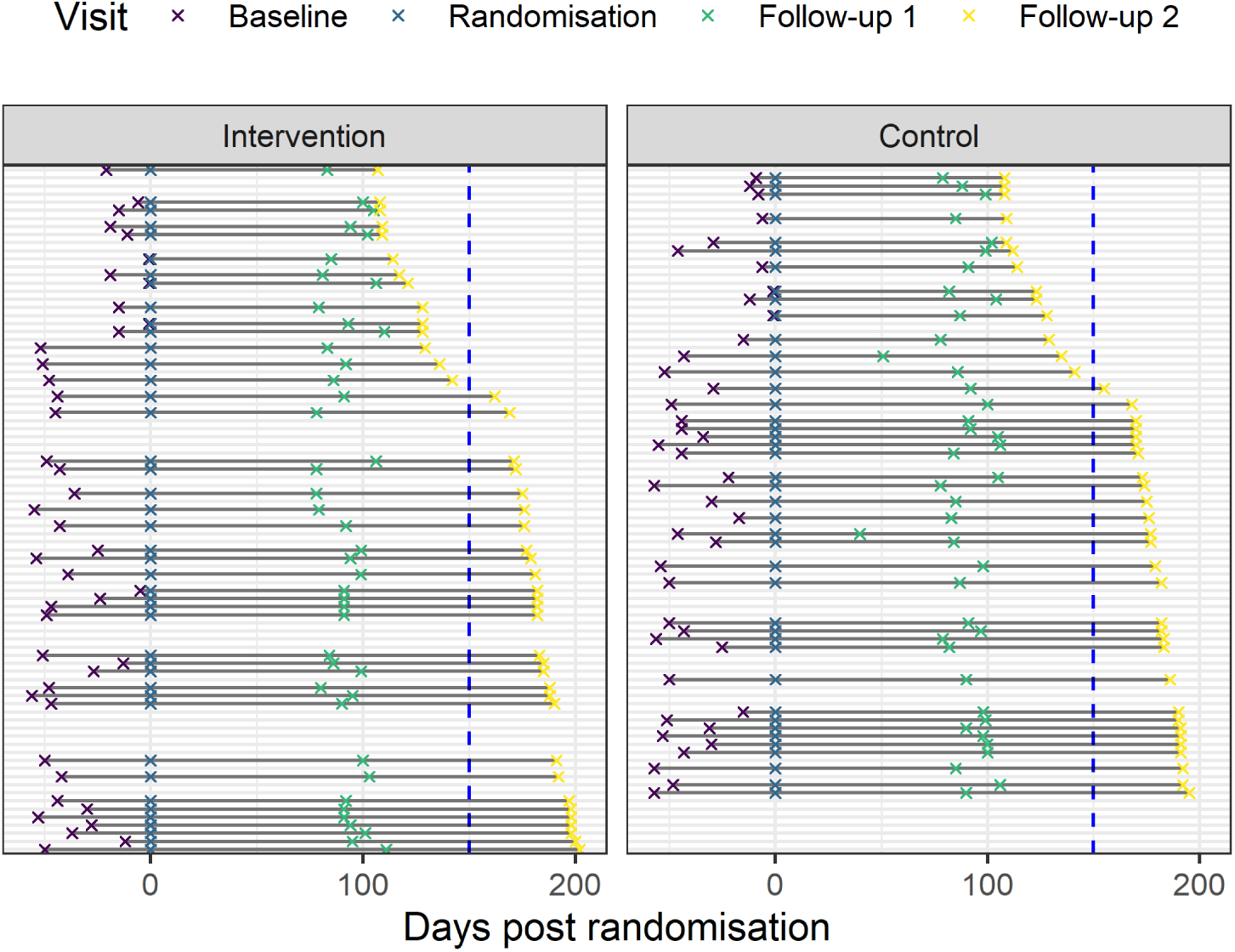
Abacus plot of visit dates. Participants with data to the right of the blue vertical dashed line were included in the primary analyses, the remainder were only included in the relevant sensitivity analyses.

#### 3.2.2 Resource use

We measured six categories of resource use: (a) primary and community care, (b) secondary care, (c) medication use, (d) out of pocket expenses, (e) productivity loss, and (f) school absences. The first three categories of resource use (a) to (c), were used in the primary analysis; the remaining categories (d) to (f), were used in sensitivity/scenario analyses.

We measured resource use using a complimentary approach. Resource use was measured irrespective of whether they were related to epilepsy or not.^10^

The primary source for secondary care resource use data was Patient Level Information and Costing System (PLICS) data, which was supplemented with responses to resource use questionnaires (RUQ) collected as part of the trial. Where both sources of data were available for a given category of resource use, PLICS data were used as the primary source of resource use data as they reflect the care recorded and RUQs introduce risk of recall bias.

Resource use and associated costs were aggregated across two periods:

1. Pre-randomisation - capturing baseline resource use and costs for each participant,
2. Within-trial - covering resource use and costs during the trial period.

##### 3.2.2.1 PLICS

PLICS data are the data used to cost individual episodes of patient care.^11^ The PLICS data requested for the analysis included: outpatient (OP) data, admitted patient care (APC) data, and emergency care (EC) data. PLICS data were the primary source of resource use data for OP, APC, and EC with each episode of care being assigned a Health Resource Group (HRG) code.

There were 24 sites with participants randomised in the study, 22 of which has participants in the primary analysis population. Once the final follow-up for a given site had been completed, the Trial Manager requested that the PLICS data from that site were sent to the Health Economist (WH). When required, this request was followed up with reminders and offers of support by (a) the Trial Manager, (b) the Chair of the Programme Steering Committee, and (c) the Health Economist.

PLICS data were requested for each participant for the period starting 180 days pre-randomisation up to their final scheduled follow-up date, whichever was earlier of 180 days post-randomisation or 2024-01-31. Where an episode of care overlapped randomisation, costs were apportioned accordingly to the relevant time frame.

PLICS data were only available as requested from sites in England, one of which did not provide any data (*n_participants_* = 2). The one site in Scotland was unable to provide PLICS data (*n_participants_* = 2). The site in Northern Ireland (*n_participants_* = 3) and site in Wales (*n_participants_* = 4) were unable to provide PLICS data, however did provide similar data that we were able to include in the analysis. Consequently, there were four randomised participants for whom PLICS data (or similar) were not available, three of whom were in the primary analysis population.

In the OP data provided, it was possible to assign treatment function codes and HRGs, thereby allowing these data to be included. The APC and EC data were not provided in sufficient detail to assign HRGs. For each of these types of PLICS data from these sites, participants with observed resource use were assigned missing values and those with no observed resource use were assigned zeros.

##### 3.2.2.2 Resource Use Questionnaires

The RUQs were administered at each of the baseline, Follow-up 1, and Follow-up 2 visits; the timings of which are described in *Section 3.2.1*. The RUQs were completed electronically by the primary carer at each study timepoint and were asked to report participants’ use of:

1. Primary Care, (a) GP appointments, (b) Community care (other NHS),
2. Secondary Care, (a) Outpatient care, (b) Emergency care, (c) Day case attendances, (d) Admitted patient care, (e) Other NHS healthcare or social care professionals, (f) Tests (e.g., magnetic resonance imaging (MRI) scan, or electroencephalogram (EEG) test),
3. Medication use,
4. Productivity loss,
5. Out of pocket expenses, (a) Over the counter medicines, (b) Non-NHS (e.g., a private educational psychologist, or private sleep consultant), (c) Related travel expenses (e.g., public transport, fuel for the car, parking fees),
6. School absences.

**Caution**

There were the three following issues with the RUQs:

1. The stem for the question about other NHS healthcare or social care professionals was “because of their condition.” This may have led to under-reporting of costs not directly related to the child’s condition,
2. The question about number of days absent from school due to illness asked about the number of days missed in the last six-months at the first and second follow-up. This may have led to over-reporting of school absences,
3. The follow-up questions asked about resource use in the last 3-months; where participants second follow-up visit was early, this may have led to over-reporting of resource use.

##### 3.2.2.3 Concomitant medication

Medication use was recorded at each visit, to calculate the total cost of concomitant medication for each participant in the period up to baseline and within the trial follow-up period. The cost per medication was estimated by multiplying the number of units used by the relevant unit cost. The total cost of concomitant medication was calculated by summing the costs of each medication for each participant.

We made the following assumptions where dates were missing, incomplete, or otherwise unclear:

- If medication use began before 2022-03-03, they began > 180 days before first participant was randomised, therefore we assumed use for every day in the baseline period (180 days),
- If medication use is ongoing, medication use continues to the last follow-up visit,
- That partial dates are 1st of the month if month present, otherwise 1st of the year,
- Where multiple doses are listed in a single entry, the mean dose is used for costing, unless other details provided suggest that this is an unreasonable assumption (e.g., recorded as “10mg for 3 weeks and 20mg for one week”),
- Where strength values are missing, this has been filled based on the mode dose for that type of medication in the trial data.

#### 3.2.3 Unit Costs

Resource-use was valued in monetary terms (£GBP) at the time of analysis (cost year: 2021-2022) and costs were not discounted as the trial follow-up was less than one-year. Adjustments were made for inflation, when necessary, using the Hospital & Community Health Services (HCHS) Index according to the compendium of Personal Social Services Research Unit (PSSRU) Unit Costs of Health and Social Care.^12^ Total costs for resource-use were calculated by multiplying the unit cost per item by the recorded number of times that each resource was used.

For primary and community care, costs were taken from the PSSRU^12^ where a unit cost was available without making assumptions, and from the NHS Schedule of Costs^13^ for secondary care. Where unit costs were not available from these sources, we documented the alternate sources and any assumptions. We costed medicines with the mean unit cost of that medicine listed in the Drug Tariff for medicines.^14^

The cost of the intervention was estimated as:

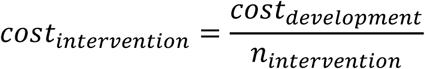

where developments costs (*cost_development_*) were taken from the details provided by the research team and comprised seven weeks of development time and three weeks of testing of the COSI. Details of the development process for COSI are detailed elsewhere.^15^

We considered two scenarios where the development cost for COSI varied:

1. The cost of a new *configurable* COSI instance, where content could be edited by non-technical staff. The costs for this version would include 8 weeks of development, three weeks of testing, initial setup, and ongoing hosting and support,
2. The cost of a new *static* COSI instance. The costs for this version would include 3 weeks development, one week testing, initial setup, and ongoing hosting and support.

We also explored how the cost-effectiveness of COSI varied as the population using it changed by setting the denominator for the intervention cost to each percentile of the eligible population (see *Table 4*).

We estimated productivity loss costs using the human capital method,^16^ multiplying the days absent from work with average annual earnings data^17^ and the cost of school absences using the average annual spending per pupil in the UK^18^ divided by the minimum number of school days in a year (190)^19^.

**Table 2:** Unit costs (£GBP) used in the economic evaluation. CL = consultant led, EL = elective, NS = National Schedule, LCTC = Liverpool Clinical Trials Centre, ASHE = Annual Survey of Household Earnings, PSSRU = Personal Social Services Research Unit.

| Resource | Source | Unit Cost (£GBP) |
| --- | --- | --- |
| Accident & Emergency department attendance | NS of NHS costs<sup>13</sup> | 241.99 |
| Actual cost of COSI development and testing | LCTC | 22,844.86 |
| Ambulance | NS of NHS costs<sup>13</sup> | 390.08 |
| Asthma clinic | NS of NHS costs<sup>13</sup> | 87.42 |
| Blood test | NS of NHS costs<sup>13</sup> | 2.96 |
| CT Scan (<6 years old) | NS of NHS costs<sup>13</sup> | 86.73 |
| CT Scan (≥ 6 years old) | NS of NHS costs<sup>13</sup> | 167.77 |
| Child community health | PSSRU<sup>20</sup> | 70.88 |
| Children and Adolescent Mental Health Services, Outpatient Attendances | NS of NHS costs<sup>13</sup> | 383.46 |
| Cost of developing a new configurable COSI | LCTC | 24,688.07 |
| Cost of developing a new static COSI | LCTC | 9,520.45 |
| Costs outside of cost perspective | - | 0.00 |
| Daycase, paediatric epilepsy | NS of NHS costs<sup>13</sup> | 1,052.84 |
| Dentist appointment | NS of NHS costs<sup>13</sup> | 269.94 |
| ECG (<6 years old) | NS of NHS costs<sup>13</sup> | 56.79 |
| ECG (≥ 6 years old) | NS of NHS costs<sup>13</sup> | 66.05 |
| EEG test | NS of NHS costs<sup>13</sup> | 266.71 |
| EL paediatric episode - epilepsy | NS of NHS costs<sup>13</sup> | 4,934.96 |
| EL paediatric episode - non-epilepsy | NS of NHS costs<sup>13</sup> | 6,131.14 |
| GP Nurse appointment, home | NS of NHS costs<sup>13</sup> | 53.74 |
| GP Nurse appointment, practice | PSSRU<sup>12</sup> | 13.43 |
| GP Nurse appointment, telephone/video | PSSRU<sup>12</sup> | 8.69 |
| GP appointment, home | Out of hours GP services in England<sup>21</sup> | 78.14 |
| GP appointment, practice | PSSRU<sup>12</sup> | 41.00 |
| GP appointment, telephone/video | PSSRU<sup>12</sup> | 15.50 |
| Generic paediatric outpatient attendance | NS of NHS costs<sup>13</sup> | 251.98 |
| Genetic test | NS of NHS costs<sup>13</sup> | 44.09 |
| Health visitor | NS of NHS costs<sup>13</sup> | 94.83 |
| Hearing test | NS of NHS costs<sup>13</sup> | 88.00 |
| LCTC hosting and support for COSI | LCTC | 513.60 |
| MRI Scan (<6 years old) | NS of NHS costs<sup>13</sup> | 250.30 |
| MRI Scan (≥ 6 years old) | NS of NHS costs<sup>13</sup> | 161.12 |
| Mean daily earnings | ONS ASHE<sup>17</sup> | 124.77 |
| Mean hourly earnings | ONS ASHE<sup>17</sup> | 18.79 |
| Non-EL Inpatient - Long Stay, epilepsy | NS of NHS costs<sup>13</sup> | 5,507.81 |
| Non-EL Inpatient - Short Stay, ear, nose, throat | NS of NHS costs<sup>13</sup> | 610.56 |
| Non-EL Inpatient - Short Stay, epilepsy | NS of NHS costs<sup>13</sup> | 841.98 |
| Occupational therapist, one-to-one | NS of NHS costs<sup>13</sup> | 167.91 |
| Optician appointment | NS of NHS costs<sup>13</sup> | 138.16 |
| Outpatient, CL, Paediatric Epilepsy Service, Face-to-Face | NS of NHS costs<sup>13</sup> | 307.70 |
| Outpatient, CL, Paediatric Epilepsy Service, Non-Face-to-Face | NS of NHS costs<sup>13</sup> | 212.55 |
| Outpatient, CL, excluding Paediatric Epilepsy Service, Face-to-Face | NS of NHS costs<sup>13</sup> | 194.39 |
| Outpatient, CL, excluding Paediatric Epilepsy Service, Non-Face-to-Face | NS of NHS costs<sup>13</sup> | 156.17 |
| Outpatient, Non-CL, Paediatric Epilepsy Service, Face-to-Face | NS of NHS costs<sup>13</sup> | 139.32 |
| Outpatient, Non-CL, Paediatric Epilepsy Service, Non-Face-to-Face | NS of NHS costs<sup>13</sup> | 108.95 |
| Outpatient, Non-CL, excluding Paediatric Epilepsy Service, Face-to-Face | NS of NHS costs<sup>13</sup> | 122.19 |
| Outpatient, Non-CL, excluding Paediatric Epilepsy Service, Non-Face-to-Face | NS of NHS costs<sup>13</sup> | 111.39 |
| Physiotherapist | NS of NHS costs<sup>13</sup> | 132.15 |
| Psychologist | NS of NHS costs<sup>13</sup> | 252.53 |
| Sickness related school absence | <sup>19</sup> | 28.92 |
| Social worker (children's services) | PSSRU<sup>20</sup> | 53.41 |
| Speech and language therapist | NS of NHS costs<sup>13</sup> | 143.21 |
| Talk therapy | PSSRU<sup>20</sup> | 125.01 |
| Ultrasound scan | NS of NHS costs<sup>13</sup> | 77.74 |

**Table 3:** Unit costs (£GBP) for ten most frequently used medicines.

| Medicine | Unit | Unit Cost (£GBP) | n |
| --- | --- | --- | --- |
| Levetiracetam | mg | 0.0006 | 25 |
| Carbamazepine | mg | 0.0003 | 19 |
| Sodium valproate | mg | 0.0024 | 16 |
| Lamotrigine | mg | 0.0491 | 12 |
| Melatonin | mg | 0.5885 | 11 |
| Ethosuximide | mg | 0.0139 | 4 |
| Clobazam (1.5mg/ml) | ml | 0.6167 | 3 |
| Methylphenidate | mg | 0.0481 | 3 |
| Midazolam 7.5mg | dose | 22.2500 | 3 |
| Sodium valproate | ml | 0.0285 | 3 |

#### 3.2.4 Health utilities

The health outcome measure for the primary economic analysis was the quality-adjusted life year (QALY), generated from utility data measured using the Child Health Utility 9 Dimension Index (CHU9D).^22^ Participants ≥ 7 years old completed the CHU9D themselves, and the primary carer of those < 7 years old complete a proxy version.

QALYs were always estimated using the area under the curve method and there was no discounting as the time horizon was less than one year.

Sensitivity analyses considered utilities measured using different instruments and the health outcomes of the participants primary carer. These sensitivity analyses were:

1. Estimating participant utility using the EuroQol Five Dimensions Youth (ED-5D-Y).^23^ As no UK youth value set is available, the adult value set was used to estimate utility scores,
2. Including primary carer QALYs additively with utility scores estimated using the EuroQol Five Dimension Five Level (EQ-5D-5L)^24^ valued using the NICE DSU cross walk.^25,26^ The age and sex of the primary carer were not recorded. To use the algorithm for estimating utilities from EQ-5D-5L data based on the EQ-5D-3L value sets, these data are required. Therefore, we assumed that parents were female and were 30 when the child was born. Subtracting the mean participant age at baseline (8.79 years old) from the year 2022 results in 2013, the standardised mean age of mothers in 2013 was 30.0 years old,^27^
3. Including primary carer QALYs additively with utility scores estimated using the Insomnia Severity Index (ISI), with scores mapped using Model 1 as proposed by Gu et al.^28^

Across all utility measures, missingness was at the scale and not the item level. Therefore, any imputation was carried out on utility estimates and not item responses.

### 3.3 Missing data

We examined missing data by assessing the patterns and proportions of missingness to determine the most appropriate methods for handling missing data based on likely mechanisms of missingness.^29^ We imputed missing data using multiple imputation with chained equations (MICE)^30^ with the *mice()* function from the *mice* package^31^ and the *2l.pmm()* extension from the *miceadds* package^32^, which allows for multilevel imputation. If data are not missing at random, complete case analyses can lead to substantive bias, that may change cost-effectiveness decisions.^29^ Multiple imputation with chained equations provides unbiased results, assuming that data are missing at random (MAR).^29,30^

We carried out our analyses on multiply imputed data to (a) minimise potential bias from a complete case analysis and (b) account for uncertainty in estimates due to the missing data. We used 70 imputed data sets for our analyses based on the largest fraction of missing information.^34^ Given that there were missing data in the covariates for the analysis models, we used multiple imputation before the mixed models for the analysis of the data.^35^

We imputed costs and utilities at each measurement time point. The imputation models included variables used in the analysis models, randomisation variables, and other variables likely to be associated with missingness. These variables were measured at the same or earlier time points relative to the target variable. This was done to maximise the use of available data, reduce the risk of bias in the imputation models, and to preserve randomisation.^36^

Given the small sample size, combined with high missingness (>70% for some items, see *Table 19*) meant that we imputed the data for both treatment groups together, including *treatment* as a covariate (which may have underestimated uncertainty in estimates),^37^ and included site as a random (rather than fixed) effect to reduce the degrees of freedom used by *site*. Both steps allowed us to include more relevant variables in the imputation models whilst not over-identifying the imputation models.^36^ Thereby following the general recommendations to over-rather than under-specify imputation models and making the missing at random assumption more plausible.^37^

We used two-level predictive mean-matching (PMM) to impute measured variables, and passive imputation for variables derived from measured variables (e.g., QALYs, total trial costs), to retain the relationships between the variable of interest and those it is derived from.^36^ Costs were divided by 1,000 and seizure counts were Winsorized to aid the imputation models by keeping variables on similar scales.

Where multiple imputation and bootstrapping were used in combination, the data were imputed first and then bootstraps were taken from the imputed datasets. This was done as the small sample combined with high level of missingness for some variables meant that imputing within bootstraps (the preferred method) did not work. This may mean that uncertainty in the estimates using this method is underestimated.^39^

We carried out several sensitivity analyses to test the robustness of the primary analysis to the imputation methods used. The first of these was a sensitivity analysis using single-level PMM instead of the two-level PMM in the primary case analysis. We also modelled data using a *within-model* imputation approach to handle missing data, modelling the missing data directly during model estimation with the *mi* command in *brms*. This method treats missing values as additional unknown parameters (*X_miss_*) alongside the analysis model parameters (*θ*) in a single joint model for *p*(*θ*, *X_miss_*| *X_obs_*, *Y*). Here, *X_obs_* denotes fully observed covariates and *Y* denotes the observed outcome. At each iteration of the Monte Carlo Markov chain sampling, draws of *X_miss_* are taken from the conditional posterior given the observed data and the current values of *θ*. These updated draws of *X_miss_* then inform the subsequent iteration for *θ*, and the final posterior distribution thus incorporates uncertainty from both the imputation and the parameter estimation processes.^40^

To explore the sensitivity of our primary analysis, and the underlying imputed CHU9D utility scores, to possible missing not at random (MNAR) mechanisms, we conducted two sensitivity analyses using the *δ*-adjustment method, focusing on CHU9D utility scores:

1. MNAR based on participant utility scores (CHU9D and EQ-5D-Y),
2. MNAR based on primary carer EQ-5D-5L utility scores (EQ-5D-5L and ISI).

The *δ*-adjustment method adds a constant (*δ*) to the imputed utility values, simulating a range of plausible shifts from the assumption that data are MAR.^29,41,42^ We use the *post* function in the *mice* package to apply these deterministic adjustments, ensuring values remained within a plausible range (i.e., a maximum of 1.0 for all utility measures, a minimum of 0 for CHU9D and ISI, and a minimum of -0.534 for EQ-5D-Y and EQ-5D-5L). The primary analysis model was then refitted to each adjusted dataset to quantify how MNAR assumptions might alter the results.

For each value of *δ*, incremental costs and QALYs are estimated from a joint model applied to 70 imputed data sets, which are then used to estimate the INMB. Consequently, small non-monotonic variations in INMB can occur, as seen most clearly in the carer utility MNAR scenario. This is a normal feature of simulation-based approaches that use multiple imputations: each imputed data set introduces additional sampling variation, and thus the aggregated estimates may occasionally depart from the overall expected monotonic trend.

### 3.4 Data Analysis

The primary analysis was based on the intention to treat approach, including all randomised participants who were eligible to attend their second follow-up at 6 ± 1 months. Statistical tests were two-sided, with credibility-intervals (CrIs) and confidence-intervals (CIs) reported at 95%.

Based on the imputed data, total costs and QALYs during the six-month trial were calculated, with summary statistics generated by randomised treatment group.

#### 3.4.1 Regression analyses

We estimated the total mean costs and QALYs for each arm using multivariate Bayesian regression models with each imputed data set. The models were run using the *brm_multiple()* function from the *brms* package,^45^ with a hurdle gamma distribution for costs^46^ and a normal distribution for QALYs.

The average marginal treatment effect for costs and QALYs were estimates with random effects for treatment site and were adjusted for any imbalances in baseline costs (continuous), utilities (continuous), and randomisation variable (using sleep medication at baseline; binary), by including these variables as covariates.^48^

The models were run with default prior distributions (*Table 17*), four chains, 7,500 warm-up iterations, and 7,500 sampling iterations. Convergence was assessed graphically and using the R-hat, effective sample size (ESS) bulk and ESS tail statistics, and the model was considered to have converged when the R-hat statistic was < 1.1 and the ESS statistics were ≥ 400 for all parameters.^49^ Unless stated otherwise, estimates are reported as the mean [95% credibility interval (CrI)].

#### 3.4.2 Incremental analysis

The cost-effectiveness of SC + COSI compared to SC alone was assessed by its incremental cost-effectiveness ratio (ICER), calculated as:

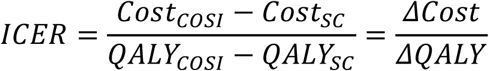

where:

*Cost_COSI_* is the mean cost in the SC + COSI group,

*Cost_SC_* is the mean cost in the SC group,

*QALY_COSI_* is the mean QALYs in the SC + COSI group,

*QALY_SC_* is the mean QALYs in the SC group,

*ΔCosts* is the difference in mean total costs between intervention groups (*Cost_COSI_* – *Cost_SC_*),

*ΔQALY* is the difference in mean QALYs between intervention groups (*QALY_COSI_* – *QALY_SC_*).

Incremental net monetary benefits (INMB) were also calculated at the £20,000 and £30,000 per QALY thresholds as:

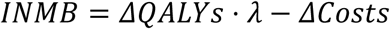

where *λ* is the cost-effectiveness threshold.^9^

Uncertainty in the costs and QALYs were evaluated using draws from the joint posterior distribution of the estimates from the regression analyses. Uncertainty in the ICER was represented graphically with a cost-effectiveness plane and a cost-effectiveness acceptability curve.^50^

#### 3.4.3 Sensitivity analyses

*Table 4* provides a summary of the sensitivity analyses conducted. The sensitivity analyses were a combination of those pre-specified in the HEAP (*Section 10.2*) and additional exploratory analyses.

**Table 4:**
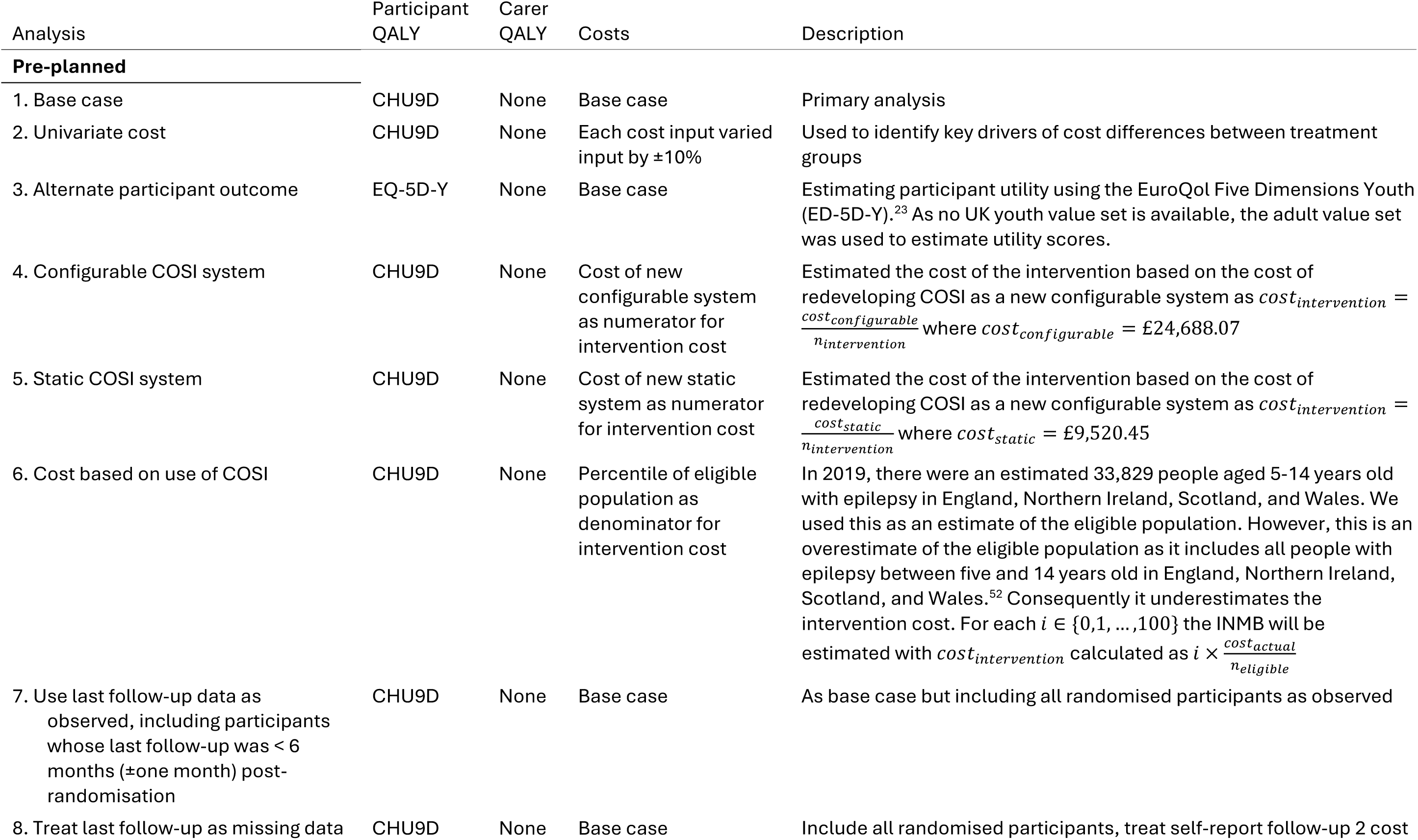
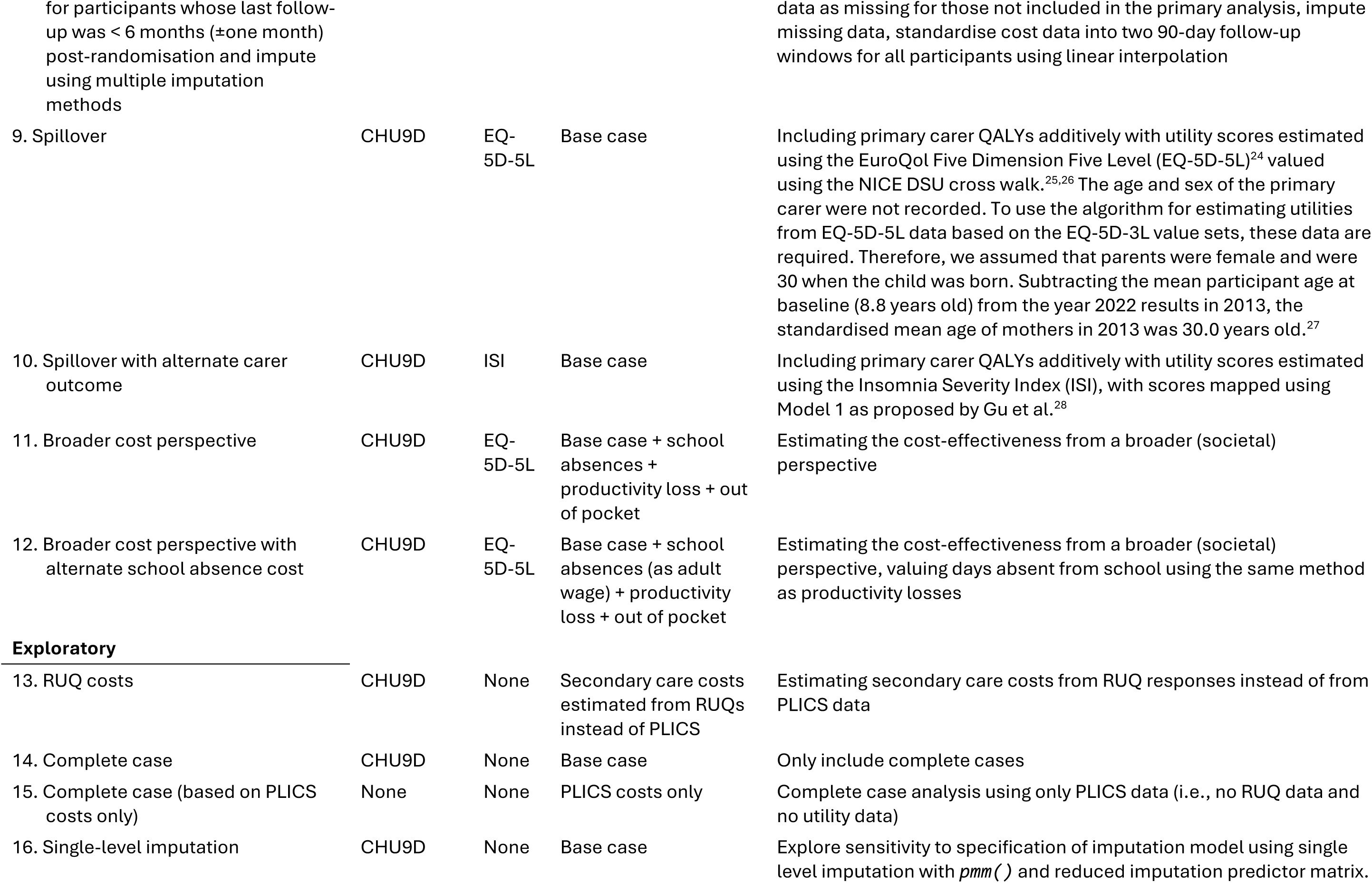
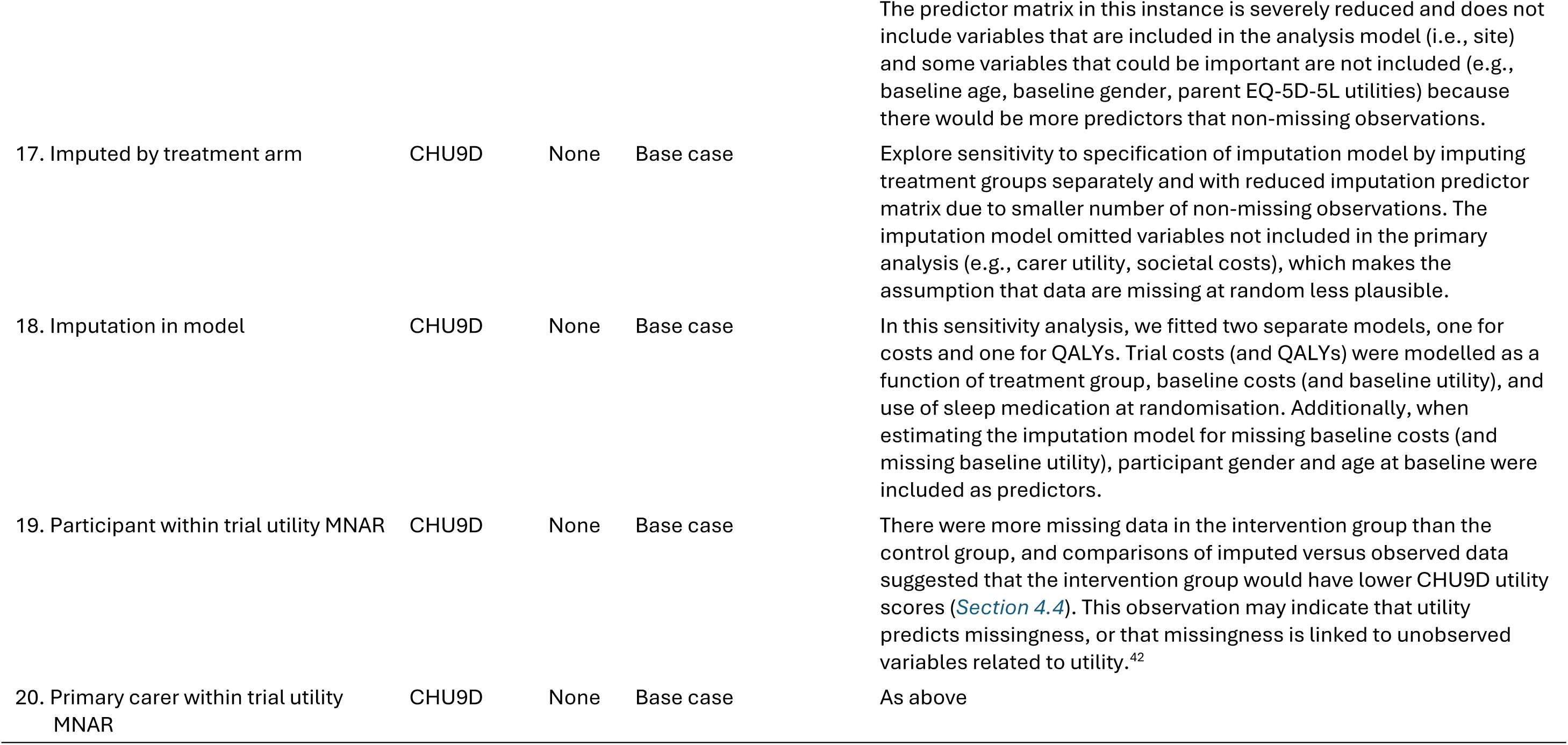
List of sensitivity analyses.

### 3.5 Deviations from analysis plan

In the HEAP, we stated that we would bootstrap analyses to represent uncertainty. Due to higher than anticipated levels of missingness in the CHU9D responses at Follow-up 2, which caused computation problems in the analyses, we opted to use Bayesian methods to estimate parameters of interest and to represent their uncertainty.^52^

## 4. Results

### 4.1 Participants

In total, 85 participants were randomised, 58 of whom were randomised early enough to be followed up for 6 ± 1 months prior to the trial closing and therefore included in the primary analysis. There were 29 participants in the control group and 29 in the intervention group. The remaining 27 participants were randomised too late to be followed up for at least five months and were therefore excluded from the primary analysis. *Table 5* summarises participant demographics for the primary analysis population and *Table 18* shows the demographics for all randomised participants.

**Table 5:**
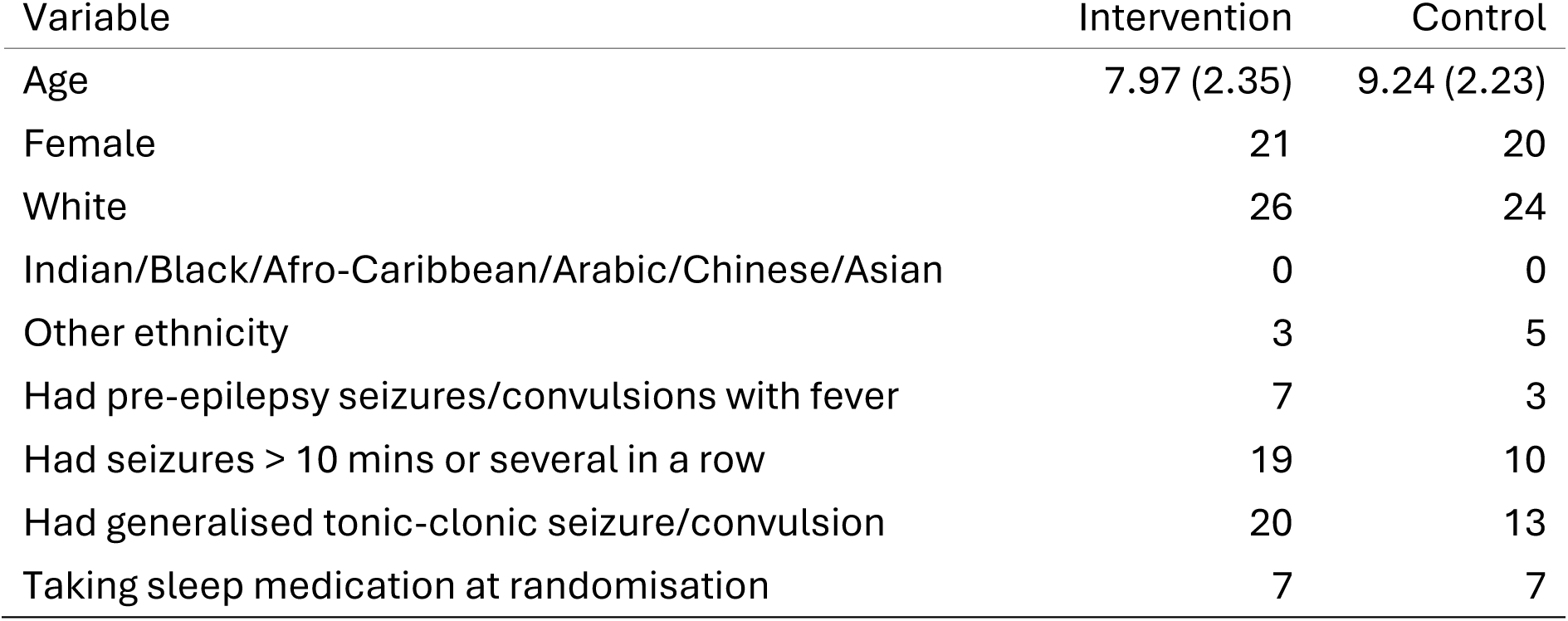
Summary of participant demographics. Data are mean (SD) for age and counts for all other variables.

| Variable | Intervention | Control |
| --- | --- | --- |
| Age | 7.97 (2.35) | 9.24 (2.23) |
| Female | 21 | 20 |
| White | 26 | 24 |
| Indian/Black/Afro-Caribbean/Arabic/Chinese/Asian | 0 | 0 |
| Other ethnicity | 3 | 5 |
| Had pre-epilepsy seizures/convulsions with fever | 7 | 3 |
| Had seizures > 10 mins or several in a row | 19 | 10 |
| Had generalised tonic-clonic seizure/convulsion | 20 | 13 |
| Taking sleep medication at randomisation | 7 | 7 |

### 4.2 Data completeness

*Table 6* shows the completeness of data included in the primary analysis, grouped by the timepoint the data were collected and as aggregated for imputation models.

*Table 19* shows the completeness of data for all randomised participants. There was a high level of missing data in several variables, with the highest overall level of missing data being in CHU9D responses at Follow-up 2 (67.24 % missing). At baseline, data completeness was approximately similar across the treatment groups. The level of missing data increased at each visit and at a greater rate for the treatment group.

*Figure 2* shows an illustrative example of this for CHU9D and RUQ primary care cost data.

**Figure 2:**
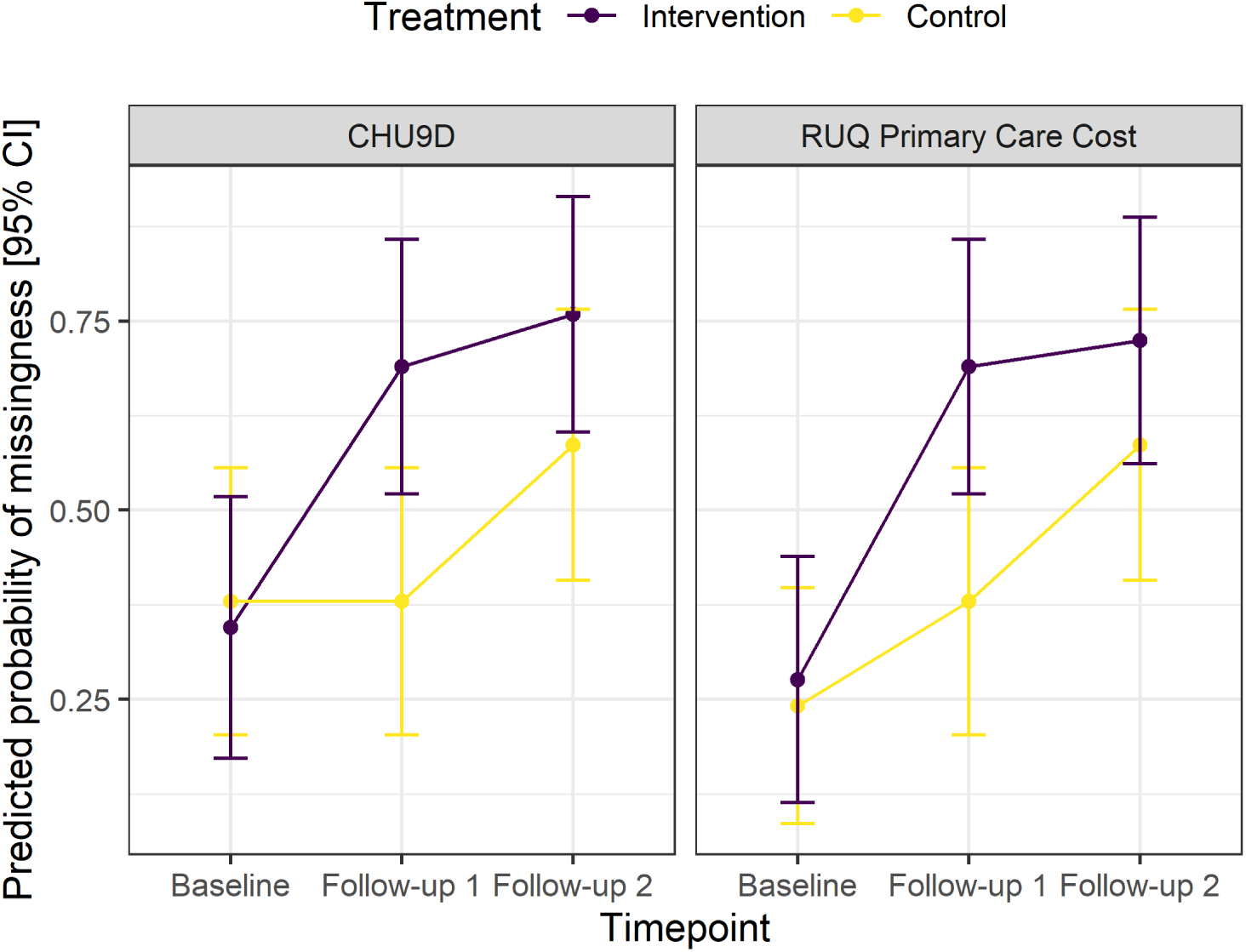
Predicted probability of missingness [95% confidence interval] for CHU9D and resource use primary care costs, summarised by treatment group.

**Table 6:** Summary of data completeness by treatment group for primary analysis population.

| Variable | Intervention (n = 29) |  |  | Control (n = 29) |  |  |
| --- | --- | --- | --- | --- | --- | --- |
|  | Complete | Missing | % missing | Complete | Missing | % missing |
| **Baseline** |  |  |  |  |  |  |
| CHU9D | 19 | 10 | 34.5 | 18 | 11 | 37.9 |
| EQ-5D-5L parent | 21 | 8 | 27.6 | 23 | 6 | 20.7 |
| EQ-5D-Y | 18 | 11 | 37.9 | 17 | 12 | 41.4 |
| ISI parent | 21 | 8 | 27.6 | 23 | 6 | 20.7 |
| Medication | 29 | 0 | 0.0 | 29 | 0 | 0.0 |
| RUQ primary care | 21 | 8 | 27.6 | 22 | 7 | 24.1 |
| RUQ secondary care | 21 | 8 | 27.6 | 22 | 7 | 24.1 |
| RUQ societal | 21 | 8 | 27.6 | 22 | 7 | 24.1 |
| **Randomisation** |  |  |  |  |  |  |
| PLICS APC | 25 | 4 | 13.8 | 29 | 0 | 0.0 |
| PLICS EC | 26 | 3 | 10.3 | 29 | 0 | 0.0 |
| PLICS OP | 25 | 4 | 13.8 | 27 | 2 | 6.9 |
| School absences | 29 | 0 | 0.0 | 29 | 0 | 0.0 |
| **Follow-up 1** |  |  |  |  |  |  |
| CHU9D | 9 | 20 | 69.0 | 18 | 11 | 37.9 |
| EQ-5D-5L parent | 9 | 20 | 69.0 | 18 | 11 | 37.9 |
| EQ-5D-Y | 9 | 20 | 69.0 | 18 | 11 | 37.9 |
| ISI parent | 9 | 20 | 69.0 | 20 | 9 | 31.0 |
| RUQ primary care | 9 | 20 | 69.0 | 18 | 11 | 37.9 |
| RUQ secondary care | 9 | 20 | 69.0 | 18 | 11 | 37.9 |
| RUQ societal | 9 | 20 | 69.0 | 18 | 11 | 37.9 |
| School absences | 22 | 7 | 24.1 | 28 | 1 | 3.4 |
| **Follow-up 2** |  |  |  |  |  |  |
| CHU9D | 7 | 22 | 75.9 | 12 | 17 | 58.6 |
| EQ-5D-5L parent | 9 | 20 | 69.0 | 12 | 17 | 58.6 |
| EQ-5D-Y | 8 | 21 | 72.4 | 12 | 17 | 58.6 |
| ISI parent | 9 | 20 | 69.0 | 12 | 17 | 58.6 |
| RUQ primary care | 8 | 21 | 72.4 | 12 | 17 | 58.6 |
| RUQ secondary care | 8 | 21 | 72.4 | 12 | 17 | 58.6 |
| RUQ societal | 8 | 21 | 72.4 | 12 | 17 | 58.6 |
| School absences | 22 | 7 | 24.1 | 26 | 3 | 10.3 |
| **Trial** |  |  |  |  |  |  |
| Medication | 27 | 2 | 6.9 | 26 | 3 | 10.3 |
| PLICS APC | 26 | 3 | 10.3 | 29 | 0 | 0.0 |
| PLICS EC | 26 | 3 | 10.3 | 28 | 1 | 3.4 |
| PLICS OP | 25 | 4 | 13.8 | 29 | 0 | 0.0 |

### 4.3 Resource use and costs

*Table 7* summarises the mean and standard deviation for frequencies of resource use at each timepoint for each randomised treatment group based on the RUQs. *Table 8* shows unadjusted mean [95% CI] resource use costs aggregated by type of resource and timepoint for each treatment group based on the imputed and bootstrapped data. Overall, the majority of costs are for secondary care both from outpatient and admitted patient care. However, the total APC costs are driven by the intervention group. There is a smaller difference in the cost estimates for secondary care based on PLICS and RUQ data in the baseline/randomisation data than during the within-trial period.

In the PLICS data, there were 81 unique HRG codes used across the baseline and trial periods, with 291 episodes of care, 16 of which were APC, 237 of which were OP care, and 38 of which were EC. *Table 9* shows the top five most frequently used HRG codes within each category across the baseline and trial periods, along with their unit costs, for the primary analysis population.

Based on the imputed and bootstrapped data (*Table 8*), unadjusted baseline costs (i.e., the costs for the 180 days up to the baseline visit) were £2,261 [1,362; 3,351] and £3,816 [2,137; 5,775] for the intervention and control groups respectively. The unadjusted within trial costs were £3,068 [1,910; 4,767] and £1,184 [ 790; 1,708] for the intervention and control groups respectively. The unadjusted difference (Intervention - Control) in costs for the within trial period was £ 1,885 [ 581; 3,654].

In the primary analysis, the mean adjusted within trial cost in the intervention group was £2,021 [1,352; 3,455] and £ 790 [ 409; 1,628] for the control group. This corresponds to an incremental cost of £1,232 [535; 2,183].

**Table 7:** Observed resource use based on resource use questionnaires.

| Variable | Intervention |  |  | Control |  |  |
| --- | --- | --- | --- | --- | --- | --- |
|  | Mean | SD | n | Mean | SD | n |
| **Baseline** |  |  |  |  |  |  |
| Proportion of participants with GP appointment | 0.52 | 0.51 | 21 | 0.64 | 0.49 | 22 |
| Proportion of participants with nurse appointment at GP | 0.24 | 0.44 | 21 | 0.50 | 0.51 | 22 |
| Proportion of participants with NHS healthcare or social care professional appointment | 0.33 | 0.48 | 21 | 0.41 | 0.50 | 22 |
| Proportion of participants with non-NHS healthcare or social care professional appointments | 0.05 | 0.22 | 21 | 0.00 | 0.00 | 22 |
| Proportion of participants with outpatient clinic appointments | 0.43 | 0.51 | 21 | 0.45 | 0.51 | 22 |
| Proportion of participants with test or investigation | 0.38 | 0.50 | 21 | 0.14 | 0.35 | 22 |
| Proportion of participants with hospital admission | 0.19 | 0.40 | 21 | 0.00 | 0.00 | 22 |
| Proportion of participants who had been brought medicines from the pharmacy or any other retailer for their epilepsy | 0.43 | 0.51 | 21 | 0.27 | 0.46 | 22 |
| Proportion of participants who had money spent on travel to their healthcare and social care appointments | 0.57 | 0.51 | 21 | 0.68 | 0.48 | 22 |
| Proportion of primary carers in employment | 0.81 | 0.40 | 21 | 0.77 | 0.43 | 22 |
| Proportion of primary carers whose work situation had been affected by their child's epilepsy | 0.48 | 0.51 | 21 | 0.45 | 0.51 | 22 |
| Number of days taken of work | 1.95 | 4.40 | 21 | 8.73 | 18.47 | 22 |
| Lost earnings due to taking occasional days off work | 183.14 | 444.51 | 21 | 225.55 | 738.89 | 22 |
| Lost working hours per week due to giving up work | 1.67 | 6.58 | 21 | 2.14 | 6.08 | 22 |
| Number of weeks work lost due to giving up work | 0.48 | 1.54 | 21 | 1.59 | 5.71 | 22 |
| Lost earnings due to giving up work (£GBP) | 74.29 | 279.44 | 21 | 94.14 | 425.95 | 22 |
| Lost hours per week due to reducing hours | 0.24 | 1.09 | 21 | 2.45 | 6.39 | 22 |
| Number of weeks with reduced working hours | 1.24 | 5.67 | 21 | 1.36 | 5.57 | 22 |
| Lost earnings due to reducing hours (£GBP) | 61.90 | 283.68 | 21 | 250.27 | 954.46 | 22 |
| Number of GP surgery visits | 0.93 | 1.85 | 29 | 1.03 | 1.43 | 29 |
| Number of GP telephone or video appointments | 0.62 | 1.74 | 29 | 0.86 | 1.46 | 29 |
| Number of GP home visits (call-out) | 0.03 | 0.19 | 29 | 0.00 | 0.00 | 29 |
| Number of GP nurse visits at surgery | 0.38 | 1.32 | 29 | 0.79 | 1.42 | 29 |
| Number of GP nurse telephone or video appointments | 0.24 | 1.30 | 29 | 0.34 | 1.20 | 29 |
| Number of GP nurse home visits | 0.03 | 0.19 | 29 | 0.00 | 0.00 | 29 |
| Number of NHS healthcare or social care professional appointments | 1.48 | 3.99 | 29 | 1.62 | 5.56 | 29 |
| Number of outpatient clinic appointments, doctor, face-to-face | 0.66 | 1.17 | 29 | 0.48 | 0.83 | 29 |
| Number of outpatient clinic appointments, doctor, telephone or video | 0.17 | 0.76 | 29 | 0.07 | 0.26 | 29 |
| Number of outpatient clinic appointments, nurse, face-to-face | 0.10 | 0.41 | 29 | 0.07 | 0.26 | 29 |
| Number of outpatient clinic appointments, nurse, telephone or video | 0.14 | 0.44 | 29 | 0.14 | 0.44 | 29 |
| Number of visits to accident and emergency | 0.48 | 0.99 | 29 | 0.34 | 1.11 | 29 |
| Number of ambulances required | 0.17 | 0.54 | 29 | 0.28 | 1.00 | 29 |
| Number of day case admissions | 0.00 | 0.00 | 29 | 0.00 | 0.00 | 29 |
| Number of magnetic resonance imaging (MRI) scans | 0.21 | 0.49 | 29 | 0.03 | 0.19 | 29 |
| Number of electroencephalogram (EEG) tests | 0.28 | 0.53 | 29 | 0.14 | 0.44 | 29 |
| Number of other tests or investigations | 0.10 | 0.41 | 29 | 2.83 | 15.23 | 29 |
| Total cost of medicines bought from a pharmacy or other retailer (£GBP) | 9.72 | 25.06 | 29 | 2.76 | 6.29 | 29 |
| Total amount spent on travel to healthcare and social care appointment (£GBP) | 27.41 | 73.81 | 29 | 13.07 | 21.20 | 29 |
| Number of non-NHS healthcare or social care professional appointments | 0.10 | 0.56 | 29 | 0.00 | 0.00 | 29 |
| Number of nights admitted to hospital | 0.28 | 0.75 | 29 | 0.00 | 0.00 | 29 |
| **Randomisation** |  |  |  |  |  |  |
| Number of sickness related school absences (days) | 4.34 | 5.81 | 29 | 6.66 | 16.90 | 29 |
| **Follow-up 1** |  |  |  |  |  |  |
| Proportion of participants with GP appointment | 0.11 | 0.33 | 9 | 0.39 | 0.50 | 18 |
| Proportion of participants with nurse appointment at GP | 0.22 | 0.44 | 9 | 0.28 | 0.46 | 18 |
| Proportion of participants with NHS healthcare or social care professional appointment | 0.44 | 0.53 | 9 | 0.22 | 0.43 | 18 |
| Proportion of participants with non-NHS healthcare or social care professional appointments | 0.11 | 0.33 | 9 | 0.11 | 0.32 | 18 |
| Proportion of participants with outpatient clinic appointments | 0.22 | 0.44 | 9 | 0.17 | 0.38 | 18 |
| Proportion of participants with test or investigation | 0.22 | 0.44 | 9 | 0.17 | 0.38 | 18 |
| Proportion of participants with hospital admission | 0.00 | 0.00 | 9 | 0.00 | 0.00 | 18 |
| Proportion of participants who had been brought medicines from the pharmacy or any other retailer for their epilepsy | 0.22 | 0.44 | 9 | 0.28 | 0.46 | 18 |
| Proportion of participants who had money spent on travel to their healthcare and social care appointments | 0.44 | 0.53 | 9 | 0.39 | 0.50 | 18 |
| Proportion of primary carers in employment | 0.89 | 0.33 | 9 | 0.78 | 0.43 | 18 |
| Proportion of primary carers whose work situation had been affected by their child's epilepsy | 0.33 | 0.50 | 9 | 0.17 | 0.38 | 18 |
| Number of days taken of work | 0.67 | 1.00 | 9 | 0.22 | 0.65 | 18 |
| Lost earnings due to taking occasional days off work | 35.56 | 73.33 | 9 | 49.72 | 189.68 | 18 |
| Lost working hours per week due to giving up work | 0.00 | 0.00 | 9 | 0.00 | 0.00 | 18 |
| Number of weeks work lost due to giving up work | 0.00 | 0.00 | 9 | 0.00 | 0.00 | 18 |
| Lost earnings due to giving up work (£GBP) | 0.00 | 0.00 | 9 | 0.00 | 0.00 | 18 |
| Lost hours per week due to reducing hours | 0.00 | 0.00 | 9 | 0.00 | 0.00 | 18 |
| Number of weeks with reduced working hours | 0.00 | 0.00 | 9 | 0.00 | 0.00 | 18 |
| Lost earnings due to reducing hours (£GBP) | 0.00 | 0.00 | 9 | 0.00 | 0.00 | 18 |
| Number of sickness related school absences (days) | 4.95 | 7.03 | 22 | 4.11 | 8.18 | 28 |
| Number of GP surgery visits | 0.03 | 0.19 | 29 | 0.38 | 0.82 | 29 |
| Number of GP telephone or video appointments | 0.00 | 0.00 | 29 | 0.14 | 0.44 | 29 |
| Number of GP home visits (call-out) | 0.00 | 0.00 | 29 | 0.00 | 0.00 | 29 |
| Number of GP nurse visits at surgery | 0.07 | 0.26 | 29 | 0.34 | 0.90 | 29 |
| Number of GP nurse telephone or video appointments | 0.00 | 0.00 | 29 | 0.10 | 0.31 | 29 |
| Number of GP nurse home visits | 0.00 | 0.00 | 29 | 0.00 | 0.00 | 29 |
| Number of NHS healthcare or social care professional appointments | 0.17 | 0.47 | 29 | 0.21 | 0.56 | 29 |
| Number of outpatient clinic appointments, doctor, face-to-face | 0.17 | 0.66 | 29 | 0.21 | 0.49 | 29 |
| Number of outpatient clinic appointments, doctor, telephone or video | 0.10 | 0.56 | 29 | 0.00 | 0.00 | 29 |
| Number of outpatient clinic appointments, nurse, face-to-face | 0.07 | 0.37 | 29 | 0.03 | 0.19 | 29 |
| Number of outpatient clinic appointments, nurse, telephone or video | 0.00 | 0.00 | 29 | 0.00 | 0.00 | 29 |
| Number of visits to accident and emergency | 0.03 | 0.19 | 29 | 0.03 | 0.19 | 29 |
| Number of ambulances required | 0.00 | 0.00 | 29 | 0.00 | 0.00 | 29 |
| Number of day case admissions | 0.00 | 0.00 | 29 | 0.00 | 0.00 | 29 |
| Number of magnetic resonance imaging (MRI) scans | 0.00 | 0.00 | 29 | 0.03 | 0.19 | 29 |
| Number of electroencephalogram (EEG) tests | 0.07 | 0.26 | 29 | 0.03 | 0.19 | 29 |
| Number of other tests or investigations | 0.00 | 0.00 | 29 | 0.03 | 0.19 | 29 |
| Total cost of medicines bought from a pharmacy or other retailer (£GBP) | 2.21 | 11.14 | 29 | 2.17 | 5.50 | 29 |
| Total amount spent on travel to healthcare and social care appointment (£GBP) | 5.00 | 18.99 | 29 | 3.76 | 9.33 | 29 |
| Number of non-NHS healthcare or social care professional appointments | 0.07 | 0.37 | 29 | 1.07 | 4.11 | 29 |
| Number of nights admitted to hospital | 0.00 | 0.00 | 29 | 0.00 | 0.00 | 29 |
| **Follow-up 2** |  |  |  |  |  |  |
| Proportion of participants with GP appointment | 0.38 | 0.52 | 8 | 0.17 | 0.39 | 12 |
| Proportion of participants with nurse appointment at GP | 0.12 | 0.35 | 8 | 0.08 | 0.29 | 12 |
| Proportion of participants with NHS healthcare or social care professional appointment | 0.00 | 0.00 | 8 | 0.08 | 0.29 | 12 |
| Proportion of participants with non-NHS healthcare or social care professional appointments | 0.12 | 0.35 | 8 | 0.00 | 0.00 | 12 |
| Proportion of participants with outpatient clinic appointments | 0.12 | 0.35 | 8 | 0.25 | 0.45 | 12 |
| Proportion of participants with test or investigation | 0.12 | 0.35 | 8 | 0.00 | 0.00 | 12 |
| Proportion of participants with hospital admission | 0.00 | 0.00 | 8 | 0.08 | 0.29 | 12 |
| Proportion of participants who had been brought medicines from the pharmacy or any other retailer for their epilepsy | 0.12 | 0.35 | 8 | 0.08 | 0.29 | 12 |
| Proportion of participants who had money spent on travel to their healthcare and social care appointments | 0.12 | 0.35 | 8 | 0.25 | 0.45 | 12 |
| Proportion of primary carers in employment | 0.88 | 0.35 | 8 | 0.75 | 0.45 | 12 |
| Proportion of primary carers whose work situation had been affected by their child's epilepsy | 0.12 | 0.35 | 8 | 0.00 | 0.00 | 12 |
| Number of days taken of work | 0.75 | 2.12 | 8 | 0.00 | 0.00 | 12 |
| Lost earnings due to taking occasional days off work | 8.75 | 24.75 | 8 | 0.00 | 0.00 | 12 |
| Lost working hours per week due to giving up work | 0.00 | 0.00 | 8 | 0.00 | 0.00 | 12 |
| Number of weeks work lost due to giving up work | 0.00 | 0.00 | 8 | 0.00 | 0.00 | 12 |
| Lost earnings due to giving up work (£GBP) | 0.00 | 0.00 | 8 | 0.00 | 0.00 | 12 |
| Lost hours per week due to reducing hours | 0.00 | 0.00 | 8 | 0.00 | 0.00 | 12 |
| Number of weeks with reduced working hours | 0.00 | 0.00 | 8 | 0.00 | 0.00 | 12 |
| Lost earnings due to reducing hours (£GBP) | 0.00 | 0.00 | 8 | 0.00 | 0.00 | 12 |
| Number of sickness related school absences (days) | 5.05 | 6.74 | 22 | 5.38 | 11.02 | 26 |
| Number of GP surgery visits | 0.10 | 0.41 | 29 | 0.07 | 0.26 | 29 |
| Number of GP telephone or video appointments | 0.03 | 0.19 | 29 | 0.00 | 0.00 | 29 |
| Number of GP home visits (call-out) | 0.00 | 0.00 | 29 | 0.00 | 0.00 | 29 |
| Number of GP nurse visits at surgery | 0.03 | 0.19 | 29 | 0.03 | 0.19 | 29 |
| Number of GP nurse telephone or video appointments | 0.00 | 0.00 | 29 | 0.00 | 0.00 | 29 |
| Number of GP nurse home visits | 0.00 | 0.00 | 29 | 0.00 | 0.00 | 29 |
| Number of NHS healthcare or social care professional appointments | 0.00 | 0.00 | 29 | 0.07 | 0.37 | 29 |
| Number of outpatient clinic appointments, doctor, face-to-face | 0.03 | 0.19 | 29 | 0.07 | 0.26 | 29 |
| Number of outpatient clinic appointments, doctor, telephone or video | 0.00 | 0.00 | 29 | 0.03 | 0.19 | 29 |
| Number of outpatient clinic appointments, nurse, face-to-face | 0.00 | 0.00 | 29 | 0.03 | 0.19 | 29 |
| Number of outpatient clinic appointments, nurse, telephone or video | 0.00 | 0.00 | 29 | 0.00 | 0.00 | 29 |
| Number of visits to accident and emergency | 0.00 | 0.00 | 29 | 0.03 | 0.19 | 29 |
| Number of ambulances required | 0.00 | 0.00 | 29 | 0.03 | 0.19 | 29 |
| Number of day case admissions | 0.00 | 0.00 | 29 | 0.00 | 0.00 | 29 |
| Number of magnetic resonance imaging (MRI) scans | 0.00 | 0.00 | 29 | 0.00 | 0.00 | 29 |
| Number of electroencephalogram (EEG) tests | 0.03 | 0.19 | 29 | 0.00 | 0.00 | 29 |
| Number of other tests or investigations | 0.03 | 0.19 | 29 | 0.00 | 0.00 | 29 |
| Total cost of medicines bought from a pharmacy or other retailer (£GBP) | 0.69 | 3.71 | 29 | 0.24 | 1.30 | 29 |
| Total amount spent on travel to healthcare and social care appointment (£GBP) | 0.24 | 1.30 | 29 | 1.03 | 3.10 | 29 |
| Number of non-NHS healthcare or social care professional appointments | 0.03 | 0.19 | 29 | 0.00 | 0.00 | 29 |
| Number of nights admitted to hospital | 0.00 | 0.00 | 29 | 0.03 | 0.19 | 29 |

**Table 8:** Unadjusted imputed and bootstrapped resource use costs (£) summarised by treatment group [95% CI].

| Variable | Intervention | Control | Difference |
| --- | --- | --- | --- |
| **Baseline** |  |  |  |
| Medication | 35 [ 5; 78] | 23 [ 7; 45] | 12 [ -27; 59] |
| RUQ primary care | 345 [ 113; 632] | 268 [ 105; 501] | 77 [ -250; 405] |
| RUQ secondary care | 860 [ 471; 1,302] | 590 [ 297; 930] | 270 [ -239; 799] |
| RUQ societal | 811 [ 176; 1,707] | 2,022 [ 713; 3,619] | -1,211 [-2,926; 401] |
| **Randomisation** |  |  |  |
| PLICS APC | 236 [ 0; 654] | 561 [ 0; 1,356] | -325 [-1,172; 380] |
| PLICS EC | 55 [ 20; 103] | 93 [ 16; 192] | -38 [ -142; 52] |
| PLICS OP | 654 [ 485; 835] | 657 [ 487; 860] | -3 [ -268; 248] |
| PLICS secondary care | 945 [ 570; 1,484] | 1,311 [ 645; 2,235] | -365 [-1,375; 492] |
| Total absences | 125 [ 74; 195] | 192 [ 64; 401] | -67 [ -278; 87] |
| Total primary analysis | 2,261 [1,362; 3,351] | 3,816 [2,137; 5,775] | -1,555 [-3,728; 465] |
| **Trial** |  |  |  |
| Intervention | 788 [ 788; 788] | 0 [ 0; 0] | 788 [ 788; 788] |
| Medication | 49 [ 11; 101] | 56 [ 24; 97] | -8 [ -64; 54] |
| PLICS APC | 1,005 [ 0; 2,574] | 138 [ 0; 464] | 867 [ -230; 2,470] |
| PLICS EC | 76 [ 18; 148] | 56 [ 12; 116] | 20 [ -65; 108] |
| PLICS OP | 558 [ 402; 725] | 347 [ 233; 479] | 211 [ 8; 417] |
| PLICS secondary care | 1,639 [ 532; 3,324] | 541 [ 296; 932] | 1,097 [ -92; 2,801] |
| RUQ primary care | 132 [ 70; 205] | 159 [ 81; 251] | -26 [ -142; 78] |
| RUQ secondary care | 353 [ 187; 543] | 313 [ 164; 486] | 40 [ -195; 281] |
| RUQ societal | 185 [ 82; 305] | 160 [ 57; 284] | 25 [ -135; 184] |
| Total absences | 276 [ 173; 401] | 267 [ 117; 466] | 9 [ -217; 208] |
| Total primary analysis | 3,068 [1,910; 4,767] | 1,184 [ 790; 1,708] | 1,885 [ 581; 3,654] |

**Table 9:**
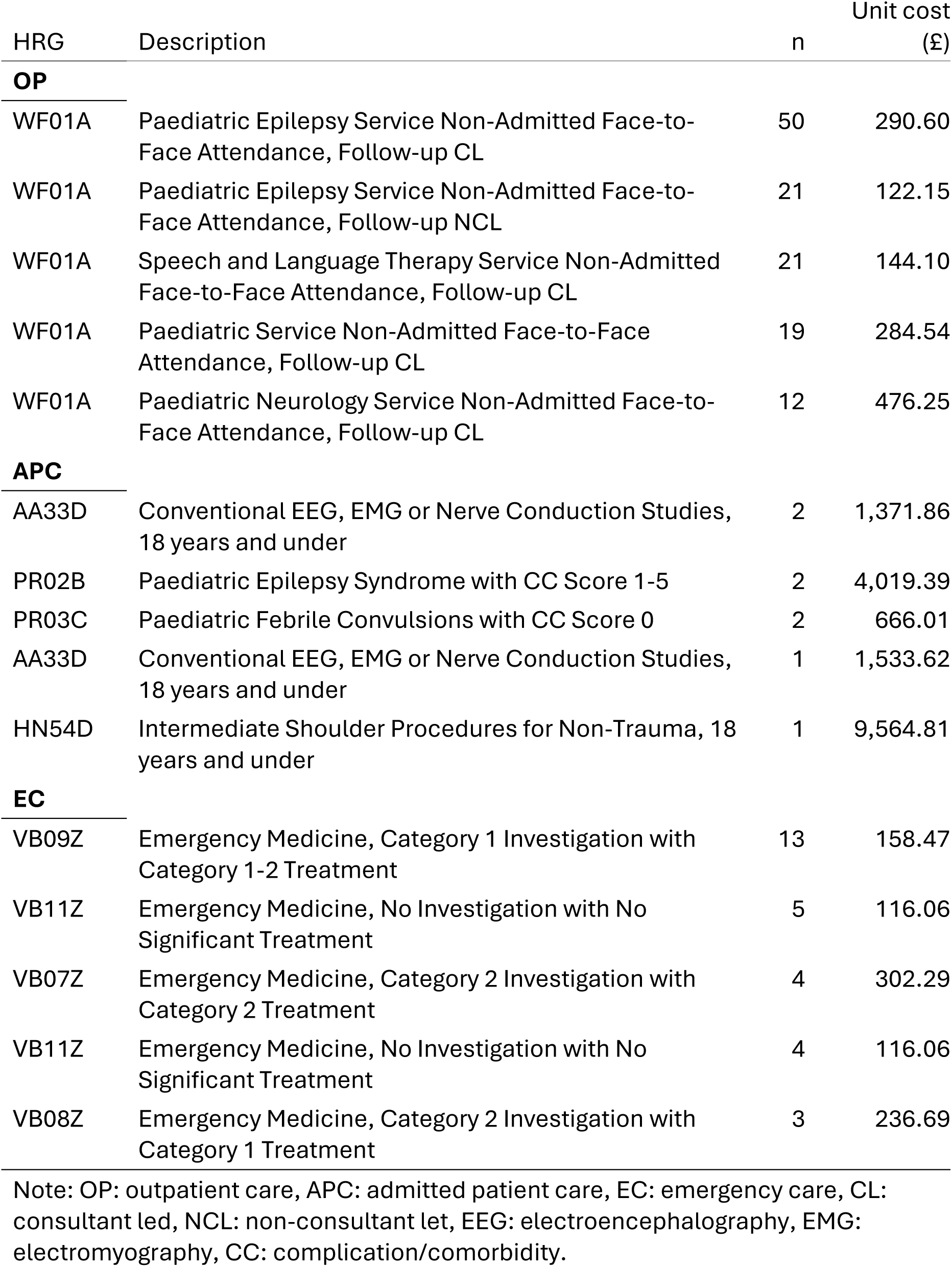
Frequency of most used HRG codes across the baseline and trial periods with unit costs. OP: outpatient care, APC: admitted patient care, EC: emergency care, CL: consultant led, NCL: non-consultant let, EEG: electroencephalography, EMG: electromyography, CC: complication/comorbidity.

### 4.4 Health utilities

Based on the imputed and bootstrapped data (*Table 11*), unadjusted baseline CHU9D utility scores were 0.80 [0.76; 0.84] for the intervention group and 0.82 [0.78; 0.85] for the control group. The unadjusted difference in baseline utility scores was -0.01 [-0.07; 0.04]. The unadjusted within-trial QALYs were similar between the groups: 0.42 [0.40; 0.43] for the intervention group and 0.41 [0.39; 0.44] for the control group. Consequently, the unadjusted difference in QALYs for the within trial period was 0.00 [−0.03; 0.03].

In the primary analysis, the estimate for mean adjusted within trial QALYs in the intervention group was 0.42 [0.39; 0.45] and 0.41 [0.38; 0.45] for the control group. This resulted in QALY difference of 0.00 [ -0.03; 0.04] (*Figure 4*).

The distributions of responses to the CHU9D questionnaire by randomised treatment group are presented in *Figure 3* based on completed responses. This shows that the most severe problems were in the tired and work domains and the lowest levels of problems were in the pain and worries domains.

Summaries of imputed and bootstrapped utility scores and QALY estimates across all measures are shown in *Table 11*, summaries of observed responses are shown in *Table 10*. Overall, no statistically significant differences were found between the intervention and control groups in utility values or QALYs at any timepoint. CHU9D utilities were consistently higher than EQ-5D-Y utilities measured at the same timepoint, similarly, ISI utilities were higher than EQ-5D-5L utilities at each timepoint.

Within the intervention group, imputed CHU9D utility values were slightly higher than corresponding observed values, potentially indicating that participants with lower utility scores were less likely to respond. This pattern was explored in the missing not at random sensitivity analyses (see *Section 4.6.2.4.2*).

*Table 12* shows the pooled correlation coefficients [95% confidence intervals] for each utility measure at the timepoints they were recorded. Correlations were generally weak and associated with considerable uncertainty, resulting in few statistically significant relationships. Given the lack of observed data (*Table 6*), there will be considerable uncertainty due to the imputation process.

**Figure 3:**
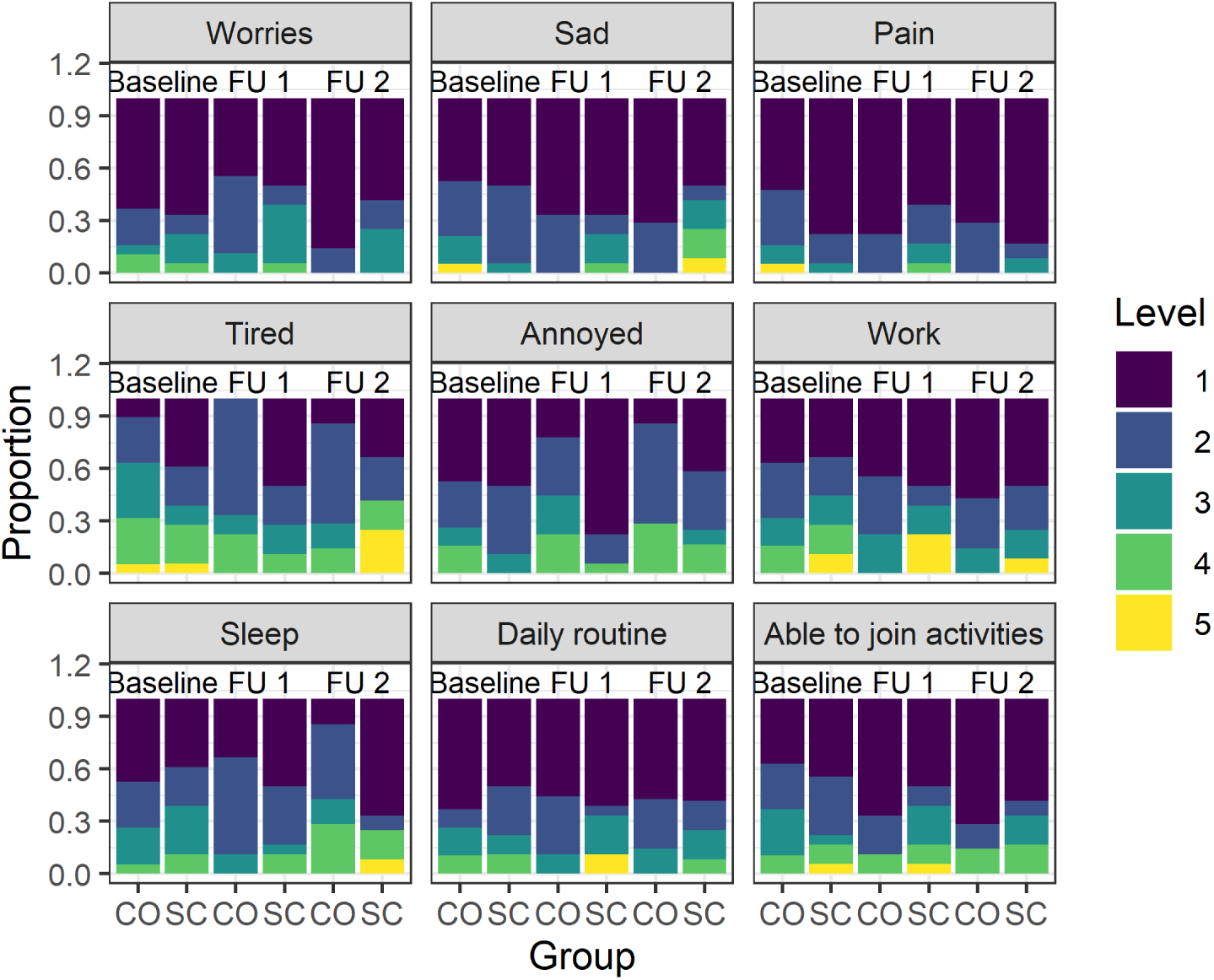
Distribution of responses to each CHU9D domain, by treatment group and timepoint. Levels ranged from 1 to 5, with 5 representing the most severe problems. Proportion is the proportion of completed responses, FU 1 = Follow-up 1, FU 2 = Follow-up 2. CO = COSI, SC = standard care.

**Table 10:** Mean utility scores for each measure, by timepoint, and intervention group for observed data.

| Variable | Intervention | Control |
| --- | --- | --- |
| **Baseline** |  |  |
| CHU9D | 0.80 (SD = 0.11; n = 19) | 0.82 (SD = 0.09; n = 18) |
| EQ-5D-Y | 0.70 (SD = 0.22; n = 18) | 0.73 (SD = 0.22; n = 17) |
| EQ-5D-5L parent | 0.78 (SD = 0.20; n = 21) | 0.69 (SD = 0.31; n = 23) |
| ISI parent | 0.80 (SD = 0.08; n = 21) | 0.76 (SD = 0.08; n = 23) |
| **Follow-up 1** |  |  |
| CHU9D | 0.82 (SD = 0.07; n = 9) | 0.84 (SD = 0.13; n = 18) |
| EQ-5D-Y | 0.81 (SD = 0.10; n = 9) | 0.81 (SD = 0.18; n = 18) |
| EQ-5D-5L parent | 0.81 (SD = 0.07; n = 9) | 0.75 (SD = 0.28; n = 18) |
| ISI parent | 0.81 (SD = 0.08; n = 9) | 0.78 (SD = 0.07; n = 20) |
| **Follow-up 2** |  |  |
| CHU9D | 0.83 (SD = 0.07; n = 7) | 0.83 (SD = 0.15; n = 12) |
| EQ-5D-Y | 0.72 (SD = 0.35; n = 8) | 0.75 (SD = 0.23; n = 12) |
| EQ-5D-5L parent | 0.80 (SD = 0.10; n = 9) | 0.68 (SD = 0.27; n = 12) |
| ISI parent | 0.81 (SD = 0.07; n = 9) | 0.76 (SD = 0.11; n = 12) |

**Table 11:**
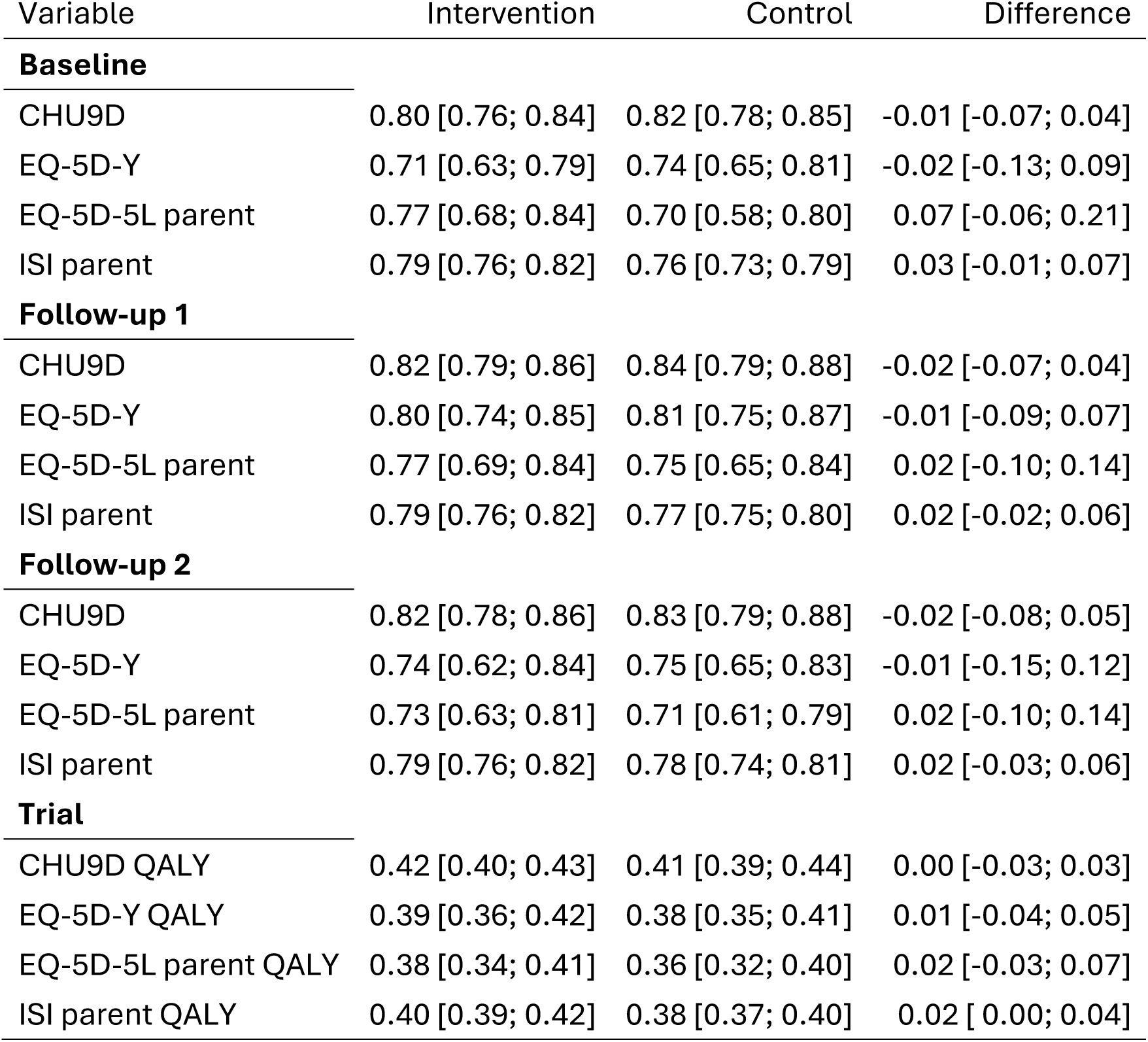
Mean unadjusted imputed and bootstrapped utility summarised by treatment group [95% confidence interval].

**Table 12:**
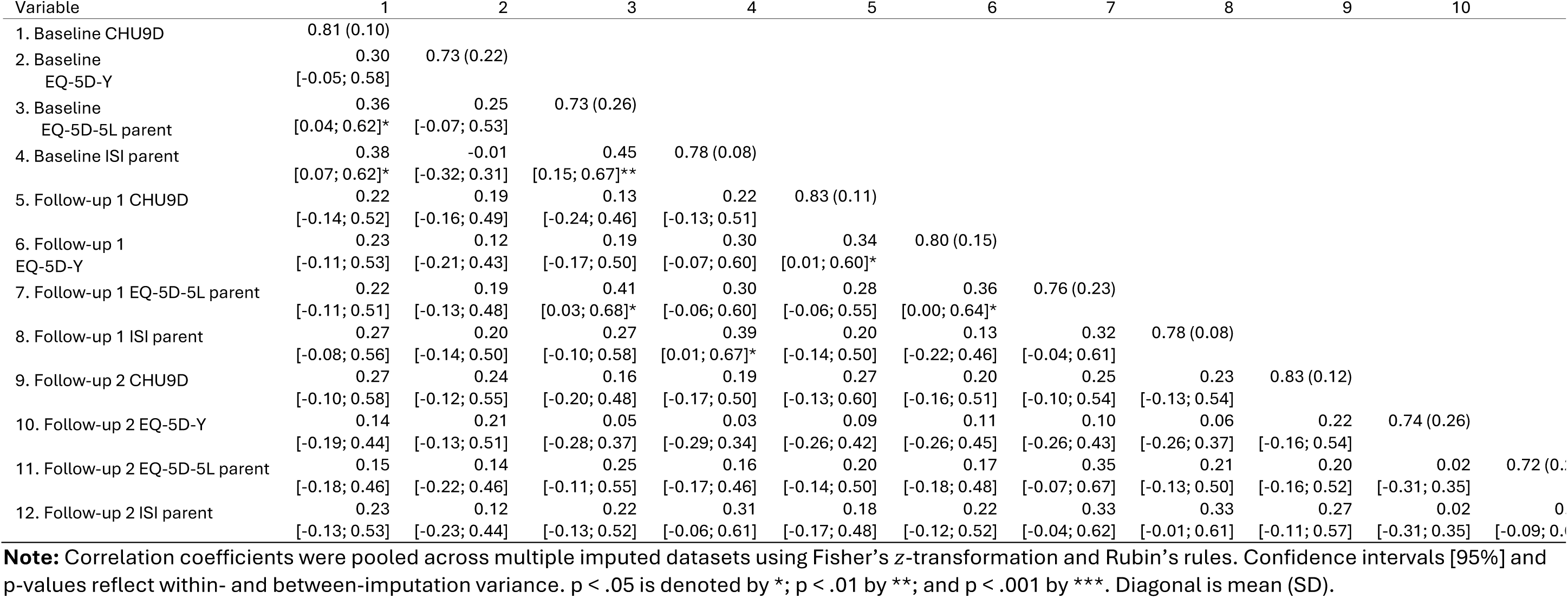
Correlation matrix of utility measures based on imputed data for primary analysis population.

### 4.5 Incremental analysis

Based on the point-estimate mean costs and QALYs, SC + COSI was more costly (£1,232 [535; 2,183]) and there was a difference of 0.0028 [ -0.0326; 0.0368] QALYs compared to SC alone. The resulting ICER is £433,167 per QALY gained and has negative net monetary benefits at both the £20,000 and £30,000 per QALY gained thresholds.

*Figure 4* shows that 57.14% of posterior draws were in the north-east quadrant, indicating that SC + COSI is associated with higher costs and more QALYs than SC alone. Based on the posterior draws, it is unlikely that COSI has lower costs (0.0009 probability), however it is possible that COSI is associated with lower QALYs (0.43 probability). In the primary analysis, at WTP thresholds of £20k and £30k per QALY, the probabilities that the intervention is cost effective from an NHS + PSS perspective are 0.01 and 0.04 respectively (*Figure 5*).

**Figure 4:**
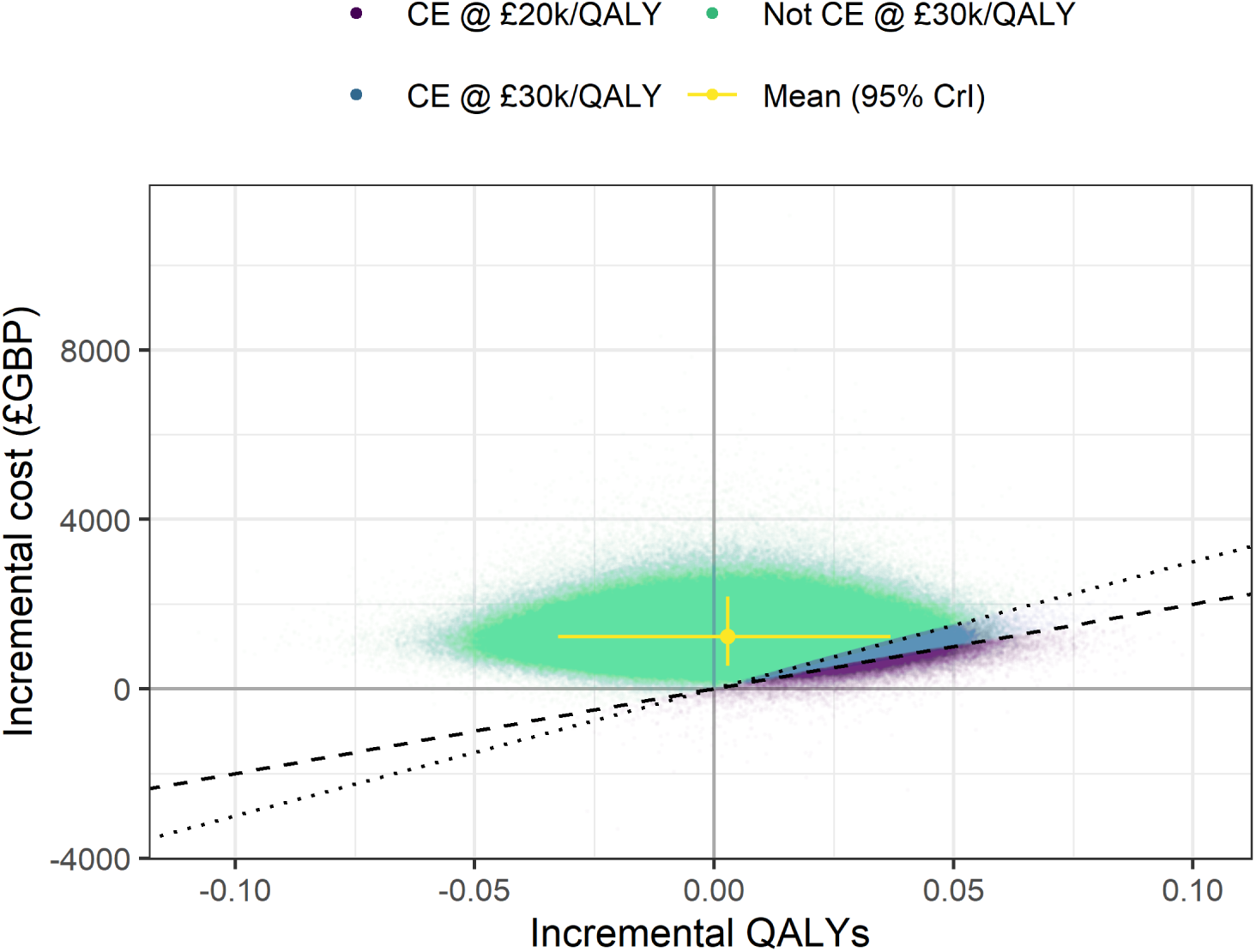
Cost-effectiveness plane. Dashed line represents £20,000 per QALY and dotted line represents £30,000 per QALY gained thresholds, CE: cost effective, CrI: credibility interval.

**Figure 5:**
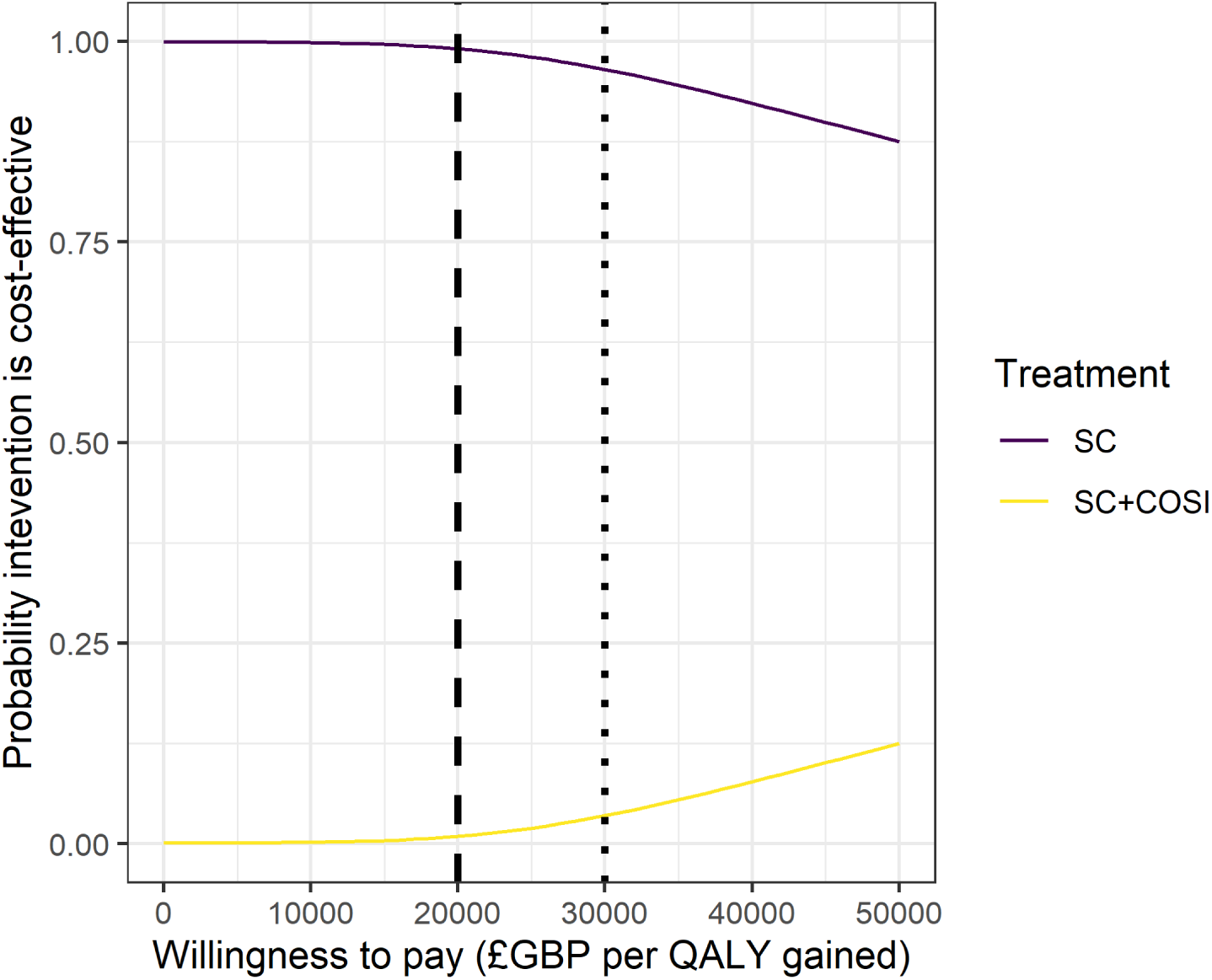
Cost-effectiveness acceptability curve for the base case analysis. Dashed line represents £20,000 per QALY and dotted line represents £30,000 per QALY.

### 4.6 Sensitivity analyses

In all the sensitivity analyses conducted, SC + COSI would not be considered cost-effective compared to SC alone at either the £20,000 or £30,000 per QALY gained thresholds. Despite the high levels of uncertainty, resulting from the high levels of missing data, the analyses are consistent in suggesting that the SC + COSI is likely to be associated with higher costs, no more QALYs than SC, and is unlikely to be cost-effective at the £30,000 per QALY threshold. *Table 13* presents the results of the sensitivity analyses that provide a single point estimate of incremental costs and/or QALYs. Sensitivity analyses that warrant more detailed explanation of their results or include figures are detailed in sub-sections below. *Figure 6* shows the distribution of posterior draws for the estimated incremental cost for each of the pre-planned scenario analyses and *Figure 7* shows the corresponding distributions for incremental QALYs. This shows that the 95% credibility intervals for incremental costs never cross zero and that the 80% credibility intervals for incremental QALYs always cross zero.

**Table 13:** Summary of incremental costs and QALYs by sensitivity analysis (mean [95% credibility interval]), incremental cost-effectiveness ratio (ICER), and incremental net monetary benefit (INMB) at £20,000 and £30,000 per QALY gained. Unless stated otherwise, n = 58.

| Scenario | Incremental costs | Incremental QALYs | ICER £/QALY | INMB @<br>£20k/QALY | INMB @<br>£30k/QALY |
| --- | --- | --- | --- | --- | --- |
| 1. Primary | 1,232 [ 535; 2,183] | 0.00 [ -0.03; 0.04] | 433,167 | -1175 | -1147 |
| 3. EQ-5D-Y | 1,232 [ 535; 2,186] | 0.01 [ -0.05; 0.06] | 165,004 | -1083 | -1009 |
| 4. Configurable COSI | 1,305 [ 608; 2,255] | 0.00 [ -0.03; 0.04] | 458,625 | -1248 | -1219 |
| 5. Static COSI | 782 [ 85; 1,732] | 0.00 [ -0.03; 0.04] | 274,755 | -725 | -696 |
| 7. Early follow-up observed (n = 85) | 1,346 [ 779; 2,120] | 0.01 [ -0.07; 0.10] | 98,776 | -1073 | -937 |
| 8. Early follow-up missing (n = 85) | 1,441 [ 690; 2,443] | 0.01 [ -0.06; 0.07] | 267,266 | -1333 | -1279 |
| 9. Parent EQ-5D-5L | 1,231 [ 536; 2,179] | 0.01 [ -0.08; 0.08] | 190,168 | -1102 | -1037 |
| 10. Parent ISI | 1,232 [ 536; 2,182] | 0.01 [ -0.03; 0.06] | 85,202 | -943 | -798 |
| 11. Societal | 1,659 [ 742; 2,870] | 0.01 [ -0.08; 0.08] | 255,376 | -1529 | -1464 |
| 12. Societal alternate absence cost | 2,251 [ 845; 4,006] | 0.01 [ -0.08; 0.08] | 346,426 | -2121 | -2056 |
| 13. RUQ costs | 738 [ 223; 1,285] | 0.00 [ -0.03; 0.04] | 258,741 | -681 | -653 |
| 14. Complete case (n = 12) | 748 [ 191; 1,333] | -0.02 [ -0.07; 0.03] | Dominated | -1110 | -1291 |
| 15. PLICS cost complete case (n = 50) | 2,359 [1,554; 3,506] | - | - | - | - |
| 16. One-level imputation | 1,238 [ 380; 2,457] | 0.01 [ -0.04; 0.05] | 209,809 | -1120 | -1061 |
| 17. Impute by group | 1,094 [ 485; 1,975] | 0.02 [ -0.02; 0.05] | 70,246 | -783 | -627 |
| 18. Impute in model | 1,362 [ 864; 1,914] | 0.00 [ -0.05; 0.04] | Dominated | -1434 | -1470 |

**Figure 6:**
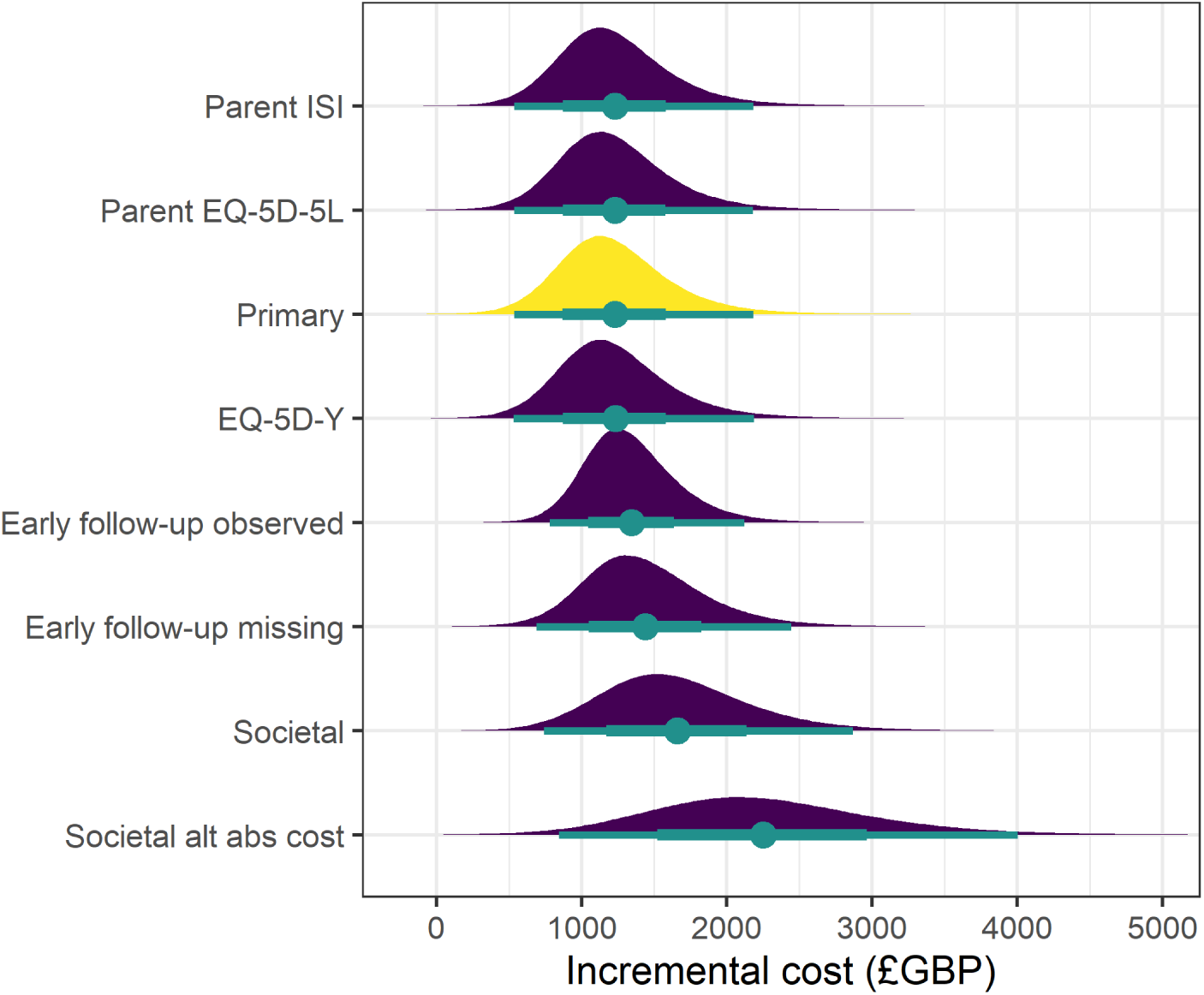
Incremental costs by sensitivity analysis, ordered by mean incremental cost. Primary analysis highlighted. Dot is mean, bars are 80% and 95% credibility intervals.

**Figure 7:**
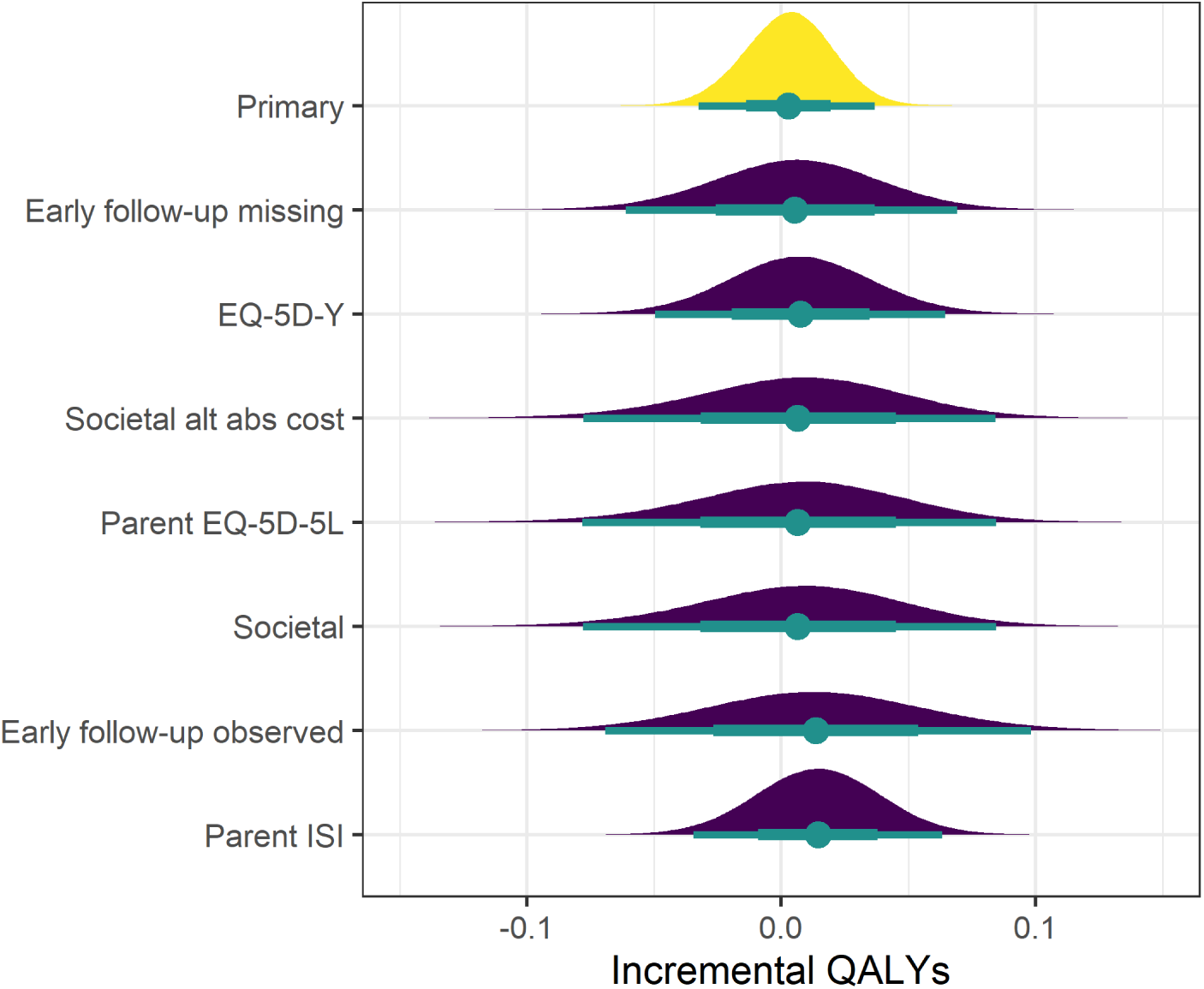
Incremental QALYs by sensitivity analysis, ordered by mean incremental QALYs. Primary analysis highlighted. Dot is mean, bars are 80% and 95% credibility intervals.

#### 4.6.1 Pre-specified

##### 4.6.1.1 Univariate cost inputs varied input by ±10% (2)

The results of the univariate sensitivity analyses presented in *Figure 8* indicated that there was no single cost input that, when varied by ±10% would have changed the cost-effectiveness decision at the £30k per QALY threshold. The intervention costs had the greatest impact on the INMB, followed by medication costs, with differences in INMB when varied by ±10% of £156.58 and £4.39 respectively. The admitted patient care costs had the smallest impact on the INMB with the difference between the two sensitivity analyses being <£0.20.

**Figure 8:**
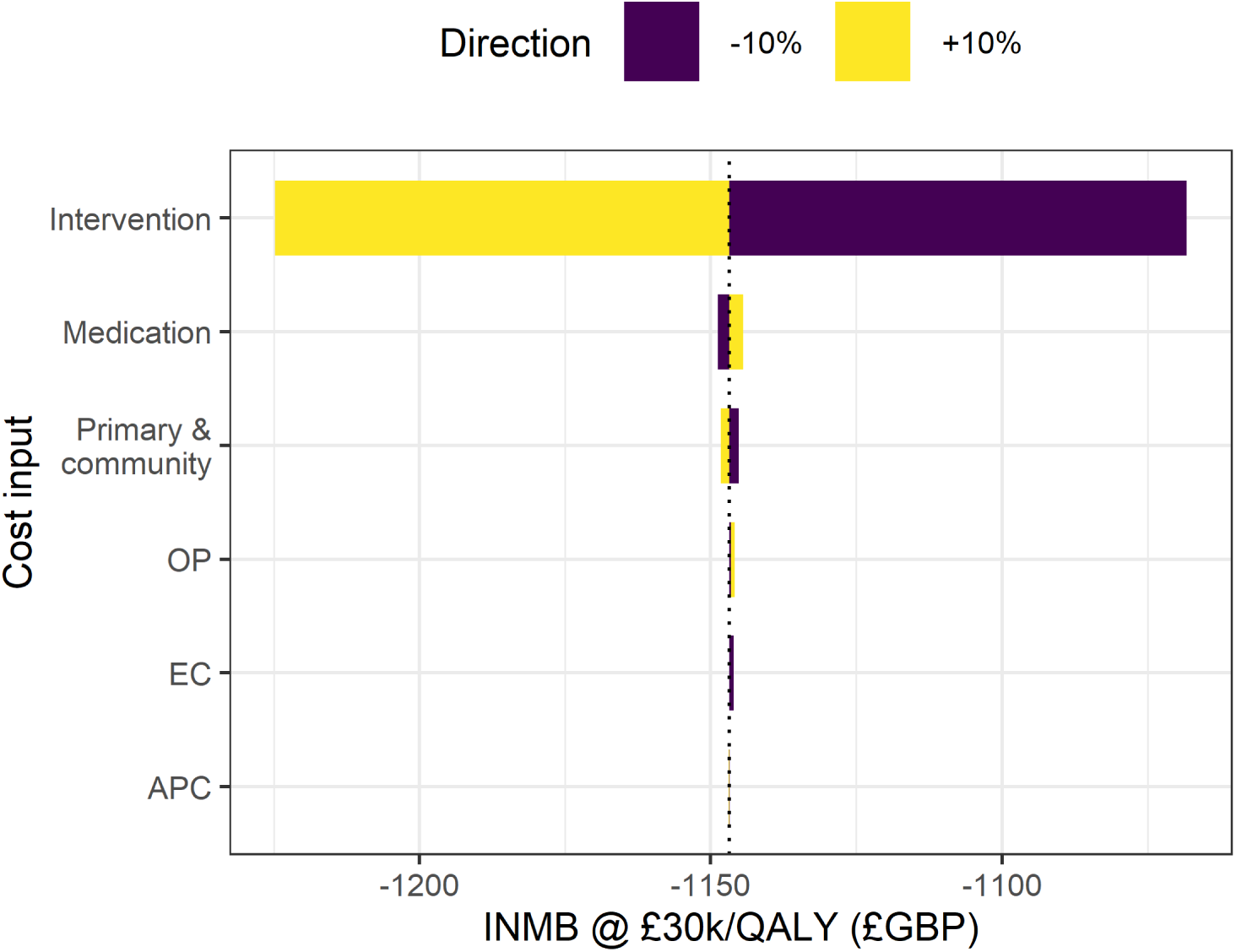
Tornado plot of univariate cost input sensitivity results at a willingness to pay threshold of £30k per QALY gained.

##### 4.6.1.2 Cost based on use of COSI (6)

*Figure 9* shows the incremental net monetary benefit (INMB) for the intervention cost based on the usage of COSI. The INMB is negative for all percentiles of the population, indicating that the intervention is not cost-effective at either the £20,000 or £30,000 per QALY gained thresholds. Given that in the primary analysis the cost of the intervention was 63.94 % of the total incremental cost, changing the cost of the intervention alone would not be expected to change a cost-effectiveness decision.

**Figure 9:**
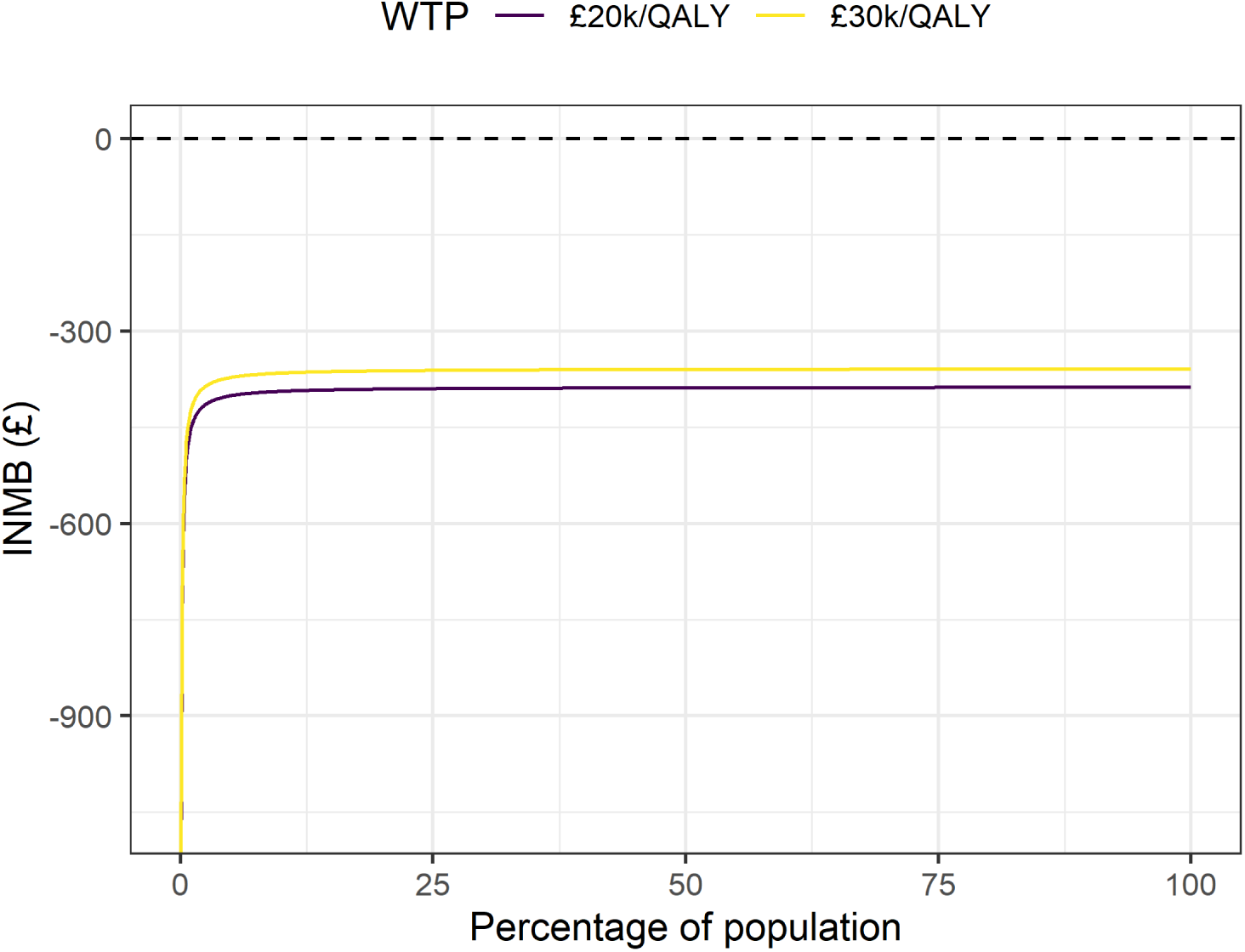
Incremental net monetary benefit with intervention cost based on use of COSI.

#### 4.6.2 Exploratory

##### 4.6.2.1 RUQ costs (13)

The sensitivity analysis that estimated secondary care costs from the RUQ data only, resulted in a lower estimate of incremental costs and a similar estimate of incremental QALYs to the primary analysis. This was partly because the estimated within-trial costs for each treatment group was lower than in the primary analysis. This suggests that RUQ data under-reports secondary care use when compared to PLICS data (*Table 14*). Another interpretation is that only those with lower resource use costs completed the RUQs and that the RUQ data is not missing at random.

There was one case where PLICS data suggested zero cost for secondary care and the RUQ data suggested greater than zero cost. This difference is very unlikely to have a substantive impact on results, therefore we did not conduct a sensitivity analysis to estimate its effect.

**Table 14:** Summary of within trial costs (£GBP) with secondary care costs estimated from PLICS and RUQ data.

| Data source | Intervention | Control |
| --- | --- | --- |
| PLICS | 2,021 [1,352; 3,455] | 790 [ 409; 1,628] |
| RUQ | 1,347 [1,010; 1,943] | 609 [ 282; 1,196] |

##### 4.6.2.2 Complete case analysis (14)

We were limited to very simple, separate, models for estimating incremental costs and QALYs due to the limited sample of complete cases (n = 12). Incremental trial costs/QALYs were each estimated with a model that included treatment group, baseline costs/utility, and use of sleep medication at randomisation as predictors. This model ignored any site effect; therefore, standard errors may be underestimated.

Including site would lead to an over specified model. The complete case analysis, as in the alternate secondary care cost analysis, suggested the incremental costs would be lower than in the primary analysis and that the incremental QALYs would be similar.

##### 4.6.2.3 PLICS costs only complete case analysis (15)

Given the small sample size in the complete case analysis, we also conducted a sensitivity analysis to estimate the incremental cost using only complete cases based on the PLICS data. PLICS data are relatively complete (maximum proportion missing = 0.12, n = 50). Here we were able to use the same model for the costs as in the primary analysis. In this instance, any bias in a complete case analysis is likely to be small. This analysis does not consider costs outside of secondary care, however, as shown in *Table 8*, the majority of costs and differences between the two groups, are associated with secondary care and the intervention.

*Table 15* shows the estimated costs for each treatment group (excluding the cost of the intervention) based on complete cases only, compared to the primary analysis. The discrepancy between the estimates for within-trial costs for the intervention group in the complete case and PLICS only complete case may simply be due to the differences in sample size, may be due to under reporting of secondary care costs in the RUQ data, or due to the exclusion of costs outside of secondary care in the PLICS only complete case analysis. Comparison with the primary analysis suggests that the exclusion of costs outside of secondary care in the PLICS only complete case analysis would only partially account for this difference.

**Table 15:** Summary of within trial costs (£) based on complete cases, excluding the cost of the intervention.

| Analysis | Intervention | Control |
| --- | --- | --- |
| Primary ($n = 58$) | 1,233 [ 564; 2,668] | 790 [ 409; 1,628] |
| Complete ($n = 12$) | 601 [ -84; 1,586] | 495 [ -33; 933] |
| PLICS only complete ($n = 50$) | 2,208 [1,132; 6,718] | 591 [ 146; 1,934] |

##### 4.6.2.4 Imputation modelling

###### 4.6.2.4.1 Imputation in model fitting (18)

Overall, the results from this within-model imputation were broadly similar to those from the primary analysis (*Table 13*). When imputation was carried out within the modelling process, there was less uncertainty in the estimate of incremental costs than in the primary analysis, whereas there was more uncertainty in the estimate of incremental QALYs compared with the primary analysis.

###### 4.6.2.4.2 MNAR (19-20)

*Figure 10* presents the mean CHU9D utility scores and within-trial QALYs for each treatment group at each timepoint across the range of *δ* tested. Considering the impact of the MNAR mechanisms tested, we can see that shifting the imputed participant utility scores had a more pronounced effect on the mean CHU9D utility and resultant QALYs for both groups than shifting primary carer utility scores.

In the participant utility MNAR mechanism, the positive relationship between the *δ* value and the mean CHU9D utility scores was consistent across both groups and timepoints, with the intervention group showing a greater increase in mean CHU9D utility scores compared to the control group. This pattern was also reflected in the within-trial QALYs, where the intervention group showed a greater increase in QALYs compared to the control group. A ceiling effect can be seen from *δ* values of 0.1 and higher, where an increasing number of values are being adjusted to their maximum of 1.0 and not the by the full value of *δ*.

There was a small positive relationship between *δ* and imputed CHU9D utility scores based on the primary carer MNAR utility mechanism, where positive values of *δ* led to slightly higher CHU9D utility scores. However, the overall effect on CHU9D was much smaller compared to the participant-based MNAR mechanism. This finding suggests that while carer utility is relevant, assumptions about participant utility missingness exert a stronger influence on the primary analysis outcomes.

**Figure 10:**
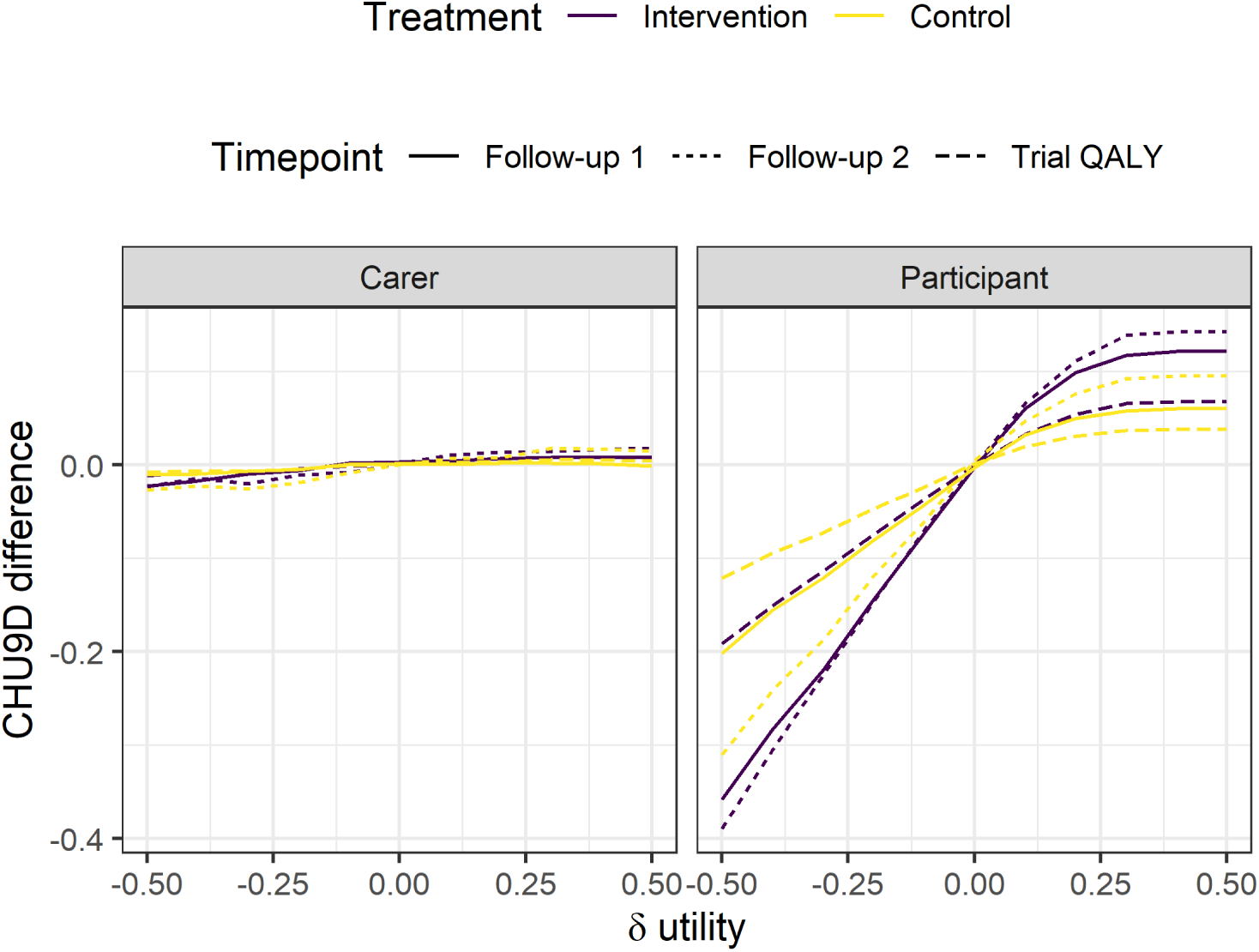
Difference in mean CHU9D utility scores and QALYs, dependent on δ applied to imputed scores.

*Figure 11* shows that even if imputations for CHU9D utility scores were increased by 0.5 (up to a maximum of 1.0), the intervention would not be considered cost-effective at a willingness to pay threshold of £30,000 per QALY gained.

**Figure 11:**
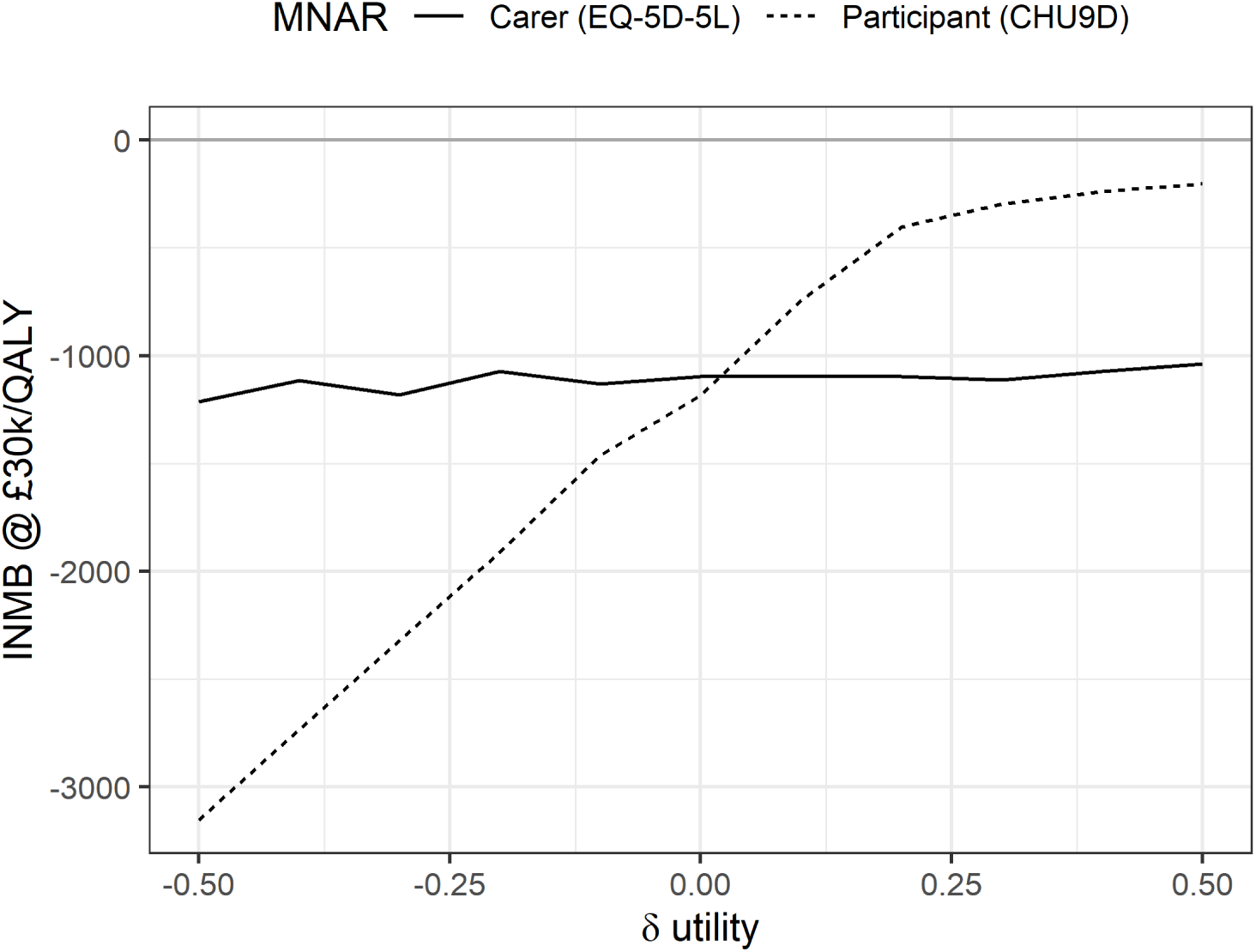
Incremental net monetary benefit (INMB) at £30k/QALY gained based on MNAR mechanism for participants’ and primary caregivers’ utility. MNAR: missing not at random, QALY: quality adjusted life year, δ: difference in imputed utility score for target MNAR mechanism.

## 5. Discussion

The results of the cost-utility analysis suggest that supplementing SC with COSI to treat epilepsy in children is unlikely to be cost-effective compared to SC alone. That is, the additional costs associated with COSI were not offset by health gains to participants or their primary carer. To be cost effective at the £30,000 per QALY threshold, the intervention would have needed to generate 0.04 additional QALYs within the trial follow-up. However, the analyses indicated that this would be very unlikely, especially in the primary analysis. This conclusion aligns with the clinical analyses, which did not demonstrate any significant benefit to SC + COSI over SC alone. The findings were consistent across all sensitivity analyses, including variations in imputation methods, assumptions about missing data, and the inclusion of different cost and outcome perspectives.

There was considerable uncertainty in both the cost and utility estimates, which is likely due to the high levels of missing data in the study. The sensitivity analyses suggest that the primary findings are most sensitive to assumptions about missing data, particularly related to participants’ utility scores. Although imputing higher utilities for participants can increase mean (incremental) QALYs, this shift is insufficient for the intervention to be considered cost-effective at the £20,000 or £30,000 per QALY gained willingness-to-pay thresholds. The results suggest that the qualitative conclusions of the primary analysis are robust to missing at random (MAR) assumptions despite the high level of missing data.

Several limitations should be considered when interpreting the findings of this study. Firstly, the reliance on self-reported data introduces the potential for reporting biases, and the high level of missing data meant we relied heavily on imputation methods.

Secondly, the small number of participants may limit the generalisability of the results. Finally, three wording issues in the RUQs were described in the methods section, these may have led to over-/under-reporting of resource use. However, because secondary care costs—estimated from PLICS data—and costs of the intervention for the SC + COSI group were the primary drivers of cost, it is unlikely that these have introduced substantive bias, particularly for the incremental analyses.

Despite the limitations, the study has several strengths. Notably, the use of PLICS data for costing provides a more accurate estimate of secondary care resource use than self-report alone, helping to reduce the impact of recall bias. Additionally, a broad spectrum of sensitivity analyses was undertaken, bolstering confidence that the overall findings—that COSI is not cost-effective at £20,000 or £30,000 per QALY—are robust to different assumptions.

## 6. Conclusion

The findings of this study indicate that supplementing standard care with COSI is unlikely to be cost-effective within an NHS and PSS or broader societal perspective. The intervention’s additional costs were not offset by improvements in quality-adjusted life years for participants or their carers, and the results were consistent across all sensitivity analyses.

## 7. Package citations

We used the following R version and packages for our analyses: R^Version 4.4.1; 53^ and the R-packages *brms*^Version 2.22.0; 43,44,45^, *broom*^Version 1.0.7; 54,55^, *broom.mixed*^Version 0.2.9.6; 55^, *dplyr*^Version 1.1.4; 56^, *emmeans*^Version 1.10.6; 57^, *eq5d*^Version 0.15.6; 58^, *forcats*^Version 1.0.0; 59^, *furrr*^Version 0.3.1; 60^, *ggokabeito*^Version 0.1.0; 61^, *ggplot2*^Version 3.5.1; 62^, *janitor*^Version 2.2.1; 63^, *kableExtra*^Version 1.4.0; 64^, *knitr*^Version 1.49; 65^, *lme4*^Version 1.1.36; 66^, *lubridate*^Version 1.9.4; 67^, *Matrix*^Version 1.7.1; 68^, *mice*^Version 3.17.0; 31,32^, *miceadds_3.17-44*^32^, *patchwork*^Version 1.3.0; 69^, *purrr*^Version 1.0.2; 70^, *Rcpp*^Version 1.0.13; 71,72^, *readr*^Version 2.1.5; 73^, *readxl*^Version 1.4.3; 74^, *RJ-2021-048*^75^, *rstan*^Version 2.32.6; 76^, *StanHeaders*^Version 2.32.10; 77^, *stringr*^Version 1.5.1; 78^, *tibble*^Version 3.2.1; 79^, *tidybayes*^Version 3.0.7; 80^, *tidyr*^Version 1.3.1; 81^, *tidyverse*^Version 2.0.0; 82^ and *writexl*^Version 1.5.1; 83^.

